# Drivers of Diagnostic Variation in a Digital Global Kidney Transplant Reader Study

**DOI:** 10.64898/2026.07.09.26357318

**Authors:** Rianne Hofstraat-Boersma, Romy du Long, Giorgio Buzzanca, Adeyemi Adefidipe Abiola, Sam Albadri, Zahabia Ali, Ahmed Altaleb, Andrea Angioi, Sultana Gulshana Banu, Marc Barry, Ami R. Bhalodia, Paola Bianco, Verena Broecker, Roman Bülow, Bertrand Chauveau, Guoli Chen, Boonyarit Cheunsuchon, Giovanna M. Crisi, Simin Daneshvar, Amelie Dendooven, Pouneh Dokouhaki, Cinthia B. Drachenberg, Alton B. Farris, Sophie Ferlicot, Sandrine Florquin, Francesco Fontana, Jean-Baptiste Gibier, Ian W. Gibson, Saurabh Gujarathi, Allen R. Hendricks, Sufia Husain, Jaynul Islam, Wesam Ismail, Geetha Jagannathan, Johannes Klager, Nicolas Kozakowski, Adriana Krizova, Anila A. Kurien, Boomi Kwon, Vincenzo L’Imperio, Felipe L. Ledesma, Julia P. Low, Jonne Martin, Shweta S. Mehta, Nidia Messias, Salvatore E. Mignano, Monik E. Miranda, Gilbert Moeckel, Chiara Pala, Brendon Price, Marc A. Ranson, Andrea N. Rodriguez, Joris J.T.H. Roelofs, Avi Rosenberg, Alireza Sadeghipour, Miroslav Sekulic, Suman Setty, Michael Sheaff, Maria F.S. Soares, Jerasit Surintrspanont, George Terinte-Balcan, Francesca Testa, David O. Henriquez Ticas, Maria F. Toniolo, Dominique van Midden, Ramya K. Velagapudi, Seethalakshmi Viswanathan, Saskia von Stillfried, Chih-Ying Wu, Nienke Kalverboer, Imke B. Bruns, Giulia Callegaro, Bob van de Water, Danny van der Helm, Aiko P.J. de Vries, Hessel Peters-Sengers, Jesper Kers

## Abstract

**Background:** Diagnostic interpretation of kidney allograft biopsies using the Banff classification remains variable, but the determinants of this variability are not fully defined. We performed a global, fully digital multi-reader study to identify the principal drivers of disagreement in Banff-based assessment.

**Methods:** Thirty-six kidney transplant biopsies were independently scored by 67 renal pathologists on a standardized digital platform. Readers assessed Banff lesions on hematoxylin and eosin, periodic acid–Schiff, and Jones’ silver stains; final diagnostic categories were assigned using prespecified Banff-based decision rules. Interobserver agreement was quantified with Gwet’s agreement coefficient (AC) statistics. Determinants of diagnostic agreement were evaluated using pairwise mixed-effects logistic regression, and reader similarity was examined by principal component analysis (PCA) with post hoc molecular annotation.

**Results:** Agreement for final diagnostic categories was moderate (Gwet’s AC1, 0.55; 95% CI, 0.47–0.63). Lesion-level agreement varied substantially, with lowest agreement for selected threshold-dependent inflammatory or semi-quantitative lesions, including interstitial inflammation in areas of IFTA, peritubular capillaritis and arteriolar hyalinosis. Diagnostic concordance differed markedly across biopsies, indicating strong case-level heterogeneity. In pairwise models, differences in active inflammatory and vascular lesion scoring were the strongest correlates of diagnostic disagreement; reader experience and geography contributed minimally. Principal component analysis showed reader variation was organized along two dominant axes: a rejection-calling threshold axis linked mainly to tubulointerstitial inflammatory injury, and a T cell-mediated (TCMR/TI) and antibody-mediated/microvascular (AMR/MVI) inflammation-oriented phenotypic classification axis.

**Conclusion:** Interobserver variation in Banff-based kidney transplant biopsy assessment is structured rather than random and driven mainly by how readers threshold and integrate key inflammatory lesion compartments rather than experience or geographic location.

## INTRODUCTION

Accurate interpretation of kidney allograft biopsies is central to the diagnosis and management of transplant injury and remains a cornerstone of clinical decision-making in kidney transplantation. The Banff classification provides a standardized framework for semi-quantitative assessment of histologic lesions and has become the dominant international reference standard for kidney allograft pathology in both clinical practice and trials.^1,2^ Despite this standardization, variability in the application of Banff criteria remains a persistent challenge, with substantial differences in lesion grading reported even among experienced renal pathologists.^3–5^ More recent multicenter studies have confirmed only fair inter-rater agreement for several key Banff inflammation scores, including interstitial inflammation (i), total inflammation (ti), and inflammation in areas of interstitial fibrosis and tubular atrophy (i-IFTA).^6^

Similar limitations in reproducibility have been described in native kidney biopsy interpretation. Systematic reviews of lupus nephritis classification have shown poor-to-moderate interobserver agreement, and recent studies have highlighted low concordance for ordinal scoring of fibrosis and inflammatory lesions, underscoring that diagnostic variability represents a broader challenge within renal pathology.^7,8^ In the transplant setting, however, such variability may have particularly direct consequences because Banff lesion interpretation determines diagnostic classification, therapeutic decision-making, and patient stratification in clinical trials. Misinterpretation of Banff criteria for antibody-mediated rejection (AMR), for example, has been shown to affect both diagnostic categorization and treatment selection in clinical practice.^9^

Several intrinsic properties of Banff lesion scoring make reproducibility difficult to achieve. Banff lesions are assessed on semi-quantitative ordinal scales that rely on visual estimation and threshold-based categorization within morphologically heterogeneous tissue compartments. Subtle boundaries between score categories, together with the low prevalence of some lesions, increase susceptibility to reader-dependent interpretation. Differences in field selection, compartment emphasis, training background, diagnostic thresholding, and local diagnostic culture may further contribute to variability. Although digital pathology platforms now enable standardized slide presentation and large-scale multi-reader studies, digitization alone does not eliminate the interpretive subjectivity inherent to semi-quantitative histopathologic scoring.

While interobserver variability in Banff lesion scoring has been recognized for decades, most prior studies have focused primarily on quantifying the magnitude of agreement rather than identifying the determinants of diagnostic disagreement. Existing reproducibility studies have generally been limited in scale, often restricted to national or regional reader panels, and have typically examined only a subset of Banff lesions, most often microvascular inflammation or transplant glomerulopathy in the context of AMR.^6,10,11^ In addition, reproducibility has often been summarized using κ-based statistics, which may perform less well when category distributions are highly imbalanced, as is common for Banff lesions and diagnostic categories.^12^ However, agreement coefficients alone provide limited insight into the mechanisms that generate diagnostic variation. In practice, disagreement in kidney transplant biopsy interpretation may arise from several distinct sources, including intrinsic case complexity, differences in how pathologists interpret specific Banff lesions, and variation in how lesions are weighted in diagnostic decision-making, depending on the clinical context.

To address these questions, we conducted an international, fully digital multi-reader study in which 67 renal pathologists from across the globe independently evaluated 36 kidney allograft biopsies across three routinely used histochemical stains (hematoxylin and eosin, periodic acid-Schiff, and Jones’ silver). Slides were assessed in blinded, randomized fashion for each pathologist using a standardized online platform. The analytical strategy was designed to move beyond descriptive agreement estimates by evaluating variability at multiple levels, including biopsy cases, lesion interpretation, and reader behavior. The objectives of this study were therefore to (1) quantify interobserver agreement across Banff lesions and final diagnostic categories, (2) identify the principal determinants of diagnostic disagreement, including lesion-level interpretive differences and case-level variability, and (3) characterize the structure of reader similarity to determine whether reproducible interpretive patterns underlie variability in Banff-based diagnosis.

## METHODS

### Study Design and Case Selection

We conducted a global, fully digital, blinded interobserver reproducibility study of Banff lesion scoring and diagnostic classification in kidney allograft biopsies. Thirty-six kidney transplant biopsies performed between 2021 and 2024 were retrospectively selected from the pathology archive of the Leiden University Medical Center (LUMC). Inclusion criteria were the availability of hematoxylin and eosin (H&E)-, periodic acid–Schiff (PAS)-, and Jones’ silver-stained sections for each case, documented donor-specific antibody (DSA) and C4d status (positive or negative), adequate tissue quality for Banff lesion assessment and availability of formalin-fixed paraffin-embedded (FFPE) tissue for TempO-Seq whole-transcriptome data. Based on the histological diagnosis in the clinical report, the complete spectrum of rejection phenotypes was selected to ensure an adequate representation of morphological variation along the spectrum of T cell-mediated and antibody-mediated rejection. All slides were pseudonymized and digitized using a whole-slide scanner (Philips IntelliSite Ultra-Fast Scanner, Philips, the Netherlands) at a resolution of 0.25 µm per pixel. Before inclusion, an experienced renal pathologist reviewed cases to confirm technical adequacy and suitability for Banff scoring (JK). The study was conducted in accordance with the Declaration of Helsinki and was approved by the local ethics and privacy committee of LUMC, with a waiver of informed consent because of the retrospective and pseudonymized nature of the material (approval number RP25.044).

### Reader Recruitment and Digital Scoring Workflow

Renal pathologists were recruited internationally through the Renal Pathology Society website and professional social media channels (LinkedIn and X). Interested participants registered through an online Google Form. Participation was voluntary and eligibility required experience in renal pathology and self-reported familiarity with the Banff classification in their daily practice. Reader characteristics, including geographic region and years of renal pathology experience, were collected for prespecified subgroup analyses. All biopsies were assessed using the SlideScore platform (SlideScore B.V., Amsterdam, the Netherlands). For each case, H&E, PAS, and Jones’ silver stains were available to the reader simultaneously. Trichrome is not regularly used in clinical practice (including the LUMC) and was therefore not included. Case order was randomized separately for each reader to minimize systematic order bias, fatigue, or learning effects. Readers were blinded to all clinical information, case identifiers, original diagnoses, DSA and C4d status and the assessments of other participants. Scoring was completed independently within a predefined study window (3 months). Readers were instructed to score all cases once without revisiting previous assessments (locked in the platform). For the primary pooled agreement analyses, readers were included if they completed the full biopsy set. Only scores by pathologists who finished the assessment within the time window were included for analysis (N = 67).

### Histopathological Scoring and Lesion Definitions

Histopathological scoring was performed according to the Banff lesion definitions used in the 2022 Banff classification, which remained unchanged in the 2024 Banff update.^1,13^ Readers scored the prespecified Banff lesions required for diagnostic classification, including glomerulitis (*g*), peritubular capillaritis (*ptc*), interstitial inflammation (*i*), tubulitis (*t*), total inflammation (*ti*), inflammation in areas of interstitial fibrosis and tubular atrophy (*i*-IFTA), tubulitis in areas of interstitial fibrosis and tubular atrophy (*t*-IFTA), interstitial fibrosis (*ci*), tubular atrophy (*ct*), interstitial fibrosis and tubular atrophy (IFTA), vascular fibrous intimal thickening (*cv*), mesangial matrix expansion (*mm*), arteriolar hyalinosis (*ah*), intimal arteritis (*v*), and transplant glomerulopathy (*cg*) (full list in **Table S1**). Lesion scores were recorded on the ordinal scales defined by the Banff classification. Readers were provided with written Banff lesion definitions within the scoring interface but received no clinical information or additional interpretive guidance. For each biopsy, readers generated an integrated Banff assessment based on review of all three stains. Handling of non-assessable structures was predefined; for example, when arteries were absent, vascular lesion scores were recorded as not assessable (N/A) and treated as missing in agreement analyses. Percentage-based lesion scores, including IFTA and *ti*, were recorded in 10% increments and subsequently converted to ordinal Banff categories using predefined thresholds of 0%, 1%–25%, 26%–50%, and >50% to enable ordinal agreement analyses and diagnostic rule assignment. For each reader-biopsy combination, final Banff diagnostic categories were derived algorithmically from prespecified Banff indicator variables based on the reader’s lesion scores and biopsy-level donor-specific antibody and C4d status, which were not provided to readers during scoring. We implemented a decision-tree approach based on Yoo et al., with modifications to reflect the 2024 Banff diagnostic criteria.^13,14^ When multiple diagnostic labels were present, final diagnostic categories were assigned using a predefined hierarchical resolution strategy prioritizing mixed rejection, followed by AMR, TCMR, borderline changes, and no rejection. Diagnostic categories were encoded using a fixed set of category levels to ensure consistency across analyses and resampling procedures. The four histological indices (Activity Index, Chronicity Index, TCMR/TI index and AMR/MVI index) were calculated for each individual biopsy and reader according to the definition used by Vaulet and colleagues and integrated with the C4d status (not provided during scoring).^15–17^

### Quantification of Interobserver Agreement

Interobserver agreement for individual Banff lesion scores was quantified using Gwet’s agreement coefficient for ordinal data (AC2) with linear weighting.^12^ AC2 was selected because Banff lesion scores are ordinal and frequently exhibit imbalanced categorical distributions. Linear weighting was used to apply a proportional penalty according to the distance between ordinal score categories. Observed agreement proportions were calculated as descriptive measures to complement chance-corrected agreement coefficients. Interobserver agreement for final Banff diagnostic categories was quantified using Gwet’s AC1 for nominal data and was prespecified as the primary measure of diagnostic agreement. To characterize biopsy-level heterogeneity in diagnostic reproducibility, case-level concordance was defined as the proportion of readers assigning the most frequently selected diagnostic category for each biopsy. Agreement for the continuous histological indices, including the Activity Index, Chronicity Index, TCMR/TI index, and AMR/MVI index, was assessed using intraclass correlation coefficients (ICC). Because some index values were unavailable when required biopsy structures were non-assessable, variance components were estimated using linear mixed-effects models rather than standard ANOVA-based ICC estimation with listwise deletion. Biopsy case and reader were modeled as random effects. Absolute-agreement and consistency ICCs were calculated to distinguish overall agreement from systematic differences in reader scoring thresholds. Detailed ICC formulas, missing-data handling, and bootstrap procedures are provided in the **Supplementary Methods**.

### Subgroup Analyses

Agreement coefficients were calculated within prespecified reader strata defined by geographic region and years of experience in renal pathology. Experience was categorized into predefined groups (<5 years, 5–10 years, and >10 years). To account for unequal subgroup sizes, subgroup analyses were performed using case-level bootstrap resampling. In each bootstrap replicate, biopsy cases were sampled with replacement while preserving all reader assessments within sampled cases. Agreement coefficients were recalculated within each stratum, and pairwise differences between strata were derived from bootstrap distributions with percentile-based 95% confidence intervals.

### Pairwise Determinants of Diagnostic Agreement

To identify determinants of diagnostic agreement, reader assessments were transformed into a pairwise comparison dataset in which each observation represented two readers evaluating the same biopsy. For each reader pair and biopsy, diagnostic agreement was defined as assignment of the same final Banff diagnostic category. This yielded 79,596 reader-pair-by-biopsy observations. Mixed-effects logistic regression models were used to estimate the probability of diagnostic agreement, with agreement specified as a binary outcome. Models included reader-level covariates, including absolute difference in years of renal pathology experience and geographic similarity, defined as practice on the same continent. A random intercept for biopsy case was included to account for clustering of reader-pair observations within biopsies and for variability in case difficulty. Absolute differences in Banff lesion scores between readers were included as fixed effects to quantify the contribution of lesion-level interpretive variation to diagnostic agreement. Model fit was compared using Akaike’s information criterion (AIC), and effect sizes were expressed as odds ratios with 95% confidence intervals.

### Structural Analysis of Reader Similarity

To characterize latent patterns of diagnostic interpretation among readers, principal component analysis was applied to the reader-by-reader diagnostic similarity matrix. Diagnostic assignments were first harmonized into final Banff diagnostic categories, and a case-by-reader diagnosis matrix was constructed. Pairwise diagnostic similarity between readers was then quantified across all biopsies using Gwet’s AC1, generating a symmetric reader-by-reader agreement matrix. Principal component analysis was performed on this matrix to identify the dominant dimensions of variation in reader interpretation. The proportion of variance explained by each component was calculated from the corresponding eigenvalues. To aid interpretation of the reader axes, reader coordinates on the principal components were related to diagnostic proportions, mean Banff lesion scores, and composite histological indices.

### Molecular correlates of pairwise inter-reader variability

To provide post hoc biological annotation of reader-derived patterns of diagnostic variability, biopsy-level transcriptomic pathway activity and inferred immune cell composition (deconvolved with CIBERSORTx) were integrated with the reader similarity analysis after principal component construction. Molecular data were not used to derive the reader similarity matrix or principal component coordinates. Biopsy transcriptomic profiling was performed using TempO-Seq on formalin-fixed paraffin-embedded tissue. In short, probe sequences were reconciled with the most recent NCBI gene symbol annotation (release 2024-12-10). Probes exhibiting <10 counts per million (CPM) in >95% of samples were discarded, leaving 14,220 high-confidence probes. Counts from probes mapping to the same gene were subsequently aggregated to obtain gene-level expression values (N genes = 12,740). Batch effects arising from the five sequencing runs were mitigated with ComBat-seq (sva v3.56.0, Bioconductor). Biopsy-level pathway activity was quantified using gene set variation analysis with predefined transplant-related gene sets (Halloran et al., Molecular Microscope Diagnostic System (MMDx); **Tables S2-3**). Inferred cell-type composition was estimated from normalized expression data from the same transcriptomic dataset using CIBERSORTx in absolute mode with a custom immune and kidney-resident cell-type signature matrix (**Table S4**).^18^ Absolute CIBERSORTx scores were used as per-sample abundance estimates. For each reader, molecular summary features were derived from the biopsies assigned to prespecified diagnostic groupings by that reader. These reader-level molecular summaries were then merged with the corresponding reader principal component coordinates. Molecular annotation analyses were performed for the leading principal components of reader variability. P-values were adjusted for multiple testing within each molecular analysis using the Benjamini-Hochberg false discovery rate method. Detailed definitions of the reader-level molecular summaries, diagnostic groupings, and principal-component-specific annotation analyses are provided in the **Supplementary Methods**.

### Statistical Analysis

All analyses were performed in R (version 4.5.1) using scripted workflows to ensure reproducibility. Categorical and ordinal variables were summarized as counts and percentages, and continuous or composite numerical variables were summarized as medians with interquartile ranges or means with standard deviations, as appropriate. Interobserver agreement was quantified using Gwet’s agreement coefficients, with AC2 applied to ordinal Banff lesion scores and AC1 applied to nominal final diagnostic categories. Observed agreement proportions were calculated descriptively. Agreement for composite histological indices was assessed using intraclass correlation coefficients estimated from linear mixed-effects variance components. Associations involving ordinal, continuous, or composite numerical variables were evaluated using rank-based correlation methods. Associations involving categorical grouping variables were evaluated using non-parametric group-comparison tests. Pairwise determinants of diagnostic agreement were analyzed using mixed-effects logistic regression, with diagnostic agreement modeled as a binary outcome and biopsy case included as a random effect. Model fit was compared using Akaike’s information criterion, and regression effect sizes were reported as odds ratios with 95% confidence intervals. Principal component analysis was applied to the reader-by-reader diagnostic similarity matrix to identify dominant axes of reader variability. Where applicable, 95% confidence intervals were derived from analytical estimates, bootstrap distributions, or model-based estimates. P-values for molecular annotation analyses were adjusted for multiple testing using the Benjamini-Hochberg false discovery rate method. Analyses of molecular correlates of reader variability were considered exploratory and post hoc. Additional details, full analytical scripts and package versions are provided in the **Supplementary Material**.

## RESULTS

### Study Cohort and Reader Characteristics

The study design and a heatmap of pairwise comparisons are summarized in **Figure 1**. The cohort included 36 biopsies from 34 patients (**Table 1**). Of the 34 patients, 16 (47%) were female, the median recipient age was 52 years (interquartile range (IQR) = 43 - 60 years) and the median donor age was 58 (IQR = 48 - 66). Fifteen grafts (44%) were from deceased donations, of which 7 were donations after cardiac death (21% of total). Two patients (6%) underwent re-transplantation. Two biopsies (6%) were protocol biopsies, 11 (31%) were DSA positive (all *de novo*; i.e., no preformed DSA were detected). Eleven biopsies (31%) were C4d positive, of which 7 (19%) were both DSA and C4d positive. For 14 cases an SV40 staining was performed and negative in all. The median number of glomeruli per biopsy was 19 (range 7 - 57). The final clinicopathological diagnoses were as follows: 7 active AMR, 6 chronic active AMR, 6 acute TCMR, 6 chronic active TCMR and 11 index biopsies with other diagnoses. Of the 12 TCMR cases, 5 had a v-lesion and were therefore classified as TCMR grade >1. The other disease category consisted of pure acute tubular necrosis (n = 3), acute tubular necrosis with nephrocalcinosis (n = 1), acute tubular necrosis with hypertensive vascular changes (n = 1), acute tubular necrosis with recurrence of IgA nephropathy (n = 1), recurrence of primary podocytopathy/minimal change disease (n = 1), pyelonephritis (n = 1), calcineurin-inhibitor toxicity (n= 1), interstitial fibrosis and tubular atrophy grade 2 not otherwise specified (NOS, n = 1) and nonspecific changes/normal histology (n = 1).

**Figure 1.**
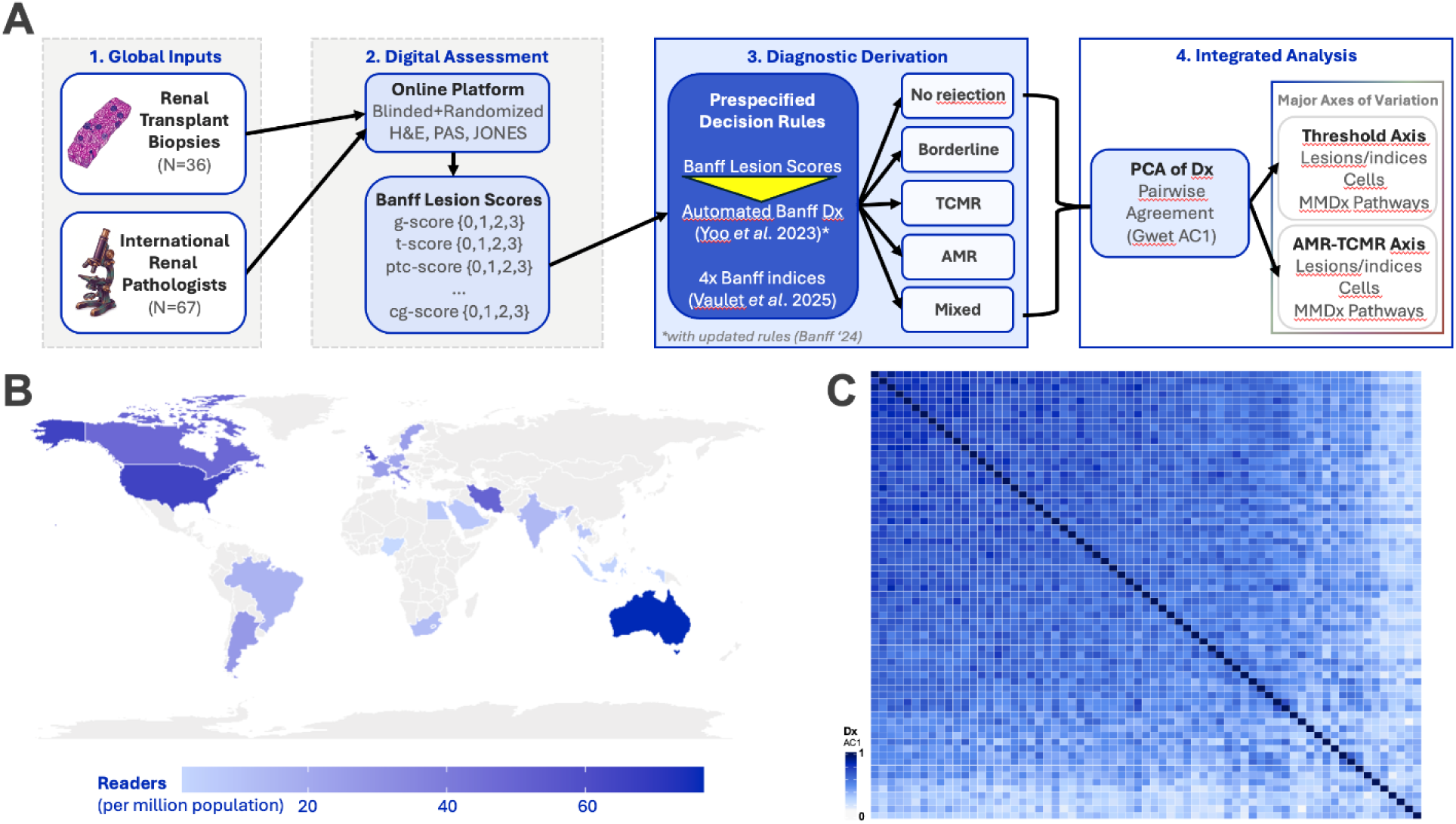
Global digital multi-reader study design and distribution of pairwise disagreement. **A.** Schematic overview of the reader study (panel A). **B.** Global distribution of renal pathologists in the study, expressed as a the number per million population. **C.** Heatmap of pairwise comparisons in diagnosis agreement (Gwet AC1) across all readers. Each cell displays a single reader pair.

**Table 1.**
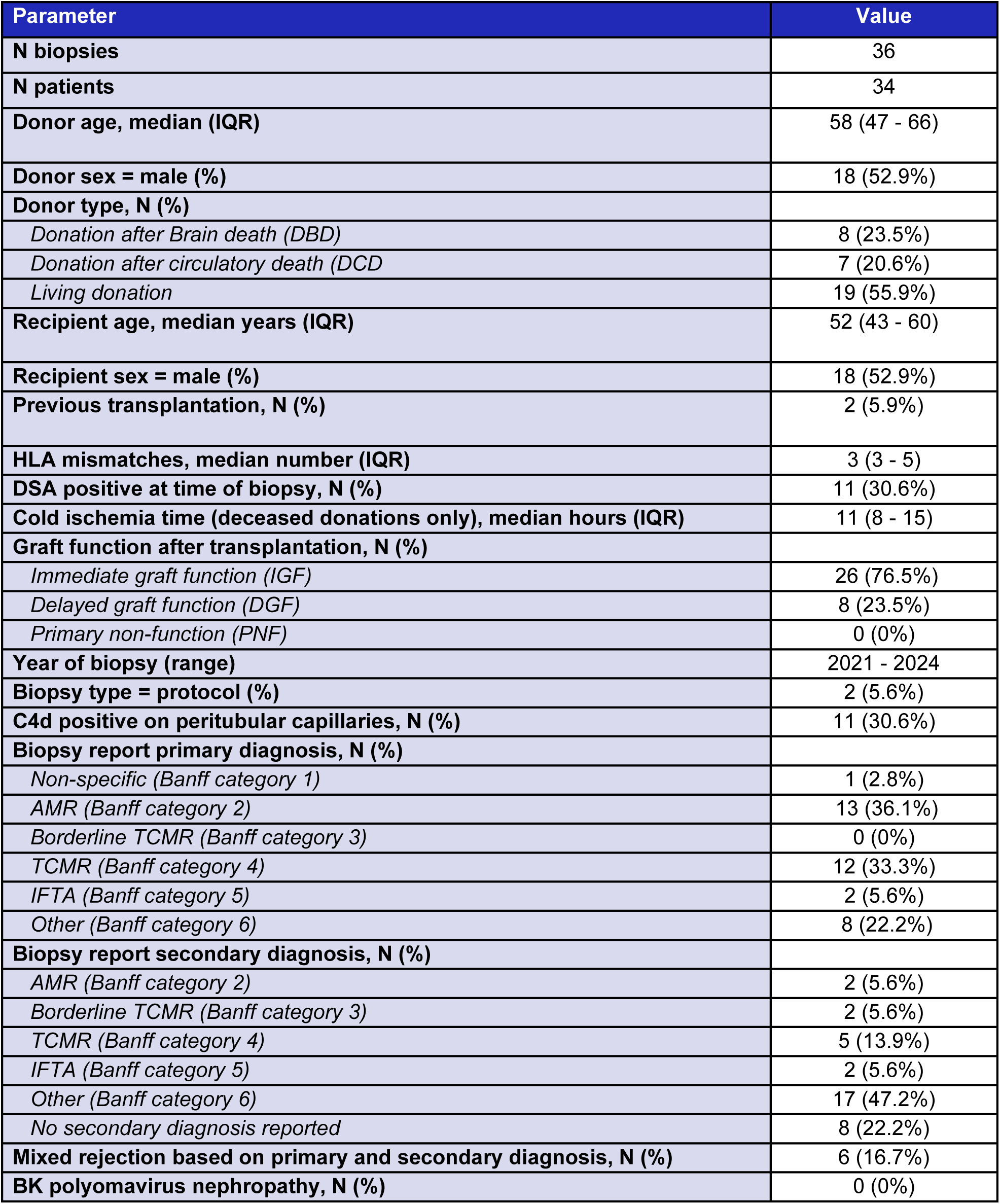
Baseline characteristics of the patients and biopsies.

| Parameter | Value |
| --- | --- |
| **N biopsies** | 36 |
| **N patients** | 34 |
| **Donor age, median (IQR)** | 58 (47 - 66) |
| **Donor sex = male (%)** | 18 (52.9%) |
| **Donor type, N (%)** |  |
| *Donation after Brain death (DBD)* | 8 (23.5%) |
| *Donation after circulatory death (DCD)* | 7 (20.6%) |
| *Living donation* | 19 (55.9%) |
| **Recipient age, median years (IQR)** | 52 (43 - 60) |
| **Recipient sex = male (%)** | 18 (52.9%) |
| **Previous transplantation, N (%)** | 2 (5.9%) |
| **HLA mismatches, median number (IQR)** | 3 (3 - 5) |
| **DSA positive at time of biopsy, N (%)** | 11 (30.6%) |
| **Cold ischemia time (deceased donations only), median hours (IQR)** | 11 (8 - 15) |
| **Graft function after transplantation, N (%)** |  |
| *Immediate graft function (IGF)* | 26 (76.5%) |
| *Delayed graft function (DGF)* | 8 (23.5%) |
| *Primary non-function (PNF)* | 0 (0%) |
| **Year of biopsy (range)** | 2021 - 2024 |
| **Biopsy type = protocol (%)** | 2 (5.6%) |
| **C4d positive on peritubular capillaries, N (%)** | 11 (30.6%) |
| **Biopsy report primary diagnosis, N (%)** |  |
| *Non-specific (Banff category 1)* | 1 (2.8%) |
| *AMR (Banff category 2)* | 13 (36.1%) |
| *Borderline TCMR (Banff category 3)* | 0 (0%) |
| *TCMR (Banff category 4)* | 12 (33.3%) |
| *IFTA (Banff category 5)* | 2 (5.6%) |
| *Other (Banff category 6)* | 8 (22.2%) |
| **Biopsy report secondary diagnosis, N (%)** |  |
| *AMR (Banff category 2)* | 2 (5.6%) |
| *Borderline TCMR (Banff category 3)* | 2 (5.6%) |
| *TCMR (Banff category 4)* | 5 (13.9%) |
| *IFTA (Banff category 5)* | 2 (5.6%) |
| *Other (Banff category 6)* | 17 (47.2%) |
| *No secondary diagnosis reported* | 8 (22.2%) |
| **Mixed rejection based on primary and secondary diagnosis, N (%)** | 6 (16.7%) |
| **BK polyomavirus nephropathy, N (%)** | 0 (0%) |

### Distribution of Automatically Assigned Diagnoses

Diagnostic assignments were imbalanced across Banff categories reflecting distribution of rejection diagnoses in real clinical practice, with a limited number of categories accounting for most reader-case evaluations. Across 2412 individual reads, the most frequently assigned diagnosis was *no rejection* (886/2412, 36.7%), followed by *mixed rejection* (542/2412, 22.5%) and *chronic active AMR* (471/2412, 19.5%). *Chronic active TCMR* accounted for 223/2412 reads (9.2%), *borderline TCMR* for 148/2412 (6.1%), *active AMR* for 77/2412 (3.2%), *acute TCMR* for 57/2412 (2.4%), and *probable AMR* for 8/2412 (0.3%).

### Overall Interobserver Agreement

Overall interobserver agreement for final Banff diagnostic categories was moderate, with a Gwet’s AC1 of 0.55 (95% CI, 0.47-0.63). Agreement varied across individual Banff lesions. The highest agreement was observed for *v* (Gwet’s AC2 = 0.88; 95% CI, 0.84–0.93), *cg* (Gwet’s AC2 = 0.84; 95% CI, 0.77–0.92), and *g* (Gwet’s AC2 = 0.71; 95% CI, 0.61–0.81), whereas the lowest agreement was observed for *i-IFTA* (Gwet’s AC2 = 0.36; 95% CI, 0.24–0.48), *ptc* (Gwet’s AC2 = 0.49; 95% CI, 0.34–0.64), and *ah* (Gwet’s AC2 = 0.51; 95% CI, 0.38–0.64; **Table S5**). These findings indicate that substantial variability remained in diagnostic assignment despite the use of standardized Banff criteria and automated diagnostic label allocation. Agreement appeared lower for lesions requiring integration of multiple assessment thresholds, such as both lesion extent and severity. For example, *ptc* requires assessment of whether more than 10% of peritubular capillaries are inflamed, followed by grading according to the number of inflammatory cells in the most severely involved capillary. Lesions requiring such multi-step assessment (*mm*, *i*, *t*, *i-IFTA*, *t-IFTA*, *ptc*, and *ah*) showed lower agreement than lesions based predominantly on a single assessment threshold (*g*, *ti*, *ci*, *ct*, *IFTA*, *cg*, *v*, and *cv*), with mean Gwet’s AC2 values of 0.58 (SD = 0.11) versus 0.67 (SD = 0.13), respectively (one-sided t test, p = 0.05).

### Case-level Heterogeneity in Diagnostic Agreement

Concordance for automatically assigned diagnostic labels varied markedly across biopsies, with majority proportions ranging from near-complete agreement to substantially lower concordance, indicating pronounced case-level heterogeneity in reproducibility (**Figure 2A**). Low-concordance cases occurred across multiple diagnostic categories rather than being confined to a single classification group. Heatmap visualization likewise showed disagreement was concentrated in a subset of biopsies with heterogeneous labeling across readers, whereas other cases were classified more consistently (**Figure 2B**). These findings indicate that interobserver variability was strongly case-dependent and concentrated in biopsies near interpretive or phenotypic boundaries.

**Figure 2.**
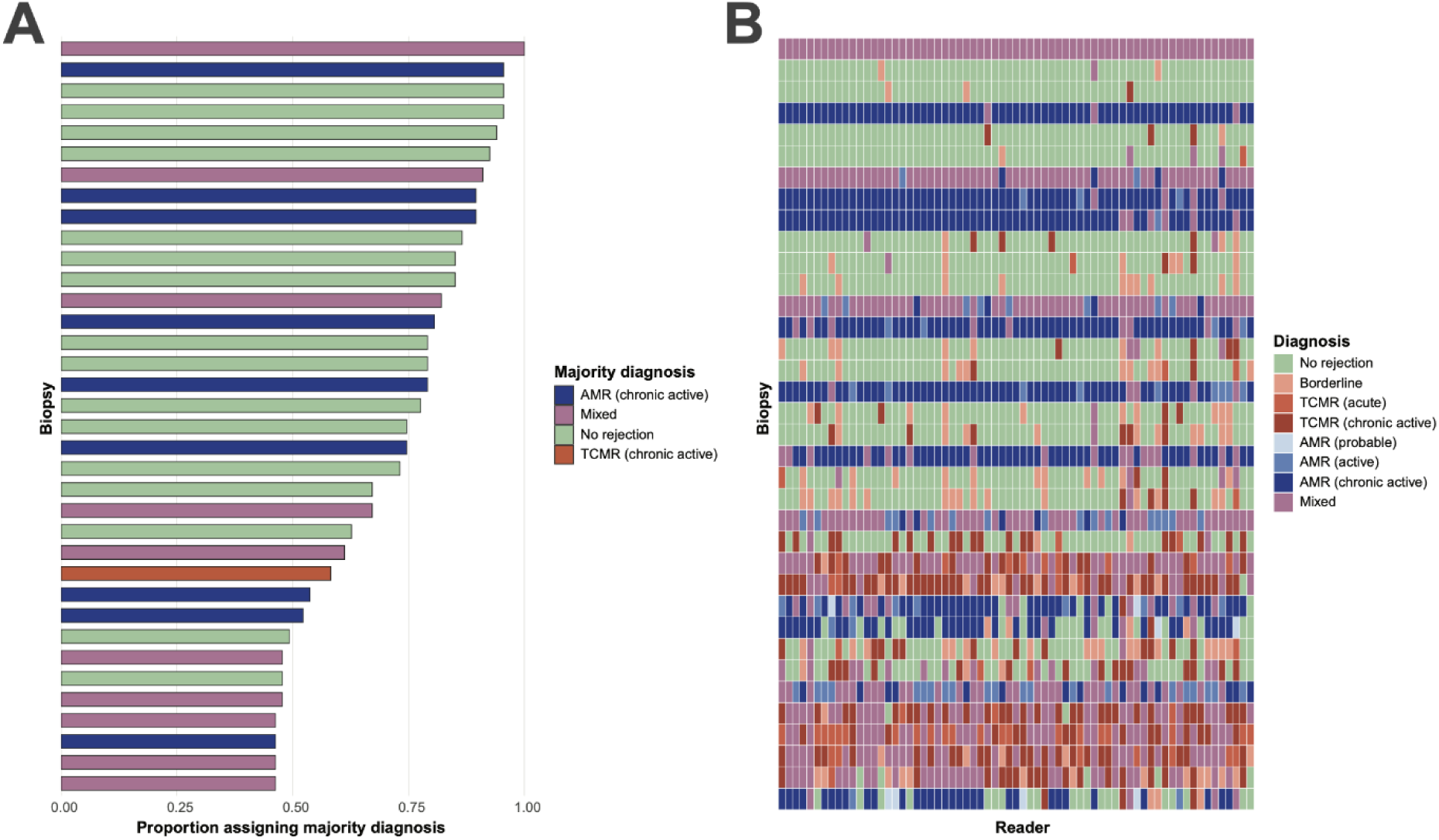
Case-level heterogeneity in diagnostic concordance across readers. **A.** Case-level diagnostic concordance for each biopsy, defined as the proportion of readers assigning the majority diagnosis. The majority diagnosis was defined as the final diagnostic category assigned most frequently to that biopsy across all readers. Biopsies are ordered by concordance and colored according to their majority diagnosis. **B.** Reader-by-case heatmap of final diagnostic assignments shown in the same concordance-based biopsy order as in panel A. Each row represents a biopsy, and each column represents a reader; colors indicate the assigned final diagnostic category.

### Interobserver agreement of the Activity, Chronicity, TCMR/TI and AMR/MVI indices

In the most recent 2024 iteration of the Banff classification, the potential implementation of an Activity Index (AI) and Chronicity Index (CI) was proposed. In a recent study by Vaulet et al., these AI and CI, together with novel indices representing the spectrum of diagnostic probability along the TCMR/TI and AMR/MVI axes, were extensively assessed and externally validated. Since all four indices are by definition dependent on multiple Banff lesion scores, theoretically, *systematic* disagreement across multiple lesions can accumulate. Dendrogram clustering based on the correlation matrix of pairwise lesion differences indeed shows a hierarchical structure of intercorrelated (dis)agreement patterns (**Figure S1**). These correlations in pairwise disagreement were most pronounced for chronic and active tubulointerstitial lesions (IFTA, ci, ct, i-IFTA, t-IFTA and t, i and ti, respectively). **Figure 3A** shows that the agreement for the 4 indices is relatively high with absolute agreements ranging from 0.62 (95% CI 0.46 – 0.71) for the Chronicity Index to 0.83 (95% CI 0.76 – 0.89) for the AMR/MVI index. The effect of systematic bias in scoring is relatively minor (the difference between absolute agreement and consistency). **Figure 3B** shows the ranking of each pathologist compared to the mean consensus for each of the 4 indices. The absolute mean bias in pairwise comparison between pathologists was at maximum 4.50/17 (26%) for the Activity Index, 4.28/15 (29%) for the Chronicity Index, 2.33/10 (23%) for the AMR/MVI index and 2.73/10 (27%) for the TCMR/TI index.

**Figure 3.**
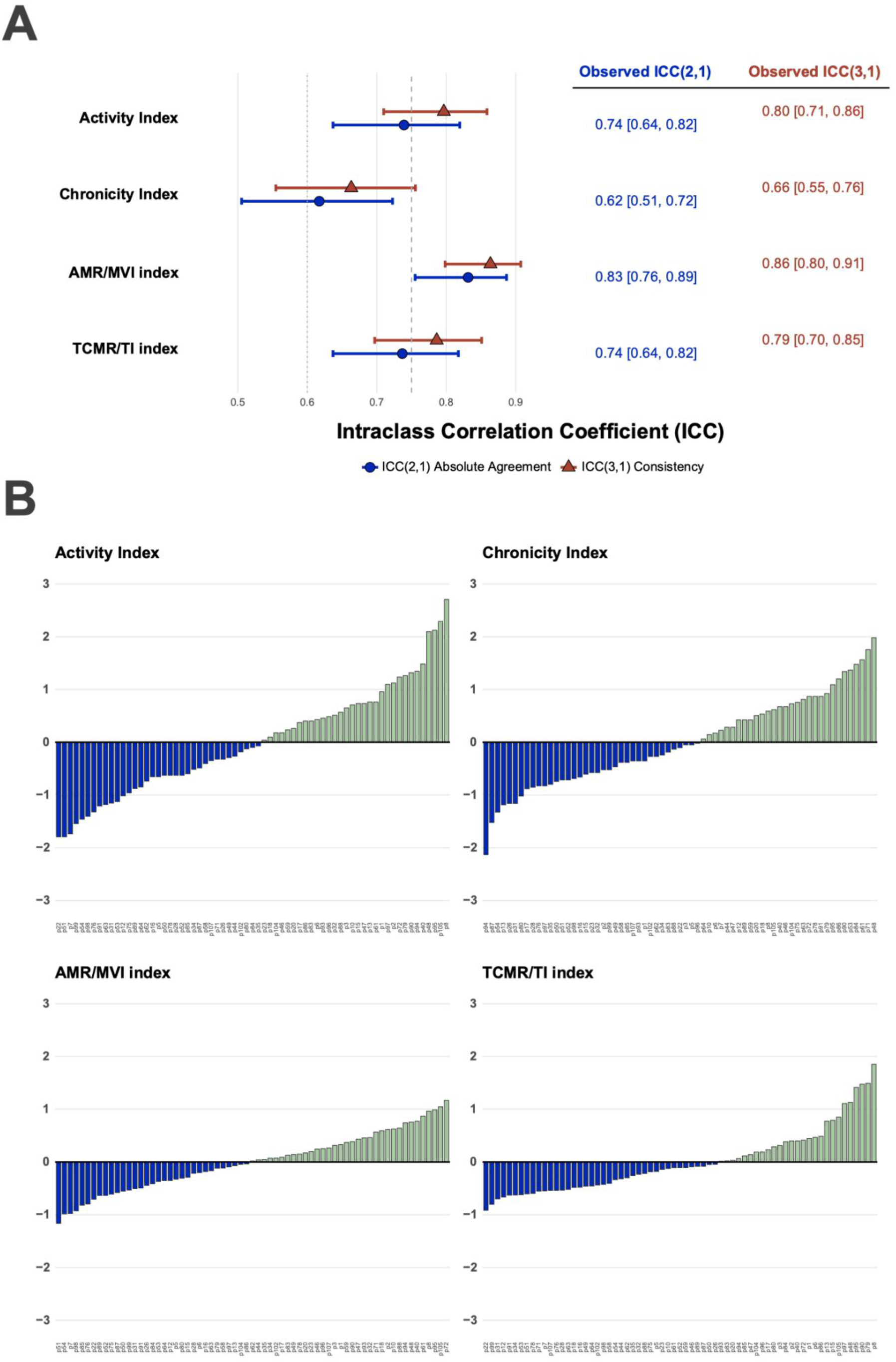
Interobserver agreement of the Activity, Chronicity, AMR/MVI and TCMR/TI indices. **A.** Absolute and relative intraclass concordance (ICC) for the 4 indices showing only minor loss of agreement, indicative of only minor additive/cumulative effect of individual lesion disagreement. **B.** Reader ranking according to their absolute deviation of the mean consensus index.

### Lesion-level Differences Explain Pairwise Diagnostic (Dis)agreement

In models containing only reader-level variables, pairwise diagnostic agreement showed only modest associations with experience difference and shared continent between pathologists, whereas substantial between-case variability was present (random intercept variance 2.01), indicating strong case-dependent effects. Adding lesion-level interpretation differences markedly improved model fit (AIC 91,234 for the base model vs 71,102 for the full lesion model), and a reduced core lesion model captured most of this improvement (AIC 72,118). In the core model, disagreement in key inflammatory and vascular lesions was strongly associated with lower odds of concordant diagnosis, particularly for arteritis (*v*; OR, 0.15; 95% CI, 0.14–0.16; p < 0.001), tubulitis (*t*; OR, 0.45; 95% CI, 0.43–0.46; p < 0.001), interstitial inflammation (*i*; OR, 0.53; 95% CI, 0.52–0.55; p < 0.001), and peritubular capillaritis (*ptc*; OR, 0.54; 95% CI, 0.52–0.55; p < 0.001; **Figure 4A; Table S6**). By contrast, chronic structural lesions had substantially smaller effects, with only modest negative associations for *ci* and *ct* in the core model (OR, 0.92 and 0.93, respectively; **Figure 4A; Table S6**). These patterns were maintained in the full lesion model, indicating that a limited set of active inflammatory and vascular lesions accounted for most of the explanatory signal, whereas additional lesions contributed comparatively little (**Figure 4B; Table S6**). Reader-level covariates remained near null after adjustment (experience difference OR approximately 1.00; same continent OR approximately 0.96; **Table S6**). Residual case-level variability persisted, consistent with an additional contribution from intrinsic biopsy complexity. Bootstrap subgroup analyses likewise showed only modest differences in agreement by experience and geography, without a consistent pattern suggesting that reader demographics were primary drivers of diagnostic variation (**Figure S2; Table S7-9**). Overall, these findings indicate that diagnostic disagreement was associated predominantly with differences in interpretation of active inflammatory and vascular lesions rather than with reader experience or geography.

**Figure 4.**
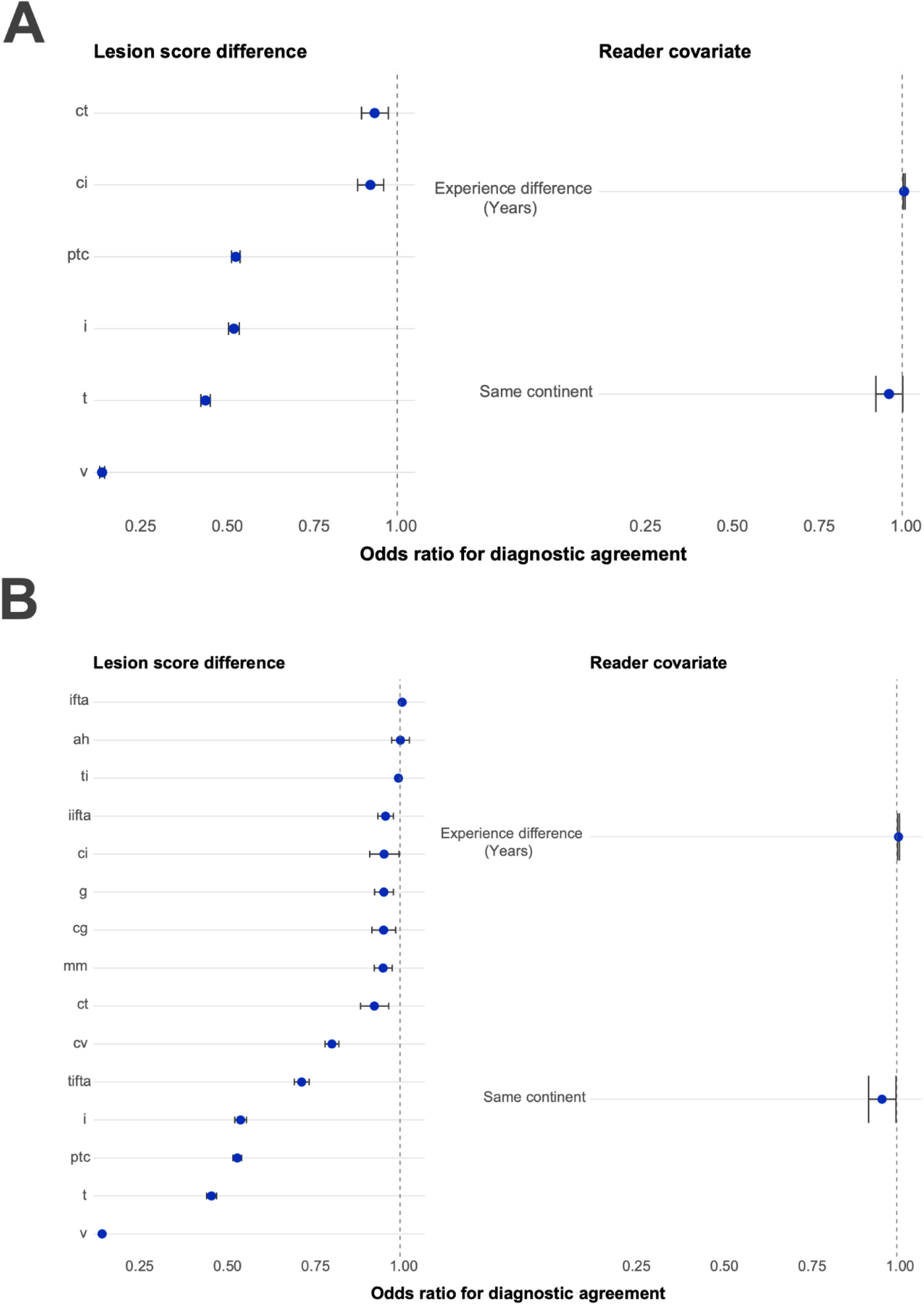
Impact of Banff lesion interpretation differences on pairwise diagnostic agreement in mixed-effects models. Forest plots showing odds ratios (ORs) and 95% confidence intervals from mixed-effects logistic regression models of pairwise diagnostic agreement, with biopsy case included as a random intercept. The outcome was whether a reader pair assigned the same final diagnosis to a given biopsy. Lower odds ratios indicate lower odds of diagnostic agreement with increasing disagreement in lesion scoring. **A. Core mixed-effects model.** Predictors included the absolute between-reader differences in key Banff lesion scores (*v, ptc, i, t, ci,* and *ct*), together with reader-level covariates for difference in years of experience and shared continent. Odds ratios are shown per 1-unit increase in absolute lesion score difference. **B. Full mixed-effects model.** Predictors included the absolute between-reader differences in Banff lesion scores across acute, chronic, and composite lesions, together with reader-level covariates for difference in years of experience and shared continent. Odds ratios are shown per 1-unit increase in absolute lesion score difference

### Deconvolution of the Histological Drivers of Rejection Diagnosis Disagreement

To identify the major patterns underlying diagnostic disagreement, we performed pairwise comparisons between readers across the 36 biopsies and applied principal component analysis (PCA) to the reader similarity matrix derived from pairwise Gwet’s AC1 agreement. To visualize the structure underlying this analysis, pairwise reader similarity was displayed as a reader-by-reader heatmap ordered by PC1 (**Figure 5A**). This revealed a gradual shift in inter-reader similarity across the PC1 axis rather than discrete reader clusters, indicating that diagnostic variation was organized along continuous interpretive gradients. Inter-reader variation was dominated by two principal dimensions (**Figure S3**). PC1 explained 60.1% of the total variance and PC2 explained 12.9%, together accounting for 73.0% of the structure in reader similarity (**Table S10**). PC1 captured a reader-level threshold for assigning rejection-related diagnoses, rather than a strict biological separation between rejection and no rejection. Higher PC1 values were associated with a greater tendency to classify biopsies as rejection, as reflected by positive correlations with *any rejection* diagnosis (ρ = 0.70, p < 0.001), the TCMR/TI index (ρ = 0.64, p < 0.001), interstitial inflammation (*i*; ρ = 0.62, p < 0.001), and arteritis (*v*; ρ = 0.40, p < 0.001), and by an inverse correlation with *no rejection* diagnosis (ρ = −0.70, p < 0.001). PC1 was therefore interpreted as a rejection-calling threshold axis, with higher values indicating a lower reader threshold for assigning rejection-related categories, particularly in biopsies with tubulointerstitial inflammatory injury (**Table 2**). PC2 captured phenotypic orientation among rejection-related classifications, with strongest associations along the AMR/MVI spectrum. Higher PC2 values correlated strongly with peritubular capillaritis (*ptc*; ρ = 0.79, p < 0.001), mixed rejection diagnoses (ρ = 0.78, p < 0.001), the AMR/MVI index (ρ = 0.71, p < 0.001), and glomerulitis (*g*; ρ = 0.51, p < 0.001). PC2 also correlated with any rejection diagnosis (ρ = 0.45, p < 0.001) and the TCMR/TI index (ρ = 0.44, p < 0.001), indicating that this component did not simply separate rejection from non-rejection, but rather differentiated how readers resolved rejection-type biopsies across TCMR-oriented, AMR/microvascular-oriented, and mixed-rejection patterns. PC2 was therefore interpreted as an AMR/MVI-oriented phenotypic classification axis among rejection-related diagnoses (**Table 2**). Visualization of reader coordinates in PCA space supported these interpretations, with gradients in rejection-associated and TCMR-related features aligned predominantly along PC1 and gradients in AMR/microvascular-associated features aligned predominantly along PC2 (**Figure S4**). Higher-order components explained substantially less variance (PC3 3.9%, PC4 3.0%) and were not considered central to the main interpretive structure (**Table S11**). Reader characteristics showed minimal association with PCA coordinates: years of experience were not correlated with PC1 (ρ = 0.00, p = 0.99), PC2 (ρ = 0.11, p = 0.39), PC3 (ρ = −0.10, p = 0.40), or PC4 (ρ = 0.15, p = 0.24), and geographic region was not associated with either major component (PC1, p = 0.67; PC2, p = 0.66). Visualization of the PCA space colored by experience and geography similarly showed no clear clustering by these variables (**Figure 5B**). Overall, the PCA demonstrated that inter-reader variability was structured along two major biologically interpretable axes rather than by demographic characteristics.

**Figure 5.**
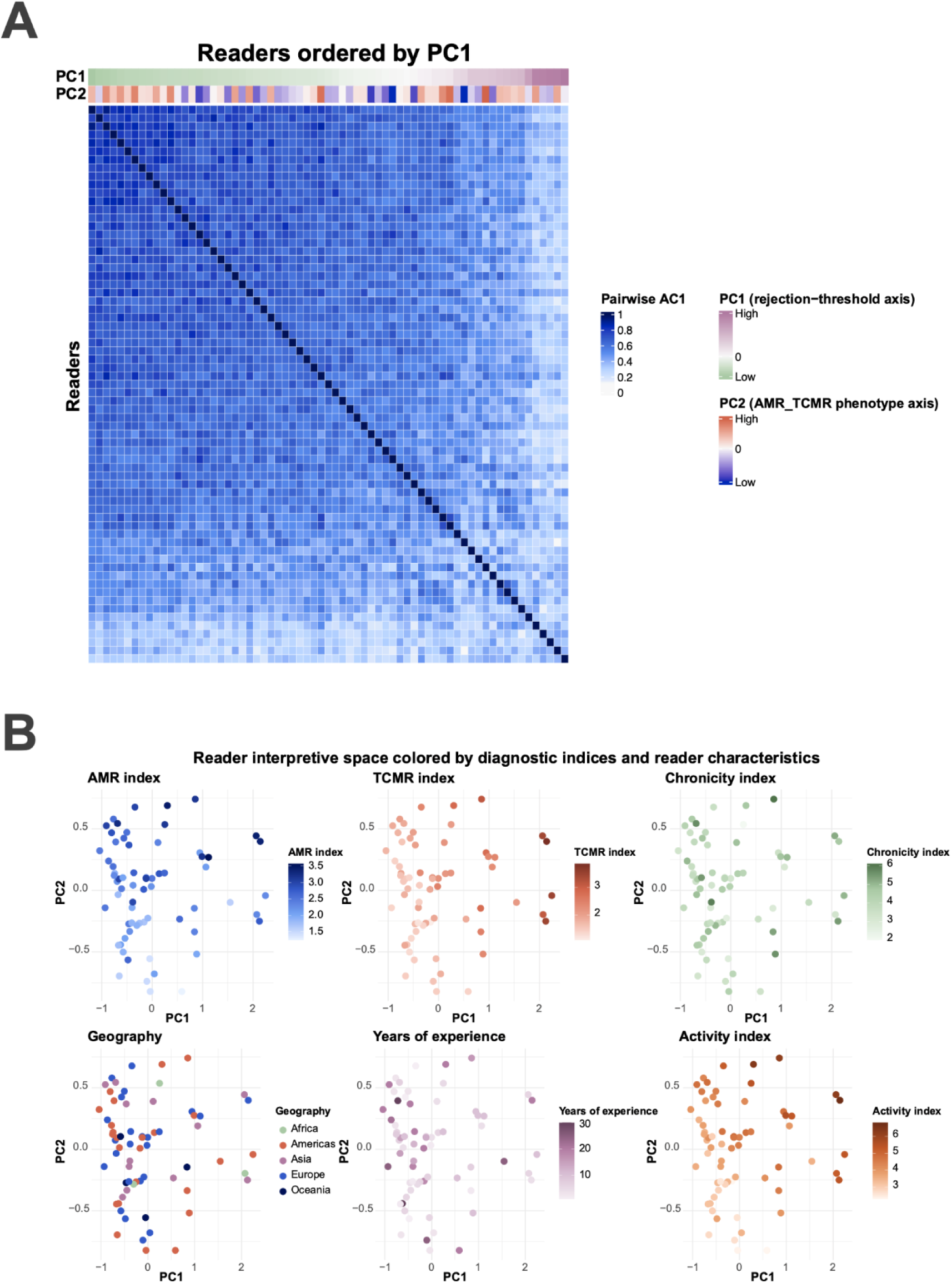
Distribution of pairwise diagnostic (dis)agreement patterns. **A.** Reader similarity structure ordered by the dominant rejection-calling threshold axis. Heatmap of pairwise reader similarity based on Gwet’s AC1 for final Banff diagnostic classification. Each row and column represents an individual reader, and color intensity indicates the magnitude of pairwise diagnostic agreement across the 36 biopsies. Readers are ordered according to their PC1 coordinates derived from principal component analysis of the reader similarity matrix. Annotation bars indicate reader position along PC1 and PC2. PC1 represented the dominant rejection-calling threshold axis, reflecting variation in readers’ tendency to assign rejection-related diagnoses, whereas PC2 represented an AMR/MVI-oriented phenotypic classification axis. The gradual shift in pairwise agreement across the ordered matrix supports a continuous structure of reader interpretation rather than discrete reader clusters. **B.** Multipanel scatterplots showing readers projected onto the first two principal components (PC1 and PC2) derived from the reader-by-reader pairwise Gwet’s AC1 matrix for final Banff diagnostic classification. Each point represents one reader and is colored according to the corresponding reader-level mean AMR/MVI index, TCMR/TI index, Chronicity Index, Activity Index, geographic region, or years of renal pathology experience.

**Table 2.**
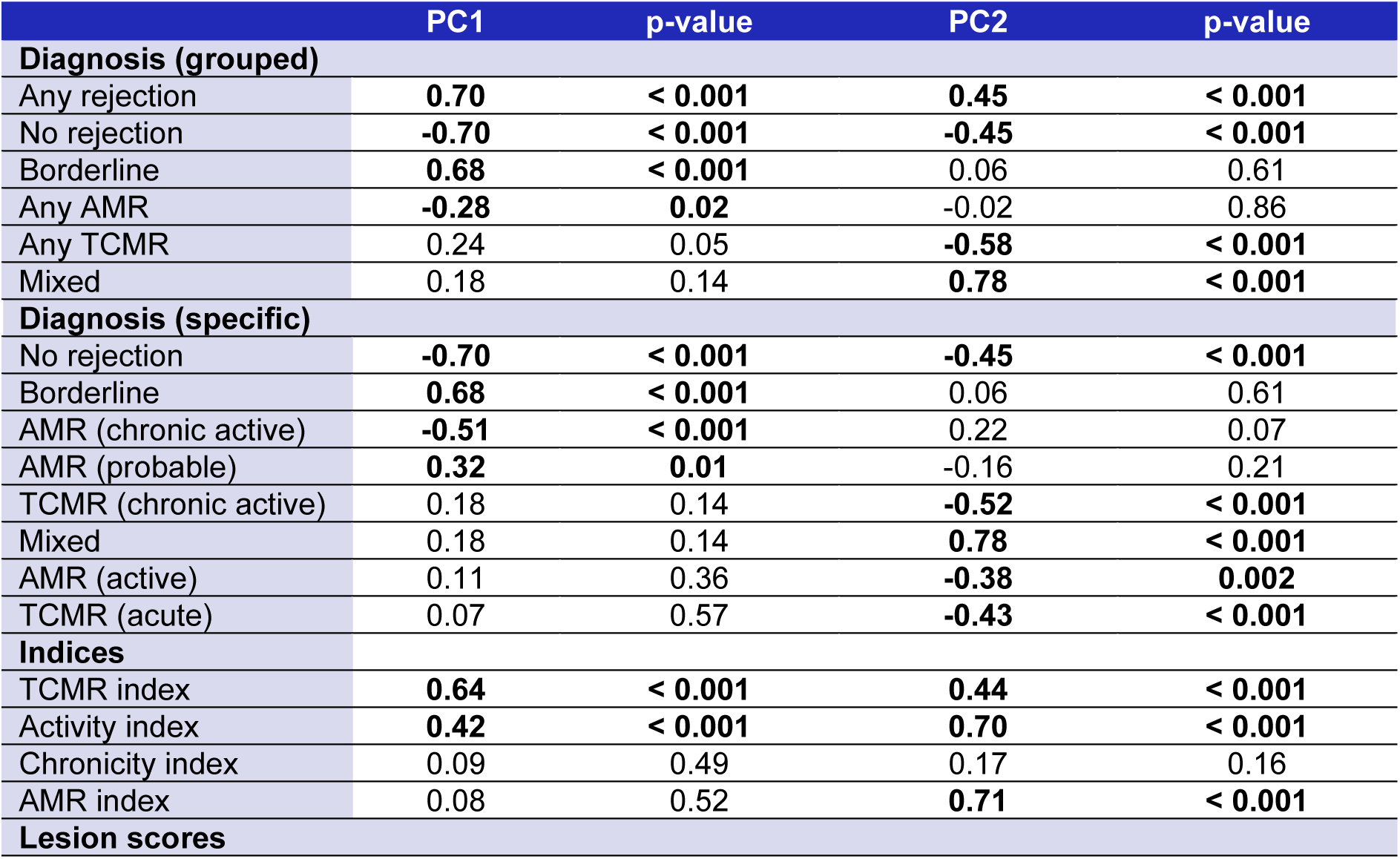
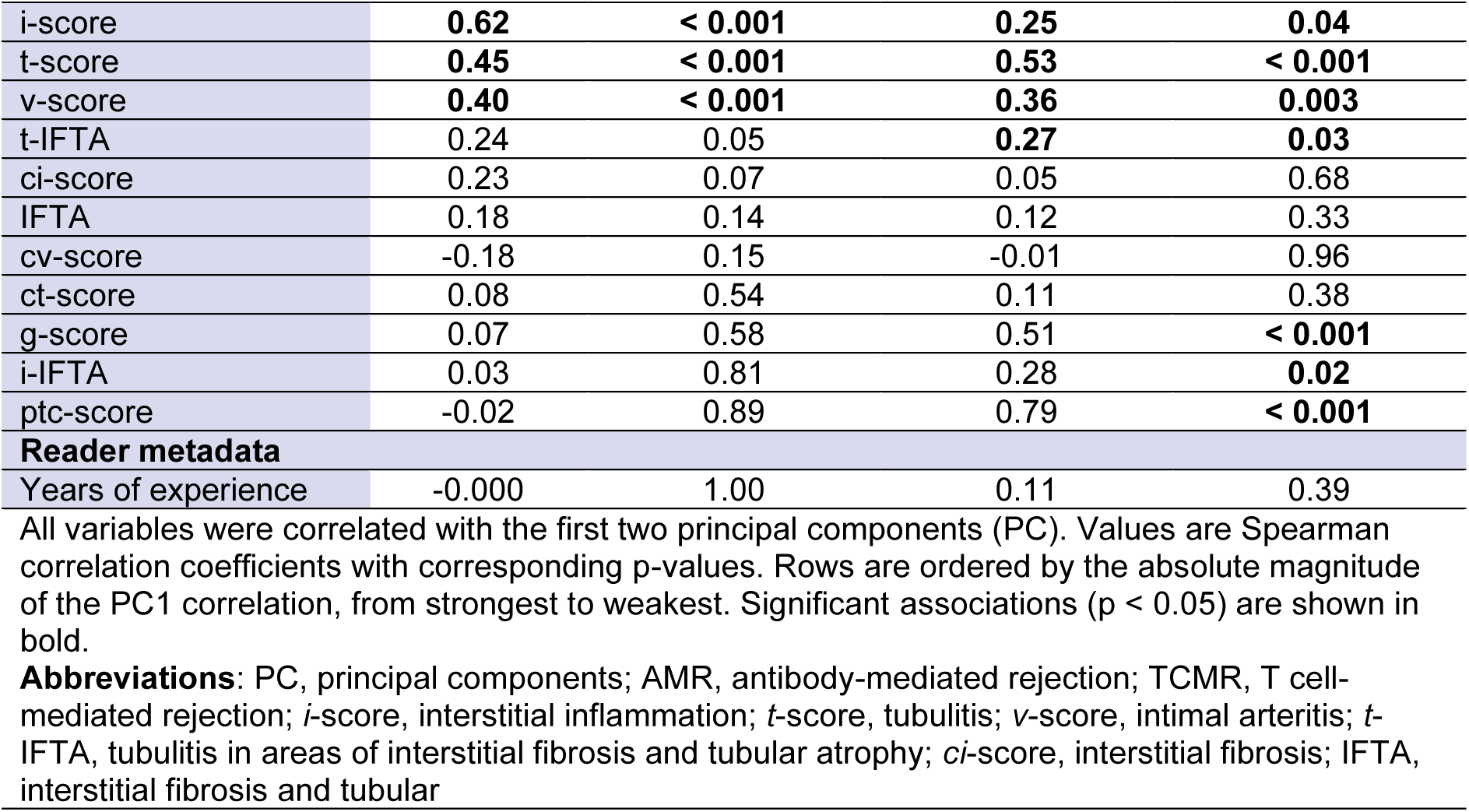
Correlations of reader-level variables diagnostic (dis)agreement principal components.

|  | PC1 | p-value | PC2 | p-value |
| --- | --- | --- | --- | --- |
| **Diagnosis (grouped)** |  |  |  |  |
| Any rejection | **0.70** | **< 0.001** | **0.45** | **< 0.001** |
| No rejection | **-0.70** | **< 0.001** | **-0.45** | **< 0.001** |
| Borderline | **0.68** | **< 0.001** | 0.06 | 0.61 |
| Any AMR | **-0.28** | **0.02** | -0.02 | 0.86 |
| Any TCMR | 0.24 | 0.05 | **-0.58** | **< 0.001** |
| Mixed | 0.18 | 0.14 | **0.78** | **< 0.001** |
| **Diagnosis (specific)** |  |  |  |  |
| No rejection | **-0.70** | **< 0.001** | **-0.45** | **< 0.001** |
| Borderline | **0.68** | **< 0.001** | 0.06 | 0.61 |
| AMR (chronic active) | **-0.51** | **< 0.001** | 0.22 | 0.07 |
| AMR (probable) | **0.32** | **0.01** | -0.16 | 0.21 |
| TCMR (chronic active) | 0.18 | 0.14 | **-0.52** | **< 0.001** |
| Mixed | 0.18 | 0.14 | **0.78** | **< 0.001** |
| AMR (active) | 0.11 | 0.36 | **-0.38** | **0.002** |
| TCMR (acute) | 0.07 | 0.57 | **-0.43** | **< 0.001** |
| **Indices** |  |  |  |  |
| TCMR index | **0.64** | **< 0.001** | **0.44** | **< 0.001** |
| Activity index | **0.42** | **< 0.001** | **0.70** | **< 0.001** |
| Chronicity index | 0.09 | 0.49 | 0.17 | 0.16 |
| AMR index | 0.08 | 0.52 | **0.71** | **< 0.001** |
| **Lesion scores** |  |  |  |  |
| i-score | **0.62** | **< 0.001** | **0.25** | **0.04** |
| t-score | **0.45** | **< 0.001** | **0.53** | **< 0.001** |
| v-score | **0.40** | **< 0.001** | **0.36** | **0.003** |
| t-IFTA | 0.24 | 0.05 | **0.27** | **0.03** |
| ci-score | 0.23 | 0.07 | 0.05 | 0.68 |
| IFTA | 0.18 | 0.14 | 0.12 | 0.33 |
| cv-score | -0.18 | 0.15 | -0.01 | 0.96 |
| ct-score | 0.08 | 0.54 | 0.11 | 0.38 |
| g-score | 0.07 | 0.58 | 0.51 | **< 0.001** |
| i-IFTA | 0.03 | 0.81 | 0.28 | **0.02** |
| ptc-score | -0.02 | 0.89 | 0.79 | **< 0.001** |
| **Reader metadata** |  |  |  |  |
| Years of experience | -0.000 | 1.00 | 0.11 | 0.39 |

All variables were correlated with the first two principal components (PC). Values are Spearman correlation coefficients with corresponding p-values. Rows are ordered by the absolute magnitude of the PC1 correlation, from strongest to weakest. Significant associations ( $p < 0.05$ ) are shown in bold.
**Abbreviations:** PC, principal components; AMR, antibody-mediated rejection; TCMR, T cell-mediated rejection; *i*-score, interstitial inflammation; *t*-score, tubulitis; *v*-score, intimal arteritis; *t*-IFTA, tubulitis in areas of interstitial fibrosis and tubular atrophy; *ci*-score, interstitial fibrosis; IFTA, interstitial fibrosis and tubular

### Molecular Annotation of the Principal Component Axes

To biologically annotate the major principal component axes of reader variation, reader-level molecular summaries were derived from the set of biopsies assigned to specific diagnostic categories by each reader. For PC1, summaries were defined as the mean feature value across biopsies each reader classified as rejection, excluding borderline classifications. Although higher PC1 values were associated with a greater reader-level tendency to assign rejection-related diagnoses, PC1 correlated inversely with multiple rejection-associated MMDx pathways among biopsies classified as rejection by each reader. The strongest inverse correlations were observed for QCAT, TCMR-RATs, NKAAG, Rej-RATs, RATs, AAG, QCMAT, TCB, and IIAAG (all false discovery rate (FDR)-adjusted p < 0.001) (**Figure 6A**, **Table 3**). CIBERSORTx showed corresponding inverse associations with immune-active cellular profiles, including plasma cells, CD8 T cells, CD4 T cells, NK cells, monocytes, and macrophages (all FDR-adjusted p < 0.001), whereas plasmacytoid dendritic cells showed the opposite pattern (ρ = 0.29, p = 0.02) (**Figure 6B**, **Table 4**). Together, these findings support the interpretation of PC1 as a rejection-calling threshold axis: readers with higher PC1 values classified a broader set of biopsies as rejection, including cases with comparatively lower molecular rejection activity. For PC2, reader-level molecular summaries were defined as the contrast between biopsies each reader classified as TCMR and those classified as AMR, excluding mixed, borderline, and no rejection classifications. PC2 was associated with multiple GSVA pathways linked to antibody-mediated or microvascular injury biology, including NK cells, B cells, monocytes/macrophages and CD4+ T cells, as well as ABMR-RATs, IIAAG, DSAST, GRIT1, AAG, GRIT3, NKAAG, and NKB pathogenesis-based transcript gene sets (FDR-adjusted p-values ranging from 0.01 to 0.04) (**Figure 6C-D**, **Table 3**).

**Figure 6.**
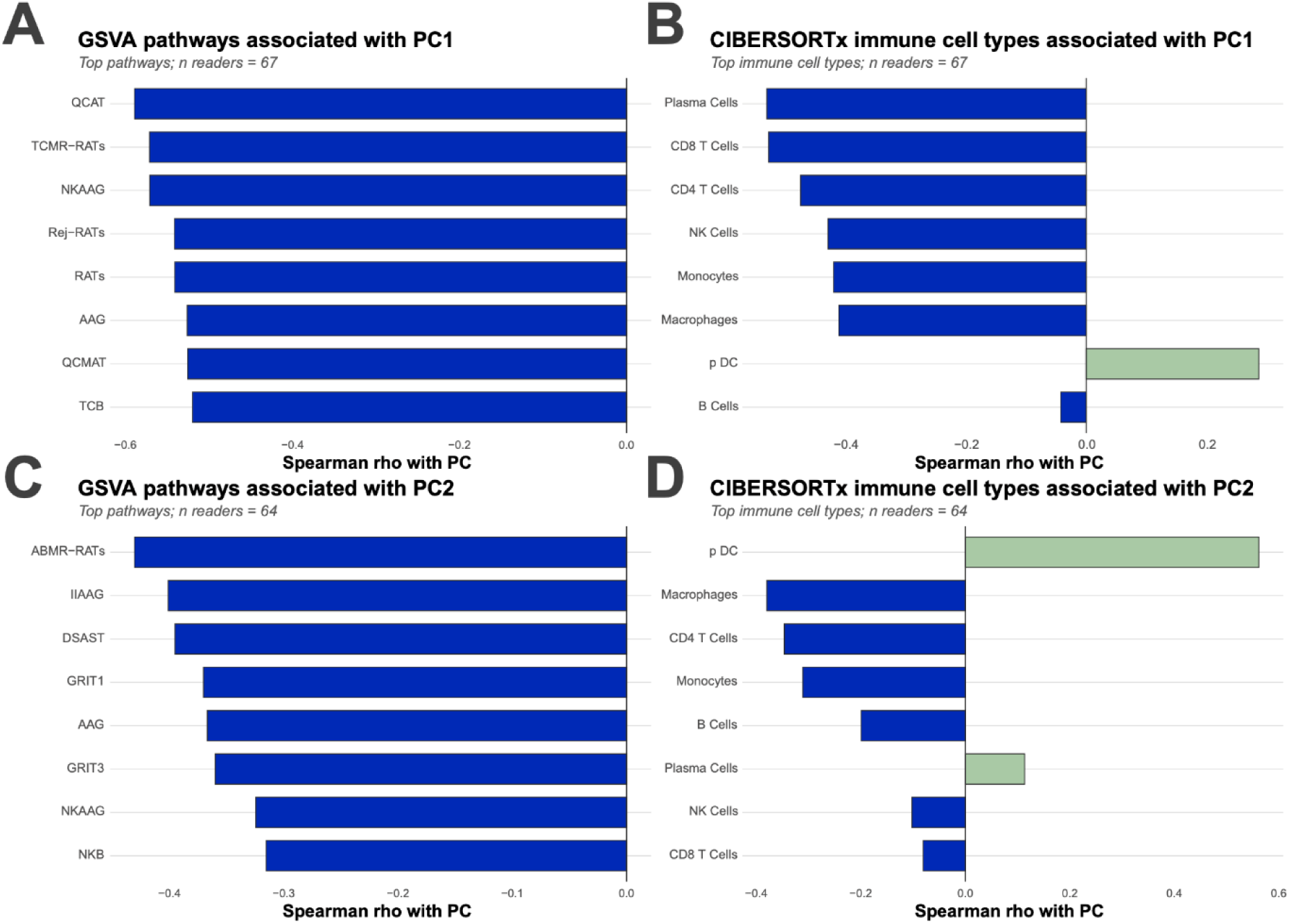
Molecular annotation of the principal axes of reader variation. Four-panel summary of the strongest molecular correlates of the first two principal components derived from the reader-by-reader pairwise Gwet’s AC1 matrix for final Banff diagnostic classification. **A.** Top GSVA pathways associated with PC1. **B.** Top CIBERSORTx-inferred immune cell types associated with PC1. **C.** Top GSVA pathways associated with PC2. **D.** Top CIBERSORTx-inferred immune cell types associated with PC2. Bars represent Spearman correlation coefficients between reader-level molecular summary features and the corresponding principal component. For PC1, molecular summaries were calculated across biopsies each reader classified as rejection; negative correlations indicate lower average rejection-associated molecular activity among biopsies classified as rejection by readers with higher PC1 values. This pattern supports interpretation of PC1 as a rejection-calling threshold axis, whereby higher-PC1 readers classified a broader set of biopsies as rejection. For PC2, molecular summaries were calculated as TCMR-minus-AMR contrasts; correlations therefore indicate molecular features associated with reader-level phenotypic orientation along the AMR/MVI–TCMR classification spectrum. Together, these findings support the biological interpretation of the major axes of reader variation.

**Table 3.**
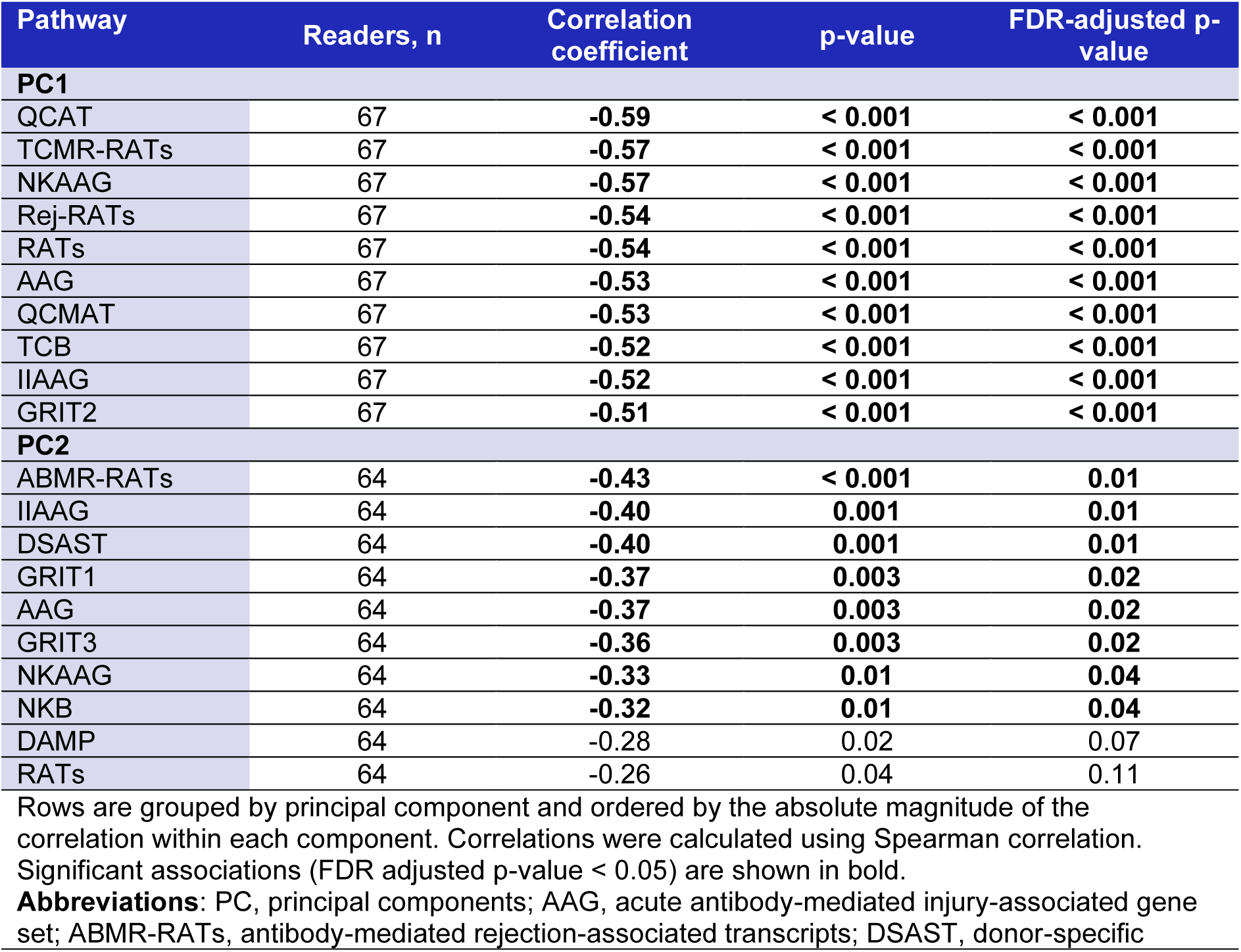
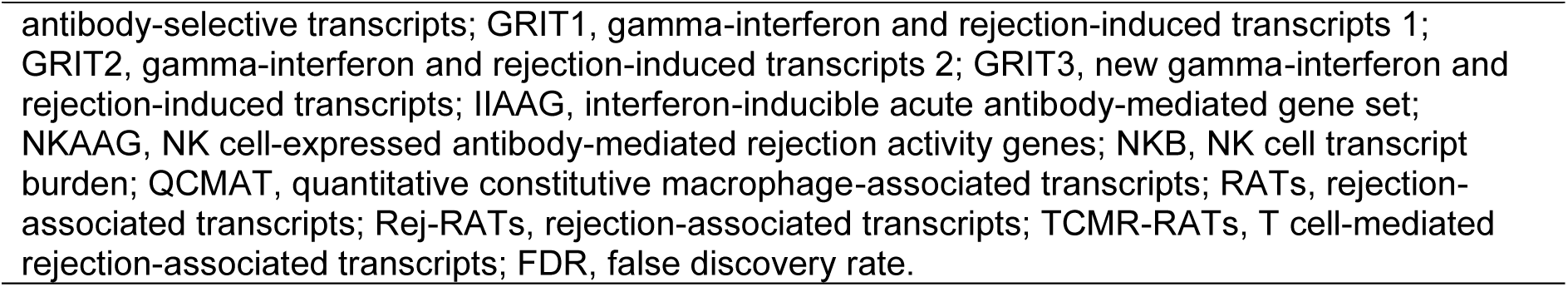
Top 10 GSVA pathway associated with PC1 and PC2.

**Table 4.**
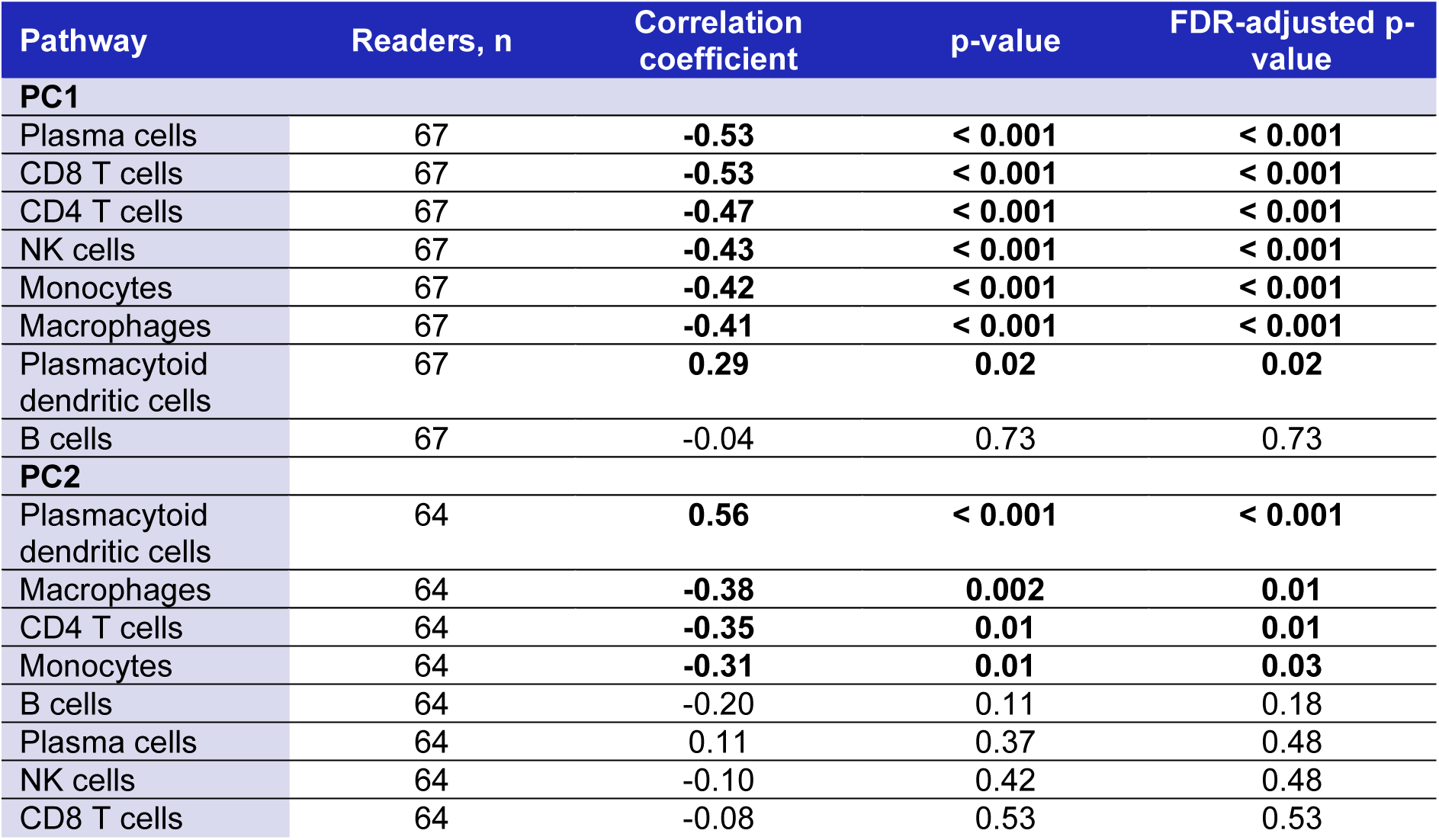
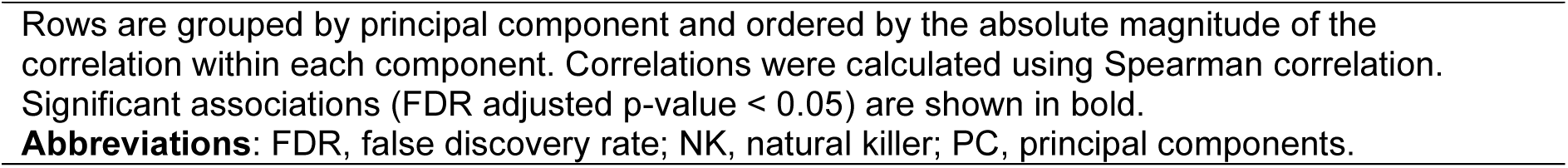
CIBERSORTx immune cell associated with PC1 and PC2.

CIBERSORTx showed a more selective immune-cell pattern, with plasmacytoid dendritic cells positively associated with PC2 (ρ = 0.56, p < 0.001), and macrophages (ρ = −0.38, p = 0.002), CD4 T cells (ρ = −0.35, p = 0.01), and monocytes (ρ = −0.31, p = 0.01) negatively associated (**Figure 5D**, **Table 4**). Together, these findings annotate PC2 as an TCMR-AMR/MVI-oriented phenotypic classification axis related to how readers resolve biopsies with overlapping tubulointerstitial and microvascular inflammatory features. Overall, these analyses suggest that the dominant axes of reader variation aligned with biologically coherent molecular programs, corroborating the explainability of diagnostic (dis)agreement by the indices and individual lesion scores. Full GSVA results are provided in **Supplementary Table S12**.

## DISCUSSION

In this international, fully digitized multi-reader study, interobserver variability in Banff classification was structured rather than random. Although overall agreement for final diagnostic categories was moderate, reproducibility varied substantially across biopsies, with disagreement concentrated in a subset of cases. Pairwise modeling showed that variability was hardly related to reader experience or geographic background and was instead driven primarily by differences in interpretation of specific Banff lesions along the thresholds/burden of inflammation, mainly active inflammatory and vascular lesions. Principal component analysis unified these findings by showing that reader variation was organized along two dominant dimensions corresponding to rejection thresholding, most prominently the cumulative burden of TCMR/TI activity (∼60% of diagnostic variation), and the cumulative burden of AMR/MVI activity (∼13% of variation). Minor components of pairwise diagnostic variation could be explained by combinations of individual lesions related to (chronic active) TCMR vs no rejection (∼4% of variation), and a more complex pattern of borderline TCMR and chronic active AMR vs chronic active TCMR and mixed rejection, mostly driven by intimal arteritis, peritubular capillaritis and tubulointerstitial inflammation in areas of fibrosis (t-IFTA and i-IFTA).

The dominant axis of variation reflected a reader-level threshold for assigning rejection-related diagnoses. PC1 was strongly associated with tubulointerstitial inflammatory lesions, including interstitial inflammation, tubulitis, and arteritis, as well as with TCMR-related indices and overall activity burden. Readers with higher PC1 values more readily classified biopsies as rejection, whereas readers with lower PC1 values appeared to require stronger inflammatory signals before doing so. This suggests that a substantial component of interobserver disagreement arises from differences in diagnostic thresholding and integration of active inflammatory lesions, rather than simply from inconsistent recognition of isolated lesions. This interpretation is supported by the pairwise models, in which arteritis, tubulitis, and interstitial inflammation were among the strongest lesion-level correlates of diagnostic disagreement. This largest variation is therefore driven by conservative versus permissive lesion calling not related to years of experience but likely related to pathologist characteristics (as has been described for e.g., PD-L1 scoring in lung cancer) or even the experience pathologists had in their particular health system (legally conservative and medically aggressive).^19^

The observation that this dominant axis was not explained by years of experience or geography suggests that rejection-calling thresholds may reflect more persistent reader-level interpretive tendencies. These tendencies may be influenced by local diagnostic culture, perceived clinical consequences of underdiagnosis versus overdiagnosis, or individual approaches to uncertainty, rather than by experience alone. Similar concepts have been described in other areas of diagnostic pathology. For example, in dermatopathology, malpractice concerns have been associated with *assurance behaviors* such as requesting additional stains, deeper sections, molecular tests, second opinions, or additional sampling, and such behaviors have been linked to diagnostic upgrading.^20^ Although comparable data are lacking in kidney (transplant) pathology, these observations provide a useful analogy for understanding how diagnostic thresholds may vary between pathologists. The present findings therefore suggest that reducing interobserver variability among expert renal pathologists may require more than additional instruction; it may also require clearer operationalization of lesion thresholds and diagnostic integration rules.

The second major axis captured phenotypic orientation among rejection-related classifications. PC2 was associated with peritubular capillaritis, glomerulitis, AMR-related indices, and mixed diagnoses, indicating that this dimension reflects how readers classify biopsies along the TCMR-AMR/MVI spectrum. Importantly, this axis did not primarily reflect whether rejection was recognized, but rather how its phenotype was resolved once injury was considered present. The close relationship with mixed diagnoses supports the interpretation that this source of variability is concentrated in biopsies with overlapping tubulointerstitial and microvascular inflammatory features, where readers may differ in how they weigh these components within the Banff framework.

The molecular analyses biologically annotated these two principal axes. For PC1, higher values were associated histologically with a greater tendency to assign rejection-related diagnoses, yet molecular summaries among biopsies classified as rejection showed inverse correlations with multiple rejection-associated transcript sets and immune-cell profiles. This pattern is consistent with the interpretation of PC1 as a rejection-calling threshold axis: readers with higher PC1 values classified a broader set of biopsies as rejection, including cases with comparatively lower molecular rejection activity. Thus, PC1 should not be interpreted as a marker of greater biological rejection severity, but as a reader-level tendency to apply a lower diagnostic threshold for rejection-related classification. For PC2, the strongest molecular associations involved pathways linked to antibody-mediated or microvascular injury biology and more selective immune-composition patterns, supporting its interpretation as an TCMR-AMR/MVI-oriented phenotypic classification axis. These findings do not imply that pathologists interpret molecular data directly. Rather, they indicate that the histologic decision patterns captured by PCA align with underlying biopsy biology and reflect systematic differences in how pathologists classify biopsies with shared but variably expressed inflammatory and microvascular injury signals.

Taken together, these findings support a conceptual model in which interobserver variability in Banff classification arises from two largely distinct processes: a rejection-calling threshold axis governing how readily rejection-related diagnoses are assigned, driven mainly by tubulointerstitial inflammatory lesions, and a phenotypic classification axis governing whether inflammatory injury is resolved in a more TCMR-oriented, AMR/MVI-oriented, or mixed-rejection direction. Notably, these dimensions closely parallel the biological compartments emphasized within the Banff classification itself. Variability therefore appears to arise less from arbitrary reader behavior than from differences in how biologically meaningful lesion groups are weighted and integrated in complex biopsies.

These findings have potential implications for future refinement of Banff-based assessment. The identification of a rejection-calling threshold axis suggests that variability may arise in part from differences in how minimum inflammatory requirements for rejection are operationalized, particularly for interstitial inflammation, tubulitis, and arteritis. Similarly, the presence of a TCMR-AMR/MVI-oriented phenotypic classification axis highlights continued ambiguity in the interpretation and integration of microvascular lesions, especially in biopsies with overlapping features. A clearer and more standardized approach to weighing lesion compartments may therefore improve consistency, particularly in diagnostically intermediate or mixed-pattern cases. More broadly, framing diagnostic variability within a two-axis biological model may help explain patterns of disagreement reported in prior transplant pathology studies and may provide a useful basis for future quantitative or decision-support approaches. This study also provides valuable information for the development of machine learning-derived decision support algorithms. Artificial intelligence could help reduce interobserver variability and allow for more uniform use of diagnostic thresholds in the future.

Several limitations should be acknowledged. First, the number of biopsy cases was limited, and cases were selected to represent a diagnostically informative spectrum rather than population prevalence, although each case was evaluated by a large number of independent readers. Second, participants were recruited internationally and represented a heterogeneous group of renal pathologists; while this may introduce additional variability, it also increases relevance to real-world practice. Third, readers were blinded to clinical information, whereas Banff interpretation in clinical practice is likely influenced by clinical context, including donor-specific antibody status, graft function, timing after transplantation, and prior biopsy findings. It could be imagined that more experienced renal pathologists are more capable to perform clinicopathological integration and it would therefore be interesting to perform a reader study with and without clinical information available in the future. Fourth, the study was designed to assess reproducibility rather than diagnostic accuracy, and no external reference standard was applied. Fifth, in multicenter reader studies, readers by definition assess stainings that are not from their own center, which might further increase variation. Finally, because diagnostic categories are derived from lesion scores, the observed relationships between lesion-level differences and diagnostic disagreement reflect how readers weigh and integrate lesions rather than independent causal effects of individual lesions. Considering these limitations, we have to acknowledge that any reader study tries to approach in some ways how pathologists assess their biopsies, but it is never a true reflection of clinical practice. Adequate clinical care therefore requires constant communication between the pathologists and the local transplant physicians, for instance in the setting of multidisciplinary meetings.^21^

In conclusion, interobserver variability in Banff kidney transplant biopsy interpretation is organized along two biologically meaningful dimensions: a rejection-calling threshold axis linked primarily to tubulointerstitial inflammation, and a phenotypic classification axis linked to TCMR-AMR/MVI-oriented and mixed-rejection interpretation. Diagnostic disagreement is concentrated in biopsies with intermediate or overlapping inflammatory rather than chronic features, where readers differ in how they threshold and integrate key lesion compartments. Improving reproducibility will therefore likely depend more on clearer standardization of lesion interpretation and diagnostic integration than on reader experience alone.

## Data Availability

All data produced in the present study are available upon reasonable request to the authors

## AUTHORSHIP STATEMENTS

RHB, RdL and JK were involved in the study design. Histological data collection was performed by JK. Scoring was performed by all 67 pathologists. Molecular data was generated by NK, IBB, GC, BvdW and JK. Clinical data was curated by DvdH and APJdV. Data analysis was performed by NK, RdL, RHB and JK. Original draft of the manuscript was written by RHB and RdL. Critical input to the analyses was given by GB and HPS. The final version of the manuscript was revised and approved by all authors.

## ABBREVIATIONS

AAG: Acute antibody-mediated injury associated gene set
ABMR-RATs: ABMR-associated rat transcripts
AC1: Gwet’s agreement coefficient 1
AC2: Gwet’s agreement coefficient 2
AEG: ABMR-associated endothelial genes
ah: Arteriolar hyalinosis
AI: Activity index
AIC: Akaike’s information criterion
AMAT1: Alternative Macrophage Activation Transcripts 1
AMR: Antibody-mediated rejection
ANOVA: Analysis of variance
BAT: B cell-associated transcripts
BKPyVN: BK virus-associated nephropathy
cDC1: Conventional dendritic cell 1
cDC2: Conventional dendritic cell 2
C4d: Complement split product C4d
cg: Glomerular basement membrane double contours
CI: Confidence interval
CIBERSORTx: Cell-type Identification by Estimating Relative Subsets of RNA Transcripts X
ci: Interstitial fibrosis
ComBat-seq: Batch-effect correction method for RNA sequencing count data
CPM: Counts per million
ct: Tubular atrophy
cv: Vascular fibrous intimal thickening
DAMP: Damage-associated molecular pattern transcripts
DBD: Donation after brain death
DCD: Donation after cardiac death
DCT: Distal convoluted tubule cells
DGF: Delayed graft function
DSA: Donor-specific antibody
DSA10: Top 10 transcripts associated with donor-specific antibodies
DSAST: Donor-specific antibody-selective transcripts
ECg: Endothelial glomerular cells
ECptc: Endothelial cells, peritubular capillary
ECvr: Endothelial cells, vasa recta
eDSAST: Endothelial donor-specific antibody-selective transcripts
ENDAT: Endothelial-associated transcripts
FDR: False discovery rate
FFPE: Formalin-fixed paraffin-embedded
FICOL: Fibrillar collagen transcripts
g: Glomerulitis
GRIT1: Gamma-IFN and rejection-induced transcripts 1
GRIT2: Gamma-IFN and rejection-induced transcripts 2
GRIT3: New gamma-IFN and rejection-induced transcripts
GSVA: Gene set variation analysis
Gwet AC: Gwet’s agreement coefficient
H&E: Hematoxylin and eosin
HLA: Human leukocyte antigen
i-IFTA: Inflammation in areas of interstitial fibrosis and tubular atrophy
i: Interstitial inflammation
IC A/B: Intercalated cells type A and B
ICC: Intraclass correlation coefficient
IFN: Interferon
IFTA: Interstitial fibrosis and tubular atrophy
IGF: Immediate graft function
IIAAG: IFN-inducible acute antibody-mediated gene set
IQR: Interquartile range
IRITD1: Injury and repair-induced transcripts – early
IRITD3: Injury and repair-induced transcripts – intermediate
IRITD5: Injury and repair-induced transcripts – late
IRRAT: Injury-repair response-associated transcripts
KT1: Kidney transcripts – set 1
KT2: Kidney transcripts – set 2
LOH: Loop of Henle
MCAT: Mast cell-associated transcript
mm: Mesangial matrix increase
MMDx: Molecular Microscope® Diagnostic System
MVI: Microvascular inflammation
NK cell: Natural killer cell
NKAAG: NK cell-expressed ABMR activity genes
OR: Odds ratio
PAS: Periodic acid–Schiff
PC: Principal component
PCA: Principal component analysis
pDC: Plasmacytoid dendritic cell
PD-L1: Programmed death-ligand 1
PE: Pelvic epithelium
PEC: Parietal epithelial cells
pmp: Per million population
PNF: Primary non-function
PT: Proximal tubule
PT injured: Proximal tubule (injured)
ptc: Peritubular capillaritis
QCAT: Quantitative CTL-associated transcripts
QCMAT: Quantitative constitutive macrophage-associated transcripts
RNA: Ribonucleic acid
SD: Standard deviation
TCMR: T cell-mediated rejection
TCMR-RATs: TCMR-associated rat transcripts
TempO-Seq: Templated Oligo-Sequencing
t-IFTA: Tubulitis in areas of interstitial fibrosis and tubular atrophy
ti: Total inflammation
TI index: Total inflammation index
t: Tubulitis
UMAP: Uniform manifold approximation and projection
v: Intimal arteritis
vSMp: Vascular smooth muscle cells and pericytes

## REFERENCES

1. Naesens M, Roufosse C, Haas M, et al. The Banff 2022 Kidney Meeting Report: Reappraisal of microvascular inflammation and the role of biopsy-based transcript diagnostics. Am J Transplant. Mar 2024;24(3):338–349. doi:10.1016/j.ajt.2023.10.016

2. Loupy A, Mengel M, Haas M. Thirty years of the International Banff Classification for Allograft Pathology: the past, present, and future of kidney transplant diagnostics. Kidney Int. Apr 2022;101(4):678–691. doi:10.1016/j.kint.2021.11.028

3. Furness PN, Taub N, Assmann KJ, et al. International variation in histologic grading is large, and persistent feedback does not improve reproducibility. Am J Surg Pathol. Jun 2003;27(6):805–10. doi:10.1097/00000478-200306000-00012

4. Marcussen N, Olsen TS, Benediktsson H, Racusen L, Solez K. Reproducibility of the Banff classification of renal allograft pathology. Inter- and intraobserver variation. Transplantation. Nov 27 1995;60(10):1083–9. doi:10.1097/00007890-199511270-00004

5. Veronese FV, Manfro RC, Roman FR, et al. Reproducibility of the Banff classification in subclinical kidney transplant rejection. Clin Transplant. Aug 2005;19(4):518–21. doi:10.1111/j.1399-0012.2005.00377.x

6. Huang SC, Lin YJ, Wen MC, et al. Unsatisfactory reproducibility of interstitial inflammation scoring in allograft kidney biopsy. Sci Rep. May 1 2023;13(1):7095. doi:10.1038/s41598-023-33908-3

7. Liang PI, Lin WC, Wen MC, et al. Learning more from the inter-rater reliability of interstitial fibrosis assessment beyond just a statistic. Sci Rep. Aug 15 2023;13(1):13260. doi:10.1038/s41598-023-40221-6

8. Dasari S, Chakraborty A, Truong L, Mohan C. A Systematic Review of Interpathologist Agreement in Histologic Classification of Lupus Nephritis. Kidney Int Rep. Oct 2019;4(10):1420–1425. doi:10.1016/j.ekir.2019.06.011

9. Schinstock CA, Sapir-Pichhadze R, Naesens M, et al. Banff survey on antibody-mediated rejection clinical practices in kidney transplantation: Diagnostic misinterpretation has potential therapeutic implications. Am J Transplant. Jan 2019;19(1):123–131. doi:10.1111/ajt.14979

10. Smith B, Cornell LD, Smith M, et al. A method to reduce variability in scoring antibody-mediated rejection in renal allografts: implications for clinical trials - a retrospective study. Transpl Int. Feb 2019;32(2):173–183. doi:10.1111/tri.13340

11. Mengel M, Chan S, Climenhaga J, et al. Banff initiative for quality assurance in transplantation (BIFQUIT): reproducibility of C4d immunohistochemistry in kidney allografts. Am J Transplant. May 2013;13(5):1235–45. doi:10.1111/ajt.12193

12. Gwet KL. Computing inter-rater reliability and its variance in the presence of high agreement. Br J Math Stat Psychol. May 2008;61(Pt 1):29–48. doi:10.1348/000711006x126600

13. Naesens M, Roufosse C, Cornell LD, et al. The Banff 2024 Kidney Meeting Report: Rejection as a spectrum of phenotypes and focus on differential diagnostic reasoning. Am J Transplant. May 2026;26(5):922–936. doi:10.1016/j.ajt.2026.01.018

14. Yoo D, Goutaudier V, Divard G, et al. An automated histological classification system for precision diagnostics of kidney allografts. Nat Med. May 2023;29(5):1211–1220. doi:10.1038/s41591-023-02323-6

15. Vaulet T, Divard G, Thaunat O, et al. Data-Driven Chronic Allograft Phenotypes: A Novel and Validated Complement for Histologic Assessment of Kidney Transplant Biopsies. J Am Soc Nephrol. Nov 2022;33(11):2026–2039. doi:10.1681/asn.2022030290

16. Vaulet T, Divard G, Thaunat O, et al. Data-driven Derivation and Validation of Novel Phenotypes for Acute Kidney Transplant Rejection using Semi-supervised Clustering. J Am Soc Nephrol. May 3 2021;32(5):1084–1096. doi:10.1681/asn.2020101418

17. Vaulet T, Koshy P, Wellekens K, et al. Continuous indices to assess the phenotypic spectrum of kidney transplant rejection. Nat Commun. Nov 26 2025;16(1):10417. doi:10.1038/s41467-025-65153-9

18. Newman AM, Steen CB, Liu CL, et al. Determining cell type abundance and expression from bulk tissues with digital cytometry. Nat Biotechnol. Jul 2019;37(7):773–782. doi:10.1038/s41587-019-0114-2

19. Butter R, Hondelink LM, van Elswijk L, et al. The impact of a pathologist’s personality on the interobserver variability and diagnostic accuracy of predictive PD-L1 immunohistochemistry in lung cancer. Lung Cancer. Apr 2022;166:143–149. doi:10.1016/j.lungcan.2022.03.002

20. Titus LJ, Reisch LM, Tosteson ANA, et al. Malpractice Concerns, Defensive Medicine, and the Histopathology Diagnosis of Melanocytic Skin Lesions. Am J Clin Pathol. Aug 30 2018;150(4):338–345. doi:10.1093/ajcp/aqy057

21. Sapir-Pichhadze R, Askar M, Cooper M, et al. Rethinking the Diagnosis and Management of Antibody-Mediated Rejection in Multidisciplinary Transplant Meetings: A Global Survey and Banff Working Group Recommendations. Clin Transplant. May 2025;39(5):e70167. doi:10.1111/ctr.70167

