## Supplementary Material for "Drivers of Diagnostic Variation in a Digital Global Kidney Transplant Reader Study"

### Supplemental methods

#### Data Preparation and Dataset Construction

Three primary reader-level data sources were integrated for analysis: Banff lesion scoring data, reader experience metadata, and reader geographic metadata. Biopsy-level metadata, including donor-specific antibody status, C4d status, and molecular data availability, were linked where required for diagnostic rule assignment and molecular annotation analyses. Data were imported into R using the **readxl** package, and variable names were standardized using the **janitor** package. Reader and biopsy identifiers were converted to character variables before merging to ensure consistent linkage across datasets and to avoid unintended numeric recoding. The primary analytical dataset was structured in long format, with each row representing one reader evaluation of one biopsy. Variables included biopsy identifier, reader identifier, Banff lesion scores, non-assessable lesion indicators, derived diagnostic indicator variables, final diagnostic category, composite histological indices, and reader-level characteristics, including years of renal pathology experience, country of practice, and continent. Data integrity checks were performed before analysis. These included verification that each reader-biopsy combination occurred only once, that diagnostic category assignments were complete for analyses requiring final diagnosis, that categorical variables were encoded using prespecified factor levels, and that reader and biopsy identifiers were consistently linked across all analysis tables. Missing or non-assessable lesion scores were retained as explicit missing values and excluded only from analyses requiring the corresponding lesion or derived variable.

**Derivation of Final Diagnostic Categories**

Final diagnostic categories were derived from reader-level Banff lesion scores and biopsy-level donor-specific antibody and C4d status using rule-based diagnostic indicator variables. These indicator variables encoded whether each reader-biopsy evaluation fulfilled criteria for mixed rejection, antibody-mediated rejection, T cell-mediated rejection, borderline changes, or no rejection. Because more than one diagnostic indicator could be positive for a given reader-biopsy evaluation, diagnostic indicators were consolidated into a single mutually exclusive final diagnostic category for agreement analyses. A hierarchical classification scheme was applied to resolve overlapping diagnostic flags. Mixed rejection was assigned first when criteria for both antibody-mediated and T cell-mediated rejection were fulfilled. If mixed rejection was not present, antibody-mediated rejection categories were assigned in the following order: chronic active AMR, active AMR, chronic AMR, and probable AMR. If no antibody-mediated rejection category was assigned, T cell-mediated rejection categories were assigned in the following order: chronic active TCMR and acute TCMR. Acute TCMR categories 1A, 1B, 2A, 2B, and 3 were collapsed into a single acute TCMR category for the primary diagnostic agreement analyses. Chronic active TCMR categories 1A, 1B, and 2 were collapsed into a single chronic active TCMR category. Borderline changes were assigned when the borderline indicator was positive and no higher-priority rejection category was present. Evaluations that did not meet criteria for any rejection or borderline category were assigned to no rejection. The resulting final diagnostic variable comprised nine mutually exclusive categories: no rejection, borderline changes, acute TCMR, chronic active TCMR, probable AMR, active AMR, chronic AMR, chronic active AMR, and mixed rejection. These categories were encoded using fixed prespecified factor levels to ensure stable ordering across agreement analyses, bootstrap resampling, diagnostic distribution summaries, and visualization. Quality-control checks confirmed that each reader-biopsy evaluation yielded a non-missing final diagnostic category after harmonization.

**Construction of Reader-Case Analysis Matrices**

For agreement analyses of final diagnostic categories, the long-format reader-biopsy dataset was reshaped into case-by-reader rating matrices. Rows corresponded to biopsy cases and columns corresponded to readers. Each matrix entry contained the final diagnostic category assigned to a given biopsy by a given reader after rule-based diagnostic harmonization. Diagnostic categories were encoded using fixed prespecified category levels to ensure stable ordering across agreement estimation, bootstrap resampling, diagnostic distribution summaries, heatmap visualization, and principal component analysis. The same case-by-reader structure was used to generate the reader-by-reader diagnostic similarity matrix. For each pair of readers, diagnostic agreement across the shared biopsy set was quantified using Gwet's AC1. These pairwise agreement estimates formed a symmetric reader similarity matrix that was used for structural analyses of reader interpretation patterns. Lesion-level datasets and reader-pair datasets were generated separately for analyses requiring Banff lesion-score differences between readers, as described below.

**Interobserver Agreement Analysis**

Interobserver agreement for final Banff diagnostic categories was assessed using Gwet's agreement coefficient for nominal data (AC1). AC1 was applied to the case-by-reader diagnostic rating matrix, in which each biopsy case constituted the unit of observation and each reader contributed one categorical final diagnosis per case. Diagnostic categories were encoded using fixed prespecified category levels before agreement estimation to ensure consistent interpretation across pooled and resampled analyses. Agreement for individual Banff lesion scores was quantified using Gwet's agreement coefficient for ordinal data (AC2) with linear weighting. Linear weighting was used because Banff lesion scores are ordinal and increasing distance between score categories reflects increasing disagreement. Lesion-level agreement analyses were performed separately for each Banff lesion using all reader-case observations with an evaluable score for the lesion under consideration. Non-assessable lesion entries were retained as missing values and excluded only from analyses requiring that specific lesion score. For both diagnostic-category and lesion-level agreement analyses, observed agreement proportions were calculated descriptively to provide context for chance-corrected agreement coefficients. AC estimates, standard errors, and 95% confidence intervals were calculated using the **irrCAC** package in R. Lesions or diagnostic categories with insufficient variability for stable coefficient estimation were excluded from coefficient-based analyses but retained in descriptive summaries where applicable.

#### Assessment of Case-level Diagnostic Concordance

Case-level diagnostic concordance was calculated to quantify heterogeneity in diagnostic agreement across biopsy cases. For each biopsy, the distribution of final diagnostic categories assigned by all eligible readers was tabulated. The majority diagnosis was defined as the most frequently assigned final diagnostic category for that biopsy, and case-level concordance was calculated as the number of readers assigning the majority diagnosis divided by the total number of readers with a non-missing final diagnostic category for that biopsy. This metric ranges from 0 to 1, with higher values indicating stronger reader consensus and lower values indicating greater dispersion of diagnostic interpretation. When multiple diagnostic categories had the same maximum frequency, the shared maximum proportion was used as the concordance value. Case-level concordance was visualized using ordered bar plots, with biopsies sorted by majority-diagnosis proportion. Reader-by-case heatmaps were generated using the same biopsy order to visualize the distribution of diagnostic assignments across readers and to identify cases with concentrated versus heterogeneous disagreement patterns.

**Agreement Analysis of Composite Histological Indices**

Agreement for the composite histological indices was evaluated separately from individual Banff lesion scores because the Activity Index, Chronicity Index, TCMR/TI index, and AMR/MVI index are numerical composite measures derived from multiple Banff lesion components. For each reader-biopsy evaluation, index values were calculated from the corresponding reader-level lesion scores according to the published index definitions. Index values were treated as composite numerical outcomes for agreement analysis.

The Activity Index was calculated as:

$$\text{Activity Index} = t + i + v + g + ptc + 2 \times \text{C4d}$$

where *t* denotes tubulitis, *i* interstitial inflammation, *v* intimal arteritis, *g* glomerulitis, and *ptc* peritubular capillaritis. C4d was encoded as a binary variable, with 0 indicating negative C4d staining and 1 indicating positive C4d staining according to Banff definitions. The resulting Activity Index ranges from 0 to 17.

The Chronicity Index was calculated as:

Chronicity Index = $ci + ct + cv + 2 \times cg$

where *ci* denotes interstitial fibrosis, *ct* tubular atrophy, *cv* vascular fibrous intimal thickening, and *cg* glomerular basement membrane double contours/transplant glomerulopathy. The resulting Chronicity Index ranges from 0 to 15.

The AMR/MVI index was calculated as:

AMR/MVI Index = $0.938 \times g + 0.762 \times ptc + 0.728 \times cg + 2.716 \times \mathrm{C4d}$

This index was used as a continuous measure of the antibody-mediated rejection/microvascular inflammation spectrum. C4d was encoded as a binary variable as described above. The AMR/MVI index ranges from 0 to 10.

The TCMR/TI index was calculated as:

TCMR/TI Index = $0.970 \times i + 0.623 \times t + 1.540 \times v + 0.195 \times ct$

This index was used as a continuous measure of the T cell-mediated rejection/tubulointerstitial inflammation spectrum. The TCMR/TI index ranges from 0 to 10. Because some index components were unavailable when required biopsy structures were non-assessable (arteries), variance components were estimated using linear mixed-effects models rather than standard ANOVA-based intraclass correlation coefficient estimation with listwise deletion. Biopsy case and reader were modeled as random effects. This approach allowed estimation of case-level variance, reader-level variance, and residual variance while retaining all available reader-biopsy observations for each index. Two intraclass correlation coefficients were calculated for each composite index. Absolute-agreement ICC was defined as:

ICC(2,1) = Var(Case) / [Var(Case) + Var(Reader) + Var(Residual)]

This measure reflects agreement in absolute index values and penalizes systematic differences in reader scoring thresholds. Consistency ICC was defined as:

ICC(3,1) = Var(Case) / [Var(Case) + Var(Residual)]

This measure evaluates whether readers rank biopsies similarly while ignoring systematic reader-level differences in baseline scoring tendency. A higher consistency ICC than absolute-agreement ICC was interpreted as evidence that readers preserved relative case ranking more strongly than absolute score calibration, consistent with systematic reader leniency or severity in composite index scoring. Uncertainty around ICC estimates was quantified using parametric bootstrapping. Parametric bootstrap confidence intervals for ICC estimates were derived using 200 simulations per index. For each index, 95% confidence intervals were derived from bootstrap distributions of the corresponding variance-component estimates. In addition, pairwise reader-bias analyses were performed by calculating mean between-reader differences in index values across the shared evaluable biopsy subset for each reader pair. This analysis was used to identify reader pairings contributing most strongly to absolute index disagreement.

**Bootstrap Subgroup Analysis by Reader Experience and Geography**

Subgroup analyses were performed to assess whether diagnostic agreement varied according to reader experience or geographic region. Reader experience was categorized into three prespecified strata: <5 years, 5–10 years, and >10 years of renal pathology experience. Geographic analyses were performed at the continent level using United Nations geographic classifications. Subgroup analyses were based on final Banff diagnostic categories and used Gwet's AC1 as the agreement metric. To account for unequal subgroup sizes and to estimate uncertainty around subgroup-level agreement estimates, case-level bootstrap resampling was performed. In each bootstrap replicate, biopsy cases were sampled with replacement, while preserving all reader evaluations within sampled cases. Gwet's AC1 was recalculated within each eligible subgroup for each bootstrap sample, generating empirical distributions of subgroup-specific agreement estimates. Pairwise differences in agreement between subgroups were calculated within each bootstrap replicate, and 95% confidence intervals for subgroup differences were derived from the percentile distribution of the bootstrap estimates. For experience-based analyses, all prespecified experience strata were evaluated. For continent-level analyses, groups were included in pairwise bootstrap comparisons only when they met the predefined minimum reader threshold. Experience strata were included when at least 3 readers were available. Continent-level groups were included in pairwise bootstrap comparisons when at least 5 readers were available. The primary subgroup bootstrap analyses used 500 bootstrap replicates with a fixed random seed of 123. A balanced-bootstrap sensitivity analysis was additionally performed for continent-level comparisons by sampling an equal number of readers per continent in each bootstrap replicate. For the balanced continent-level sensitivity analysis, 3 readers per continent were sampled in each bootstrap replicate. These subgroup analyses were considered supportive and exploratory.

### Pairwise Modeling of Diagnostic Agreement

To identify determinants of diagnostic agreement, the long-format reader-biopsy dataset was transformed into a pairwise reader-comparison structure. For each biopsy, all unique reader pairs evaluating that biopsy were generated, with each observation representing one reader pair for one biopsy. Reader pairs were restricted to unique unordered combinations, so that each pair contributed once per biopsy. Diagnostic agreement was encoded as a binary outcome indicating whether the two readers assigned the same final Banff diagnostic category to the biopsy. Reader-level covariates included the absolute difference in years of renal pathology experience and geographic similarity. Geographic similarity was defined as practice on the same continent in the primary model. The same-continent variable was encoded as a binary indicator. Mixed-effects logistic regression models were fitted with diagnostic agreement as the dependent variable and biopsy case included as a random intercept to account for clustering of reader-pair observations within biopsies and heterogeneity in intrinsic case difficulty. To quantify the contribution of lesion-level interpretive differences, absolute between-reader differences in Banff lesion scores were calculated for each evaluable lesion. Lesion-difference variables represented the absolute score distance between two readers for the same biopsy. Ordinal lesion scores were treated numerically for this purpose, so that a 1-unit increase represented one additional category of between-reader difference. Non-assessable or missing lesion scores were excluded from models requiring the corresponding lesion-difference variable. Three model specifications were evaluated. The base model included only reader-level covariates: absolute difference in years of experience and same-continent practice. The full lesion model included these reader-level covariates plus absolute score differences for all evaluated Banff lesions: *g*, *cg*, *mm*, *i*, *t*, *ti*, IFTA, *ci*, *ct*, *i*-IFTA, *t*-IFTA, *ptc*, *v*, *cv*, and *ah*. The core lesion model included a focused subset of diagnostically central lesions selected as a reduced biologically motivated model: v, ptc, i, t, ci, and ct. Correlations among lesion-difference variables were examined descriptively to evaluate interdependence among scoring-disagreement patterns. Model fit was compared using Akaike's information criterion. Regression coefficients were exponentiated and reported as odds ratios with 95% confidence intervals. Odds ratios below 1 indicate lower odds of diagnostic agreement with increasing between-reader difference in the corresponding lesion score or reader-level covariate. Models were fitted in R using the **lme4** package, and model estimates were summarized using **broom.mixed**.

**Principal Component Analysis of Reader Similarity**

To characterize global patterns of diagnostic similarity among readers, pairwise inter-reader agreement was calculated using Gwet's AC1 across all shared biopsy cases. These pairwise agreement estimates were assembled into a symmetric reader-by-reader diagnostic similarity matrix. Principal component analysis was applied to this matrix to identify dominant dimensions of variation in reader interpretation. The proportion of variance explained by each component was derived from the corresponding eigenvalues, and the first two components were used for primary visualization and downstream interpretive analyses. Component interpretation was performed post hoc. Reader coordinates along each principal component were correlated with reader-level summary metrics derived from the diagnostic and histological datasets, including proportions of final diagnostic categories, grouped rejection indices, composite histological indices, and mean Banff lesion scores. These correlations were used to assign descriptive biological and diagnostic interpretations to the leading reader axes. The component labels therefore reflect data-derived interpretations of reader behavior rather than prespecified clinical categories. Associations between principal component coordinates and continuous or ordinal reader-level variables were evaluated using Spearman correlation. Associations between principal component coordinates and categorical reader characteristics, including geographic region and experience category, were evaluated using Kruskal-Wallis testing. Detailed definitions of the reader-level molecular summaries used to annotate the interpreted principal components are provided in the molecular annotation section below.

**TempO-Seq Whole-Transcriptomic Profiling of FFPE Kidney Allograft Biopsies**

To enable high-throughput transcriptomic profiling of limited kidney allograft biopsy material, we used Templated Oligo-Sequencing (TempO-Seq). In contrast to conventional whole-transcriptome RNA sequencing workflows, which generally require RNA extraction, cDNA synthesis, and library preparation from fragmented RNA, TempO-Seq is based on target-specific detector oligonucleotide pairs that hybridize directly to adjacent sequences on mRNA molecules in crude sample lysates (Supplementary References 1). After hybridization, adjacent detector oligonucleotides are ligated and amplified by PCR, during which sample-specific barcodes are incorporated for next-generation sequencing. This workflow reduces dependence on conventional RNA isolation and reverse transcription steps, thereby limiting potential material loss and technical variability in small or partially degraded clinical specimens. These characteristics make TempO-Seq particularly suitable for archival formalin-fixed paraffin-embedded (FFPE) tissue and small renal allograft biopsy samples, where available material is often limited. Previous studies have shown that TempO-Seq can generate reproducible gene-expression profiles with favorable signal-to-noise characteristics and reduced unexplained variance compared with conventional RNA-seq approaches (Supplementary References 1,2). In addition, Word et al. showed that whole-transcriptome TempO-Seq applied to lysed cells generated expression profiles broadly comparable to conventional RNA-seq from purified RNA across 39 human cell lines, with high reproducibility between TempO-Seq runs and concordant expression for most shared genes (Supplementary References 3). These findings support the use of TempO-Seq for high-throughput molecular profiling of limited clinical biopsy material, while recognizing that platform-specific effects may remain for selected gene classes. Within transplant molecular diagnostics, targeted profiling is often performed using the Banff Human Organ Transplant (B-HOT) panel on the NanoString nCounter platform (Supplementary References 4). Both NanoString-based targeted profiling and TempO-Seq are compatible with challenging clinical tissue formats, but they differ in technical design and transcriptomic scope. The B-HOT panel uses fluorescently barcoded probes for direct digital counting of endogenous transcripts without enzymatic amplification (Supplementary References 4). TempO-Seq instead uses a ligation- and amplification-based assay followed by a sequencing readout. In addition, whereas the B-HOT panel is restricted to a curated 770-gene set focused on known pathways of allograft rejection, tolerance, and immune injury (Supplementary

References 4,5), the whole-transcriptome TempO-Seq assay used in this study enabled broader transcriptome coverage. This allowed assessment of established transplant-associated pathways while also permitting evaluation of molecular programs outside predefined transplant gene panels. For the present study, tissue preparation for TempO-Seq followed the extraction-free FFPE workflow described by Trejo et al. for FFPE tissue sections and histology-directed tissue areas (Supplementary References 6). In brief, this approach allows whole-transcriptome gene-expression profiling directly from FFPE-derived tissue lysates without conventional RNA extraction, reducing material loss from small biopsy specimens. This is particularly relevant for renal allograft biopsy studies, where tissue availability is limited and archival FFPE material is commonly used. The detailed procedures for FFPE section processing, tissue lysis, and TempO-Seq library preparation have been described previously and were therefore not reproduced step by step here (Supplementary References 6). The resulting TempO-Seq expression matrix was used for downstream pathway-level analysis with GSVA and cell-type deconvolution with CIBERSORTx, as described below.

**GSVA Pathway Scoring**

Biopsy-level transcriptomic pathway activity was quantified using gene set variation analysis (GSVA). TempO-Seq expression data were processed to obtain a gene-level expression matrix with genes as rows and biopsy samples as columns. Gene identifiers were harmonized to current gene symbols, and probes mapping to the same gene were aggregated before pathway analysis. The final expression matrix was converted to numeric format and checked for compatibility with downstream GSVA implementation. Sample metadata were imported separately and aligned to the expression matrix by matching sample identifiers; metadata rows were reordered to correspond exactly to the column order of the expression matrix. Pathway definitions were derived from Molecular Microscope Diagnostic System gene sets provided in Gene Matrix Transposed format. Each gene set was parsed into a list of unique gene symbols. Gene-set representation in the TempO-Seq expression matrix was evaluated by calculating the number and proportion of genes present for each gene set. GSVA was performed using the **GSVA** package on log2-transformed CPM expression values. Gene-set enrichment scores were calculated using the GSVA::gsvaParam() parameter-object workflow followed by GSVA::gsva(). The expression matrix and curated gene-set list were supplied as exprData and geneSets, respectively; no additional GSVA kernel or method parameters were specified, and package defaults were used. Gene sets represented by fewer than 2 genes in the expression matrix were excluded before GSVA. The final GSVA input was therefore restricted to adequately represented transplant-related gene sets. GSVA was applied to the normalized gene-level expression matrix using the curated gene-set collection, resulting in a pathway-by-biopsy matrix of enrichment scores. Each GSVA score represented the relative activity of a given pathway within a biopsy. The resulting matrix was transposed to biopsy-level format and merged with sample metadata. Pathways exhibiting near-zero variance across biopsies were excluded to restrict downstream analyses to informative molecular features. Biopsy identifiers were harmonized across molecular and reader-study datasets using a standardized naming convention. Biopsy-level GSVA scores were then merged into the reader-biopsy dataset, assigning to each reader evaluation the pathway profile of the corresponding biopsy. Analyses were restricted to biopsies with available molecular data. Reader-level GSVA summaries used for annotation of the reader PCA axes were derived as described in the section ‘Post hoc Molecular Annotation of Reader PCA Axes.’

**CIBERSORTx Deconvolution**

To complement pathway-level analyses, biopsy-level cellular composition was inferred from the TempO-Seq transcriptomic profiles using CIBERSORTx. Deconvolution was performed in absolute mode through the CIBERSORTx web portal using log2-normalized bulk expression values as input (Newman et al., Nat Biotechnol 2019; https://cibersortx.stanford.edu; Supplementary References 7). S-mode batch correction was applied to improve compatibility between the single-cell reference and bulk transcriptomic data. Absolute mode was selected because it generates cell-type abundance scores that are not constrained to sum to one across cell populations. The reference signature matrix was generated from an integrated single-cell RNA-sequencing dataset of kidney allograft biopsies. This reference included 70,470 cells from 16 biopsy samples, comprising 8 non-rejecting and 8 donor-specific antibody-positive antibody-mediated rejection samples, compiled from published kidney allograft biopsy cohorts. Cell clusters were annotated to capture immune and kidney-resident parenchymal cell populations using marker-gene profiles and high-resolution kidney atlas resources as guidance. Cells annotated as proliferating were excluded because these profiles may reflect cell-cycle state rather than a distinct biologically interpretable lineage. To meet CIBERSORTx upload-size constraints while preserving the representation of annotated cell populations, the single-cell reference was randomly subsampled. For compatibility with the TempO-Seq assay, the reference matrix was restricted to genes represented on the assay manifest, resulting in a final signature matrix containing 18,276 genes. Deconvolution performance was evaluated using simulated pseudobulk samples generated from the single-cell reference, comparing estimated CIBERSORTx cell-type abundance scores with known cell-type proportions. This validation showed high Pearson correlations for major cell populations, including plasma cells. CIBERSORTx output was imported into R and processed using the tidyverse framework. Cell-type columns were standardized, converted to numeric format, and reviewed for consistency before downstream analysis. Biopsy identifiers were harmonized across molecular and reader-study datasets using a standardized naming convention. Biopsy-level cell-type abundance scores were then merged into the reader-biopsy dataset, assigning the inferred cellular profile of each biopsy to each corresponding reader evaluation. Analyses were restricted to biopsies with available molecular data. Primary CIBERSORTx-based reader PCA annotation analyses were restricted to immune cell types, and reader-level immune-cell summaries were derived as described in the section “Post hoc Molecular Annotation of Reader PCA Axes.”

**Post hoc Molecular Annotation of Reader PCA Axes**

Biopsy-level molecular data were incorporated as a post hoc annotation layer for the leading reader principal components. Molecular features were not used to construct the reader-by-reader similarity matrix or to derive the PCA coordinates. Because PCA coordinates were defined at the reader level, whereas transcriptomic features were measured at the biopsy level, biopsy-level molecular features were converted into reader-level molecular summaries before association with PCA coordinates. Unconditioned reader-level averages across all biopsies were not used, because all readers evaluated the same biopsy set and such averages would primarily reflect the shared case composition rather than reader-specific diagnostic behavior. Instead, reader-level molecular summaries were constructed from subsets of biopsies defined by each reader's own diagnostic classifications. These summaries were selected to match the post hoc diagnostic interpretation of the corresponding PCA axis. For PC1, which was interpreted after PCA as reflecting variation in rejection-calling tendency, molecular summaries were calculated across biopsies classified by each reader as rejection. Borderline classifications were excluded because they represent an intermediate category and do not map cleanly to either pole of a rejection-versus-no-rejection contrast. For each reader and molecular feature, the PC1 molecular summary was defined as the mean feature value across biopsies assigned to a rejection category by that reader. This yielded one reader-level value per molecular feature, representing the molecular profile of the biopsies each reader considered sufficiently abnormal to classify as rejection. For PC2, which was interpreted after PCA as reflecting phenotypic orientation among rejection-type diagnoses, reader-level molecular summaries were defined as TCMR-minus-AMR contrasts. For each reader and molecular feature, the mean feature value among biopsies classified by that reader as AMR-only was subtracted from the mean feature value among biopsies classified by that reader as TCMR-only. Mixed rejection, borderline changes, and no rejection were excluded from this analysis because these categories do not map uniquely to either pole of a TCMR-versus-AMR contrast.

Formally, for each reader $r$ and feature $f$, the PC2 molecular summary was calculated as:

$$PC2\ contrast_{r,f} = \bar{x}_{r,f,TCMR} - \bar{x}_{r,f,AMR}$$

where $\bar{x}_{r,f,\mathrm{TCMR}}$ denotes the mean GSVA pathway score or mean immune cell proportion among biopsies classified as TCMR-only by reader $r$, and $\bar{x}_{r,f,\mathrm{AMR}}$ denotes the corresponding mean among biopsies classified as AMR-only by the same reader. Positive values therefore indicate relatively higher feature values in biopsies classified as TCMR-only than in biopsies classified as AMR-only by the same reader, whereas negative values indicate relatively higher feature values in biopsies classified as AMR-only. Reader-level molecular summaries were merged with reader PCA coordinates and evaluated using Spearman correlation with the corresponding principal component. These analyses were used only to biologically annotate reader-derived PCA axes. They were not used to define diagnostic categories, construct the PCA, or determine diagnostic correctness. Resulting associations should therefore be interpreted as molecular characteristics linked to reader-specific diagnostic behavior, not as evidence that readers directly assessed molecular features. For each molecular analysis, features were ranked by effect size, and p values were adjusted for multiple testing using the Benjamini-Hochberg false discovery rate method.

**Computational Implementation and Reproducibility**

All analyses were conducted in R (version 4.5.1) within a scripted workflow to ensure reproducibility. Analyses were conducted in R using broom.mixed (v0.2.9.7), circlize (v0.4.18), ComplexHeatmap (v2.26.1), dplyr (v1.2.1), forcats (v1.0.1), ggplot2 (v4.0.2), GSVA (v2.0.7), gt (v1.3.0), irrCAC (v1.0), janitor (v2.2.1), lme4 (v2.0-1), patchwork (v1.3.2), purrr (v1.2.1), readr (v2.2.0), readxl (v1.4.5), reshape2 (v1.4.5), rlang (v1.2.0), stringr (v1.6.0), tibble (v3.3.1), and tidyr (v1.3.2). Core packages included tidyverse for data manipulation, readxl and janitor for data import and cleaning, irrCAC for agreement coefficient estimation, lme4 for mixed-effects modeling, GSVA for pathway scoring, and ggplot2 for visualization. Custom functions were implemented for construction of rating matrices, bootstrap resampling, and computation of agreement statistics. A fixed random seed (seed = 123) was used for all resampling procedures.

### Supplemental references

1. Yeakley, J. M., Shepard, P. J., Goyena, D. E., et al. A trichostatin A expression signature identified by TempO-Seq targeted whole transcriptome profiling. PLOS ONE, 2017;12(5), e0178302. https://doi.org/10.1371/journal.pone.0178302

2. Bushel, P. R., Paules, R. S., & Auerbach, S. S. A Comparison of the TempO-Seq S1500+ Platform to RNA-Seq and Microarray Using Rat Liver Mode of Action Samples. Frontiers in Genetics, 2018;9, 485. https://doi.org/10.3389/fgene.2018.00485

3. Word LJ, Willis CM, Judson RS, Everett LJ, Davidson-Fritz SE, Haggard DE, et al. TempO-seq and RNA-seq gene expression levels are highly correlated for most genes: A comparison using 39 human cell lines. PLoS ONE. 2025;20(5):e0320862. https://doi.org/10.1371/journal.pone.0320862

4. Smith, R. N., Rosales, I. A., Tomaszewski, K. T., et al. Utility of Banff Human Organ Transplant Gene Panel in Human Kidney Transplant Biopsies. Transplantation, 2023;107(5), 1188-1199. https://doi.org/10.1097/tp.0000000000004389

5. Giarraputo, A., Coutance, G., Aubert, O., et al. Banff Human Organ Transplant Consensus Gene Panel for the Detection of Antibody Mediated Rejection in Heart Allograft Biopsies. Transplant International, 2023;36. https://doi.org/10.1371/journal.pone.0212031

6. Trejo CL, Babić M, Imler E, Gonzalez M, Bibikov SI, et al. Extraction-free whole transcriptome gene expression analysis of FFPE sections and histology-directed subareas of tissue. PLoS One. 2019;14(2):e0212031. https://doi:10.1371/journal.pone.0212031

7. Newman AM, Steen CB, Liu CL, Gentles AJ, Chaudhuri AA, Scherer F, Khodadoust MS, Esfahani MS, Luca BA, Steiner D, Diehn M, Alizadeh AA. Determining cell type

abundance and expression from bulk tissues with digital cytometry. Nat Biotechnol. 2019 Jul;37(7):773-782. https://doi: 10.1038/s41587-019-0114-2

8. Lamarthée B, Callemeyn J, Van Herck Y, et al. Transcriptional and spatial profiling of the kidney allograft unravels a central role for FcyRIII+ innate immune cells in rejection. Nat Commun. 2023 Jul 19;14(1):4359. https://doi.org/10.1038/s41467-023-39859-7

9. Malone AF, Wu H, Fronick C, Fulton R, et al. Harnessing Expressed Single Nucleotide Variation and Single Cell RNA Sequencing To Define Immune Cell Chimerism in the Rejecting Kidney Transplant. J Am Soc Nephrol. 2020 Sep;31(9):1977-1986. https://doi.org/10.1681/asn.2020030326

10. Shi M, Wang Y, Zhang H, Ling Z, Chen X, Wang C, Liu J and Ma Y. Single-cell RNA sequencing shows the immune cell landscape in the kidneys of patients with idiopathic membranous nephropathy. Front. Immunol. 2023;14:1203062. https://doi.org/10.3389/fimmu.2023.1203062

11. Lake BB, Menon R, Winfree S, Hu Q, et al. An atlas of healthy and injured cell states and niches in the human kidney. Nature. 2023 Jul;619(7970):585-594. https://doi.org/10.1038/s41586-023-05769-3

**Supplementary Table S1.** Banff Lesion Scores Assessed by Readers

| Category | Gene sets | Description and role |
|---|---|---|
| **g-score** | Glomerulitis | Scored g0-g3; proportion of glomeruli showing glomerular capillary inflammation. |
| **ptc-score** | Peritubular capillaritis | Scored ptc0-ptc3; peritubular capillary inflammation, incorporating the proportion of involved capillaries and inflammatory cell burden in affected capillaries. |
| **v-score** | Intimal arteritis | Scored v0-v3; most severely affected artery, based on the presence and severity of inflammatory cells in the arterial intima. |
| **i-score** | Interstitial inflammation | Scored i0-i3; proportion of non-scared cortical parenchyma involved by inflammation. |
| **t-score** | Tubulitis | Scored t0-t3; most severely affected preserved cortical tubule, based on the number of mononuclear inflammatory cells within tubular epithelium. |
| **ti-score** | Total inflammation | Scored ti0-ti3; proportion of total cortical parenchyma involved by inflammation, including both scarred and non-scarred cortex. |
| **i-IFTA** | Inflammation in areas of interstitial fibrosis and tubular atrophy | Scored i-IFTA0-i-IFTA3; proportion of scarred cortical parenchyma involved by inflammation. |
| **t-IFTA** | Tubulitis in areas of interstitial fibrosis and tubular atrophy | Scored t-IFTA0-t-IFTA3; tubulitis in atrophic tubules within areas of interstitial fibrosis and tubular atrophy. |
| **ci-score** | Interstitial fibrosis | Scored ci0-ci3; proportion of cortical parenchyma affected by interstitial fibrosis. |
| **ct-score** | Tubular atrophy | Scored ct0-ct3; proportion of cortical parenchyma affected by tubular atrophy. |
| **IFTA** | Interstitial fibrosis and tubular atrophy | Scored IFTA0-IFTA3; overall proportion of cortical parenchyma affected by interstitial fibrosis and tubular atrophy. |

| | | |
|---|---|---|
| **cg-score** | Transplant glomerulopathy | Scored cg0-cg3; extent of glomerular basement membrane double contours in glomerular capillary loops. |
| **cv-score** | Vascular fibrous intimal thickening | Scored cv0-cv3; severity of arterial fibrous intimal thickening and resulting luminal narrowing. |
| **mm-score** | Mesangial matrix increase | Scored mm0-mm3; proportion of non-sclerosed glomeruli showing mesangial matrix expansion. |
| **ah-score** | Arteriolar hyalinosis | Scored ah0-ah3; extent and severity of PAS-positive arteriolar hyaline thickening. |

Scoring summaries describe the general basis of Banff lesion score assignment without reproducing category-specific thresholds. Detailed definitions and thresholds are provided in the current Banff reference guide (Reference 1 – main paper).

**Supplementary Table S2.** Annotated MMDx pathogenesis-based transcript (PBT) gene sets used for GSVA analysis in kidney transplant biopsies.

| Category | Gene sets | Description and role |
|---|---|---|
| **T Cell–Mediated Rejection (TCMR)** | QCAT, TCMR-RATs, TCB | Cytotoxic T cell activation and rejection signatures[1] |
| **Antibody-Mediated Rejection (AMR)** | DSAST, eDSAST, ABMR-RATs, Rej-RATs, RATs, AAG, NKAAG, AEG | Endothelial and NK cell–related pathways; key AMR activity genes[2-3] |
| **Gamma-IFN Response** | GRIT1, GRIT2, GRIT3, IIAAG | IFNG-inducible pathways involved in rejection and tissue injury[4-5] |
| **B Cell / Plasma Cell Activity** | BAT | Markers of B cell and plasma cell infiltration, linked to chronic rejection and fibrosis[6] |
| **Macrophage & Innate Immunity** | QCMAT, AMAT1, MCAT | M1/M2 macrophages and mast cell infiltration; drivers of inflammation and fibrosis[7] |
| **NK Cell Activity** | NKB, NKAAG | NK cell–specific rejection signatures, particularly in AMR[8] |
| **Injury & Repair Programs** | IRRAT, IRITD1, IRITD3, IRITD5, ENDAT | Programs of tubular and endothelial injury and repair[9-10] |
| **Chronic Damage & Fibrosis** | FICOL, DAMP | Collagen deposition, ECM remodeling, and damage-associated signals; hallmarks of chronic allograft dysfunction[11] |
| **Kidney Parenchymal Health** | KT1, KT2 | Normal kidney transcripts; loss indicates dedifferentiation or damage[12-13] |

Original articles:
[1]Hidalgo LG, Einecke G, Allanach K, et al. The transcriptome of human cytotoxic T cells: measuring the burden of CTL-associated transcripts in human kidney transplants. Am J Transplant. 2008;8:637–646.
[2] Hidalgo LG, Sis B, Sellares J, et al. NK cell transcripts and NK cells in kidney biopsies from patients with donor-specific antibodies: evidence for NK cell involvement in antibody-mediated rejection. Am J Transplant. 2010; 10:1812–1822.
[3] Halloran PF, Potena L, Van Huyen JD, et al. Building a tissue-based molecular diagnostic system in heart transplant rejection: the heart Molecular Microscope Diagnostic (MMDx) System. J Heart Lung Transplant. 2017; 36:1192–1200.
[4] Halloran PF, Venner JM, Famulski KS. Comprehensive analysis of transcript changes associated with allograft rejection: combining universal and selective features. Am J Transplant. 2017; 17:1754–1769.
[5] Famulski KS, Einecke G, Reeve J, et al. Changes in the transcriptome in allograft rejection: IFN-γ induced transcripts in mouse kidney allografts. Am J Transplant. 2006; 6:1342–1354.
[6] Einecke G, Reeve J, Mengel M, et al. Expression of B cell and immunoglobulin transcripts is a feature of inflammation in late allografts. Am J Transplant. 2008; 8:1434–1443.
[7] Famulski KS, Einecke G, Sis B, et al. Defining the canonical form of T-cell-mediated rejection in human kidney transplants. Am J Transplant. 2010; 10:810–820.
[8] Hidalgo LG, Sellares J, Sis B, et al. Interpreting NK cell transcripts versus T cell transcripts in renal transplant biopsies. Am J Transplant. 2012; 12:1180–1191.
[9] Famulski KS, de Freitas DG, Kreepala C, et al. Molecular phenotypes of acute kidney injury in kidney transplants. J Am Soc Nephrol. 2012; 23:948–958.
[10] Famulski KS, Broderick G, Einecke G, et al. Transcriptome analysis reveals heterogeneity in the injury response of kidney transplants. Am J Transplant. 2007; 7:2483–2495.
[11] Famulski KS, Reeve J, de Freitas DG, et al. Kidney transplants with progressing chronic diseases express high levels of acute kidney injury transcripts. Am J Transplant. 2013; 13:634–644.
[12] Einecke G, Kayser D, Vanslambrouck JM, et al. Loss of solute carriers in T cell-mediated rejection in mouse and human kidneys: an active epithelial injury-repair response. Am J Transplant. 2010; 10:2241–2251.
[13] Einecke G, Broderick G, Sis B, et al. Early loss of renal transcripts in kidney allografts: relationship to the development of histologic lesions and alloimmune effector mechanisms. Am J Transplant. 2007; 7:1121–1130.

**Abbreviations:** AAG, acute antibody-mediated injury-associated gene set; ABMR-RATs, antibody-mediated rejection-associated transcripts; AEG, antibody-mediated rejection-associated endothelial genes; AMAT1, alternative macrophage activation transcripts 1; BAT, B cell-associated transcripts; DAMP, damage-associated molecular pattern transcripts; DSAST, donor-specific antibody-selective transcripts; eDSAST, endothelial donor-specific antibody-selective transcripts; ENDAT, endothelial cell-associated transcripts; FICOL, fibrillar collagen transcripts; GRIT1, gamma-interferon and rejection-induced transcripts 1; GRIT2, gamma-interferon and rejection-induced transcripts 2; GRIT3, new gamma-interferon and rejection-induced transcripts; IIAAG, interferon-inducible acute antibody-mediated gene set; IRITD1, injury- and repair-induced transcripts, early; IRITD3, injury- and repair-induced transcripts, intermediate; IRITD5, injury- and repair-induced transcripts, late; IRRAT, injury-repair response-associated transcripts; KT1, kidney transcripts set 1; KT2, kidney transcripts set 2; MCAT, mast cell-associated transcripts; NKAAG, NK cell-expressed antibody-mediated rejection activity genes; NKB, NK cell transcript burden; PEC, parietal epithelial cell-associated transcripts; QCAT, quantitative cytotoxic T lymphocyte-associated transcripts; QCMAT, quantitative constitutive macrophage-associated transcripts; RATs, rejection-associated transcripts; Rej-RATs, rejection-associated transcripts; TCB, T cell transcript burden; TCMR-RATs, T cell-mediated rejection-associated transcripts

**Supplementary Table S3.** Overview of MMDx gene sets and their included genes.

| Gene Set | Genes |
|---|---|
| **AAG** | CCL3, CCL4, CCL4L1, CXCL10, CXCL11, CXCL9, FGFBP2, GBP1, GBP4, GNLY, GZMB, GZMH, IDO1, KLRD1, NKG7, PLA1A, PRF1, RARRES3, S1PR5, WARS |
| **ABMR-RATs** | ACKR1, ADAM15, ADGRL4, APLN, APOBEC3A, APOL1, APOL3, B2M, BCL2A1, BCL6B, BTN3A2, BTN3A3, C2CD4B, CALHM6, CCL3, CCL4, CCL4L1, CD160, CD244, CD74, CDH13, CDH5, CLEC9A, COL13A1, CRTAM, CST7, CX3CL1, CX3CR1, CXCL10, CXCL11, CXCL9, ECSCR, ESM1, FCGR3A, FCN1, FCRL6, FGFBP2, GABBR1, GBP1, GBP4, GBP5, GIMAP6, GJD3, GNG11, GNLY, GZMB, HLA-A, HLA-B, HLA-C, HLA-DOA, HLA-DPA1, HLA-DPB1, HLA-DRA, HLA-DRB1, HLA-E, HLA-F, HSPA12B, ICAM1, ICAM2, IDO1, IFI27, IRF1, ITGAL, KLF4, KLRC1, KLRC3, KLRD1, KLRF1, LAYN, LILRA1, LILRB2, LST1, LYPD5, MALL, MMRN2, MS4A7, MYBL1, NCR1, NKG7, NLRC5, PECAM1, PLA1A, PLAT, PRF1, PSMB9, RAMP3, RAPGEF5, RASIP1, ROBO4, S100A3, S1PR1, S1PR5, SELE, SH2B3, SH2D1B, STX11, TAP1, TBX21, TEK, TM4SF1, TM4SF18, TNF, TNFAIP3, TRDC, TRDV3, WARS, XCL1 |
| **AEG** | ADGRL4, CDH13, CDH5, ECSCR, ERG, GNG11, IFI27, MALL, MMRN2, NOS3, PECAM1, RAPGEF5, RASIP1, ROBO4, TEK, TM4SF18 |
| **AMAT1** | ARG1, CD163, CD163L1, CLEC10A, CLEC7A, MMP12, MMP9, MRC1, RNASE3, THBS1 |

| | |
|---|---|
| **BAT** | ABCB4, ADAM28, AFF3, AREG, ARHGAP24, BACE2, BANK1, BCAS4, BCL11A, BCL2L11, BIRC3, BLK, BTLA, BTNL9, CCDC188, CCDC50, CCR6, CD19, CD1C, CD200, CD22, CD72, CD79A, CD79B, CDCA7L, CHPT1, COCH, CPNE5, CR1, CR2, DENND5B, E2F5, EAF2, EBF1, EML6, FAM117B, FCER2, FCRL1, FCRL2, FCRL5, FCRLA, GABBR1, GNG7, GPR18, HDAC9, HERPUD1, HLA-DOA, HLA-DOB, IFNLR1, IFT57, IGHD, IGK, ITPR1, KIAA0125, KLHL14, LCN10, LINC00494, LY9, MACROD2, MEF2C, METTL8, MICAL3, MIR600, MS4A1, NAPB, NIBAN3, NR4A2, OSBPL10, P2RX5, PAWR, PAX5, PCDH9, PDLIM1, PEG10, PKIG, PLEKHG1, PNOC, POU2AF1, POU2F2, PRICKLE1, RAB30, RALGPS2, RNF141, RUBCNL, SCN3A, SEL1L3, SEMA4B, SKAP2, SLC15A2, SLC9A7, SMIM14, SNX29, SPIB, STRBP, TCF4, TCL1A, TCP11L2, TENT5C, TLR10, TMEM154, TNFRSF13C, TSPAN13, VPREB3, ZBTB18, ZDHHC23 |
| **DAMP** | HDGF, HMGB1, S100A12, S100A8, S100A9, S100B |
| **DSAST** | ACKR1, CDH13, CDH5, COL13A1, CX3CR1, FGFBP2, GNG11, GNLY, ICAM2, KLRF1, MALL, MYBL1, PGM5, PLA1A, PLAT, ROBO4, SH2D1B, SOX7, TEK, TM4SF18 |

| | |
|---|---|
| **ENDAT** | ABI3BP, ACE, ADCY4, ADRA2A, ALPL, ANGPT1, ANGPT2, BAGE, BAGE2, BMP6, BMX, CARD16, CASP1, CAV1, CAVIN2, CCN2, CD34, CD36, CDH26, CDH5, CETP, CMAHP, COL18A1, COL4A1, COL4A2, CRIM1, CYYR1, DAB2, DIAPH2, DLC1, DNAJB1, EDN1, EFEMP1, EHD4, EMCN, ENPEP, ENPP2, ESM1, FABP4, FGD5, FHOD1, FLI1, FOSB, FOXF2, GEM, GJA4, GMFG, HEYL, HHIP, HOXD1, HSPA6, HSPG2, ICAM2, IGFBP7, JUNB, KDR, KLF4, KMO, KMT2C, LAMB1, LAMC1, LDB2, LOC646214, LOXL2, MAOB, MCAM, MEOX1, MFNG, MGP, MPZL2, MSL3, NFATC4, NID2, NOD1, NOS3, NOX4, NPR1, NR4A1, NRF1, NTN4, NXF1, OPN3, PAK2, PALMD, PDGFB, PDGFRB, PECAM1, PLSCR4, PLVAP, PNP, PODXL, PPP1R16B, PRKACA, PTGIR, RAI14, RAMP2, RASGRP2, RASIP1, RGS5, RHOJ, RPGR, S1PR1, SEC14L1, SELE, SELP, SERPINE1, SNAI1, SOX18, SRGN, TBX2, TCF4, TDO2, TEK, THBS1, TRPV2, UACA, VCAM1, VWF, ZNF521 |
| **FICOL** | COL11A1, COL1A1, COL1A2, COL27A1, COL3A1, COL5A1, COL5A2 |
| **GRIT1** | B2M, BST2, C1R, C2, CD74, CXCL10, CXCL9, ERAP1, GBP2, GBP4, GBP6, GNL1, HERC6, HLA-A, HLA-B, HLA-C, HLA-DMA, HLA-DMB, HLA-DQA1, HLA-DQA2, HLA-DQB1, HLA-DRA, HLA-DRB1, HLA-E, IFI27L1, IFIT2, IL18BP, IRF7, IRGM, LGALS3BP, MPEG1, PARP14, PLAAT3, PSMB10, PSMB8, PSMB9, PSME1, SERPINA10, SERPING1, STAT1, TAPBP, XDH |

| | |
|---|---|
| **GRIT2** | ADAR, AIF1, AK7, AKT3, AREL1, ATP8A1, BATF2, BSPRY, BST1, C1S, CASP12, CASP7, CCL19, CCL5, CCR9, CCRL2, CD274, CD40, CD86, CIITA, CMPK2, CTSC, CYBB, CYP4V2, DCANP1, DDX60, DHX58, DNASE1L3, DTX3L, ECE2, EIF2AK2, EIF4E3, ENC1, ENPP4, EXOC3L4, FABP7, FAM49A, FBXO6, FCGR1A, FCGR3A, FCGR3B, GSDMD, HCK, HERC6, HK3, HLA-A, HLA-DOB, HPSE, IDNK, IFI16, IFI35, IFI44, IFIH1, IFIT1, IFIT3, IL18, INPP1, IRF1, IRF8, ISG15, ITGA4, LACC1, LAIR1, LY86, MIR5193, MISP, MLKL, NAAA, NOD1, OAS1, OASL, P2RY14, PARP11, PARP12, PARP14, PARP3, PARP9, PEX26, PFKP, PHF11, PIGR, PIK3IP1, PLCL2, PNP, POU2AF1, PPA1, PRDM1, PTPN6, RARRES2, RBL1, RBM43, RNASE6, RNF19B, RNF213, RSAD2, RTP4, SAMHD1, SECTM1, SELL, SERPINB9, SLAMF6, SLAMF8, SLC2A6, SNX10, STAT1, STAT2, TAP2, TAPBPL, THEMIS2, TIFAB, TLR3, TMEM140, TMPRSS4, TNFRSF14, TNFSF10, TOR3A, TOX, TRAFD1, TRIM21, TRIM5, UBE2L6, UPP1, USP18, VCAM1, VRK2, WARS, XAF1, ZC3H12D, ZC3HAV1, ZMYND15 |

| | |
|---|---|
| **GRIT3** | ACKR4, ACSL5, ANGPTL2, ANKRD22, APOBEC3G, APOL1, APOL2, APOL3, APOL4, APOL6, AQP1, ARNT2, ASPHD2, ATF3, BATF2, BST2, BTN3A1, BTN3A2, BTN3A3, C1QB, C1QC, C1R, C1S, C1orf21, C2, C5orf56, CALHM6, CARD16, CASP1, CCL2, CCL20, CCL8, CD274, CD38, CD40, CD74, CEBPD, CHFL, CLIC2, CMPK2, COL8A1, CTSS, CXCL1, CXCL10, CXCL11, CXCL2, CXCL9, DDIT4L, DDX58, DDX60, DTX3L, EDNRA, EFCAB2, EPSTI1, F3, FAM107A, FAM117B, FAM241A, FBXO6, FCGR1A, FCGR1B, GBP1, GBP2, GBP3, GBP4, GBP5, GCH1, GZMA, HCAR2, HCP5, HELLPAR, HLA-A, HLA-B, HLA-C, HLA-DMA, HLA-DMB, HLA-DOA, HLA-DPA1, HLA-DPB1, HLA-DQA1, HLA-DQA2, HLA-DQB1, HLA-DRA, HLA-DRB1, HLA-DRB4, HLA-E, HLA-F, HLA-G, ICAM1, IDO1, IDO2, IFI27, IFI30, IFI35, IFI44, IFI44L, IFI6, IFIH1, IFIT1, IFIT2, IFIT3, IFIT5, IFITM1, IGHA1, IL15, IL15RA, IL18BP, IL32, IRF1, IRF8, IRF9, ISG15, ISG20, ITGA6, JAK2, KCNJ2, KLF4, LAP3, LGALS3BP, LGALS9, LIMK2, LMNB1, LOC100129518, LOC154761, LRRK2, MAP3K7CL, MDK, MICB, MIR5193, MX1, N4BP2L1, NLRC5, NME7, NRK, NTN4, OAS1, OAS2, OAS3, P2RY14, PARP12, PARP14, PARP9, PCDH7, PCLO, PDE4B, PIGR, PIK3AP1, PLA1A, PLAAT4, PLSCR1, PMAIP1, PSMB10, PSMB8, PSMB9, PSME2, PSTPIP2, RARRES1, RELN, RHOBTB3, RIPK2, RND3, RNF19B, RNF213, RTP4, SAMD9L, SAMHD1, SECTM1, SERPING1, SLAMF7, SLC15A3, SLC27A2, SLC2A5, SLC6A12, SOCS1, SOCS3, SP110, SP140, SP140L, SRGN, STAMBPL1, STAT1, STAT2, SUCNR1, TAGAP, TAP1, TAP2, TAPBPL, TCAF2, TCIM, TGM2, TIFA, TMEM252, TNC, TNFAIP2, TNFAIP6, TNFSF10, TNFSF13B, TNFSF15, TOX3, TRIM21, |

| | |
|---|---|
| | TRIM22, TYMP, UBE2L6, USP6NL, VAMP5, VRK2, WARS, XAF1 |

| | |
|---|---|
| **IIAAG** | CX3CL1, CXCL10, CXCL11, CXCL9, GBP1, GBP4, IDO1, IL18BP, PLA1A, WARS |

| | |
|---|---|
| **IRITD1** | ABCB1, ACER2, ACOT1, ACOT2, ADGRG1, ADM, AIF1L, AKIP1, ALAS1, ANGPTL4, APLN, APOA4, APOH, APOLD1, AQP2, AQP6, AQP7, AQP7P1, ARF6, ARHGDIA, ARHGEF1, ASB11, ATN1, BLVRB, BORCS7, BORCS7-ASMT, BTG2, CAMK2N2, CCDC120, CCDC97, CCK, CCM2, CCN2, CCND1, CDH5, CDS2, CDT1, CEP85, CFL1, CHTF8, CLDN15, COL5A3, CPEB1, CTSB, CTSV, CYP2B6, CYP2B7P, CYP2S1, CYP4A11, CYP4A22, DAO, DBI, DDX47, DEPP1, DHRS3, DMPK, DVL1, ECI1, FAM114A2, FAM71E1, FBP2, FGA, FGB, FGG, FKBP11, FLOT1, FOXQ1, GAS6, GLUD1, GLUD2, GMPR, GPR182, GPX4, GREM2, GSTM3, HBCBP, HDGF, HEBP1, HESX1, HMGA1, HMGCS2, HOXB7, HOXB8, HOXD8, HS6ST1, HSPB6, HSPB8, IL34, IMMP2L, INHBB, JUP, KANK3, KAZN, KCNIP2, KCTD17, KIFC3, KLF10, KLF2, KLHDC8A, KRT20, LENG8, LGI2, LIMS2, LOC100505585, LOC100509620, LOC101930168, LOC105375355, LONRF3, LRG1, LRRC59, MAD2L2, MAOA, MAOB, MAPRE3, MAX, MCL1, MEX3D, MGST1, MMD, MRPL9, MRPS6, NDUFA4L2, NMRK1, NOS3, NPL, NT5DC2, NUAK2, P2RX4, PCGF2, PCGF5, PDK4, PER1, PGM2, PKM, PLET1, PLIN2, PLIN4, PLIN5, PLPP5, PMEPA1, PPM1J, PRUNE1, PSAT1, PTP4A3, RASGEF1B, RBP7, RFLNB, RGCC, RNASE1, RNH1, RTKN, S1PR1, SAA1, SAA2, SAA2-SAA4, SCARB2, SCG5, SEMA3F, SEPTIN11, SGK1, SH3TC2, SHB, SHISA5, SLC13A1, SLC14A2, SLC16A6, SLC20A1, SLC25A30, SLC25A34, SLC9A3R2, SPTLC2, SSH3, STX3, SUSD1, SYDE1, SYT11, TMBIM1, TMEM37, TMEM82, TNFAIP1, TPCN1, TSPAN4, TTPA, UBALD2, UCP2, UPK3B, VAMP2, VAT1, VEGFB, VNN1, WBP2, ZBTB16, ZCCHC3 |

| | |
|---|---|
| **IRITD3** | ABCA1, ABI2, ACOT7, ACSL4, ACTN1, ADA, ADAMTS1, ADGRE5, ADGRG2, AEBP1, AHNAK, AIDA, AJUBA, AKAP12, ALDH1A2, ANKRD6, ANXA1, ANXA2, ANXA3, ANXA5, ANXA6, ARG2, ARPC1B, ARVCF, ATF4, BAG2, BCL3, BHLHE40, BSPRY, C15orf39, C3, C4orf46, CALM1, CAMK1, CARHSP1, CAVIN1, CAVIN3, CCN1, CCN4, CD14, CD248, CD9, CDC25B, CDKN2AIPNL, CEBPB, CEBPD, CENPT, CHEK1, CHRNB1, CLDN1, CLDN3, CLDN4, CLDN7, CLIC1, CMTM3, COL15A1, COL4A1, COL4A2, COL8A1, COLGALT1, COMMD9, COMT, CORO1C, COX6B2, CP, CPE, CPNE8, CRISP3, CRLF1, CRLF2, CSRP1, CSTB, CTNS, CTPS1, CTSD, CTXN1, CX3CL1, CXXC5, CYGB, DAP, DDIT4, DDR1, DDX39A, DHODH, DLGAP4, DNAJC25, DPYSL3, DUT, EDN1, EEF1B2, EFEMP1, EIF4EBP1, EIF6, ELF3, EMILIN1, F2R, F2RL1, FABP4, FAM126A, FKBP10, FLVCR1, FMNL2, FNDC4, FOSL2, FST, FXYD5, GC, GDF15, GIMAP8, GJA1, GNAI2, GPIHBP1, GPRC5A, GPT2, GRN, GSTO1, HACD1, HAVCR1, HEXA, HNRNPA1, HP, HSD3B7, HSPB1, HSPB11, HTRA3, ICAM1, IER3, IFITM10, IGSF8, IHH, IL17RB, IMMT, IMPDH2, JAM2, JUN, JUND, KCNK1, KCTD1, KCTD10, KDELR3, KIFC1, KLF5, KLF6, KRT18, KRT4, KRT7, KRT8, LAD1, LEPR, LEPROT, LGALS1, LGALS3, LHFPL6, LITAF, LMNA, LOC100128751, LOC100508408, LOXL2, LXN, MAB21L3, MAL2, MAN1C1, MAP3K1, MAP4, MAP7D1, MAPKAPK3, MARCKSL1, MCL1, MCM4, MCM7, ME3, MFSD10, MIR1257, MMP24, MMP8, MT2A, MVP, MYD88, MYH9, MZT2A, NFKBIA, NOTCH3, NRAS, NREP, NRM, NUDT21, NUPR1, ORM1, OSMR, OSTF1, PABPC1, PAGR1, PANX3, PAPPA, PCNA, PDGFB, PEA15, PFKP, PGD, PHC2, PHGDH, PHLDA1, PIEZO1, PITPNM1, |

| | |
|---|---|
| | PLAT, PLAUR, PLP2, PNRC1, PPP1R14B, PPP1R18, PRCD, PRKCH, PROS1, PRR15, PRSS23, PSAP, PTPN12, PTPRF, RAB3D, RALY, RAN, RAPH1, RASD1, RASL12, RBPMS, RCSD1, RELA, RGS4, RHOB, RHOC, RHOD, RIOK1, RPL12, RPL13, RPL13A, RPL32, RPS10, RPS11, RPS12, RPS14, RPS18, RPS21, RPS24, RPS9, RRAD, RRAS, S100A10, S100A11, S100A6, SBSN, SCARB1, SELENOH, SEMA7A, SERPINA3, SERPINB6, SERTAD4, SH3BGRL3, SIPA1, SIRPA, SLC44A2, SLC7A5, SLC7A6, SMOX, SOCS2, SPARC, SPECC1, SPHK1, SPIDR, SPRR1A, SPSB1, SSBP3, SSBP4, STAP2, STAT3, STEAP1, SYNPO, TACSTD2, TAGLN, TAGLN2, TES, TGFB1, TGM2, THBD, TIMELESS, TMCC2, TMCC3, TMEM107, TMEM158, TMEM43, TMEM45A, TMEM98, TNFRSF12A, TNFRSF1A, TOB2, TPM2, TPM4, TRIM47, TSC22D1, TSPO, TTLL10, TUBA1B, TUBB, TUBB2A, TUBB2B, TUBB6, VARS, VASP, VIM, WFDC2, WSB1, YBX3, YIPF1, YWHAH, ZFP36L1, ZNF526, ZNF579, ZYX |

| | |
|---|---|
| **IRITD5** | ABCB8, ACTA2, ADAM12, ADAMTS2, ADGRE1, ADIPOQ, AKR1C4, ALAS2, ALX1, ANKH, AOC1, APOBEC1, APOE, ARHGDIB, ARRB1, ASPM, AXL, BAGE, BAGE2, BEX1, BGN, BIRC3, BSG, C1QA, CALCRL, CAPN5, CASP8AP2, CCDC34, CCNF, CCR2, CDC20, CDH11, CENPF, CFD, CFH, CFI, CKAP2, CLU, CNN2, CNP, CNTN1, COL14A1, COL18A1, COL1A1, COL1A2, COL3A1, COL5A1, COL5A2, COL6A1, COL6A2, COL6A3, COLEC12, CRYAB, CSF2RA, CTSK, CTSS, CX3CR1, CXCL12, CYP1B1, DCN, DDIT4L, DDR2, DKFZp686K1684, DNMT1, DPT, DPY19L1, DSEL, DSG2, DSP, DYSF, ECT2, EFEMP2, EGFLAM, EPB41L2, ESCO2, F13A1, FAM49B, FBN1, FCER1G, FDFT1, FKBP7, FLI1, FMC1, FN1, FRK, FSCN1, FSTL1, GIMAP1, GOLM1, GPC3, GPM6B, GPR153, GPSM1, GSN, GUCY1A1, H2AFZ, HBA1, HIST1H2AB, HMGN2, HMMR, HOXB3, ID3, IL33, IRF9, ITPRIPL2, KANK2, KIF11, KMT2C, KRT19, KYNU, LBP, LCP1, LGALS9, LOC100127888, LOC100506558, LOC728743, LPAR1, LTF, LUM, LY6E, LYN, LYZ, MARCKS, MARCKSL1, MCM2, MCM6, MDK, MFAP5, MGP, MIS18BP1, MMD, MMP2, MMP7, MRC2, MSRB3, MUC1, MXRA7, MYADM, MYH9, MYO1B, MYOF, NDN, NEK2, NID1, NPNT, NUB1, NUSAP1, OLFML3, PAN3, PAQR5, PBK, PDPN, PENK, PGLYRP1, PKDCC, PLA2G4A, PLTP, PNISR, PPBP, PRKAG2, PRPF4B, PTGDS, PTGER4, PTGFRN, PYCARD, RAB31, RAB8B, RCAN2, RCN3, REPIN1, RGS2, RHEB, RRM1, RRM2, S1PR2, SERPINA1, SERPINE2, SERPINH1, SH3BGRL, SLC19A1, SLC38A2, SLC5A1, SLC7A1, SLCO4A1, SMAD1, SMARCD3, SMOC2, SNCA, SPON1, SRPX2, SSPN, STAB1, STC2, SULF1, SYTL2, TCF25, TGFBI, THBS2, TIFA, TIMP2, TMEM176B, TNC, TRA2A, TUBA1A, TUBA1B, TUBB2B, |

| | |
|---|---|
| | TYROBP, VSTM4, WDR86, ZBTB21, ZNF462, ZNF775 |

| | |
|---|---|
| **IRRAT** | ADAM9, ADAMTS1, AKAP12, CDH6, CTSS, EGR1, EVI2A, FOS, ITGB3, LCN2, LTF, MEGF11, NFKBIZ, NNMT, OLFM4, OSMR, PI15, PTPRC, PTX3, RARRES1, S100A8, SERPINA3, SLPI, TMEM252, VCAN |

| | |
|---|---|
| **KT1** | AADAC, AADAT, AASS, ABAT, ABCC2, ABCC9, ABCG2, ABHD14B, ABHD3, ACAD10, ACADM, ACAT1, ACE2, ACKR3, ACOT12, ACOT4, ACOX1, ACOX2, ACSM1, ACSM2A, ACSM2B, ACSM3, ACSM5, ACY3, ADGRF5, ADGRG1, ADGRL4, ADH1A, ADH1B, ADH1C, ADHFE1, ADRA2B, ADTRP, AFAP1L1, AFAP1L2, AFM, AGTR1, AGXT2, AIF1L, AK4, AKR1C3, AKR1D1, AKR7A2, ALDH1L1, ALDH4A1, ALDH5A1, ALDH6A1, ALDH7A1, ALDH8A1, ALDH9A1, ALDOB, ALPL, AMACR, AMN, ANKS4B, ANKZF1, APLNR, APOM, AQP1, AQP11, AQP2, AQP3, AQP4, AQP6, ARHGAP24, ARNT2, ARSG, ASB9, ASPA, ASS1, ATP11A, ATP1B1, ATP6V0A4, ATP6V0E2, ATP6V1B1, B3GALT5, BCKDHA, BDH2, BHMT2, BICC1, BMP4, BMP6, BMP7, BPHL, C11orf54, C11orf58, C16orf58, C1QTNF3, C1orf100, C1orf210, C2orf80, C8G, CA12, CA14, CA3, CA4, CA5B, CACHD1, CALB1, CALML4, CASC4, CCDC198, CCKAR, CDA, CDCP1, CDH16, CDHR2, CDKL1, CDO1, CELA1, CGNL1, CGREF1, CHCHD10, CHPT1, CHURC1, CIDEB, CLCNKA, CLCNKB, CLDN10, CLDN2, CLDN8, CLRN3, CLTRN, CLYBL, CMBL, CNDP1, COASY, COBL, COL4A3, COL4A4, COX3, CPT2, CRYL1, CRYM, CRYZ, CSAD, CTH, CTSF, CUBN, CYB561, CYP24A1, CYP2A13, CYP2A6, CYP2E1, CYP4A11, CYP4A22, CYP4B1, CYP7B1, CYSTM1, DAB2, DAO, DBT, DCLK3, DCXR, DDAH1, DDC, DDO, DECR1, DEGS2, DEPTOR, DGAT2, DHTKD1, DIO1, DLEU7, DMAC2L, DMGDH, DMTN, DNAJC12, DNAJC22, DNAJC6, DNASE1, DNASE2, DPEP1, DPYD, DUSP22, ECHDC2, EFHD1, EGF, EGFL6, EHF, EHHADH, ELK4, ELOVL2, ELOVL7, EMCN, EMX2, ENPEP, ENPP2, ENPP3, ENPP6, ENTPD4, ENTPD5, EPB41L5, EPCAM, EPHX2, EPS8L2, ERBB3, ERC2, ESRRG, EVA1A, F13B, FADS3, FAH, FAHD1, FAM13A, FAM149A, FAM151A, FAM186A, FBLN5, FGA, FGF1, FHL1, FHOD3, FLVCR2, FMO1, FMO2, FMO5, FN3K, FOLH1, FOLR1, FOXI1, FRZB, FUT9, FXYD2, FXYD4, |

| | |
|---|---|
| **KT2** | COX3, LOC100288152, SLC10A2, SLC12A1, SLC12A3, SLC13A1, SLC13A2, SLC13A3, SLC14A2, SLC15A2, SLC16A2, SLC16A4, SLC16A9, SLC17A1, SLC17A3, SLC18A1, SLC19A3, SLC1A1, SLC22A1, SLC22A12, SLC22A13, SLC22A17, SLC22A18, SLC22A2, SLC22A4, SLC22A5, SLC22A7, SLC22A8, SLC22A9, SLC23A1, SLC25A15, SLC26A1, SLC26A4, SLC2A2, SLC2A4, SLC2A5, SLC30A2, SLC34A1, SLC34A3, SLC37A4, SLC38A3, SLC39A5, SLC39A8, SLC3A1, SLC40A1, SLC4A1, SLC4A11, SLC4A4, SLC4A9, SLC5A1, SLC5A11, SLC5A2, SLC5A8, SLC5A9, SLC6A13, SLC6A18, SLC6A19, SLC7A13, SLC7A4, SLC7A7, SLC7A9, SLC9A2, SLC9A3R2, SLCO4C1 |
| **MCAT** | CPA3, FCER1A |
| **NKAAG** | CCL4, CD160, CST7, FGFBP2, FGR, GNLY, GZMB, GZMH, KIR2DL2, KIR2DL3, KLRC3, KLRD1, KLRF1, NKG7, PRF1, S1PR5, SH2D1B, TBX21, TRDC, XCL1 |
| **NKB** | CX3CR1, KLRF1, MYBL1, SH2D1B |
| **QCAT** | CD2, CD3D, CD8A, CST7, CXCR6, GNLY, GPR171, GZMA, GZMB, GZMK, HOPX, IFNG, IL2RB, ITK, LCK, NELL2, NKG7, NUSAP1, PCLAF, PRF1, RRM2, STAT4, TRAC |
| **QCMAT** | ADAMDEC1, ADAP2, C15orf48, C5AR1, CCL18, CCRL2, CD300LF, CD36, CD68, CD86, CEBPA, CLEC12A, CSF1R, CSF2RA, CSTA, CXCL16, DMXL2, DOK3, EMILIN2, FCER1G, FLVCR2, FPR1, FPR3, GCA, GJB2, GM2A, HK3, IFIT3, IGSF6, IL1RN, IL4I1, IRF2, KYNU, LILRA6, LILRB1, LILRB2, LILRB4, LOC100996724, LYZ, MCOLN1, MNDA, MREG, MS4A7, NCF2, PILRA, PLA2G7, PRKCD, PTAFR, RNF19B, S100A9, SIGLEC1, SIGLEC7, SLAMF8, SLC15A3, SLC1A3, SLC29A3, SLC38A6, TFEC, THEMIS2, TLR2, TLR8, TYMP, UNC93B1 |

| | |
|---|---|
| **RATs** | ACKR1, ADAM15, ADAMDEC1, ADGRL4, AIF1, AIM2, ANKRD22, AOAH, APLN, APOBEC3A, APOBEC3G, APOBEC3H, APOL1, APOL3, APOL6, ARMH1, B2M, BATF, BCL2A1, BCL6B, BTLA, BTN3A1, BTN3A2, BTN3A3, BUB1, C1QA, C1QB, C2CD4B, CALHM6, CARD16, CCL3, CCL4, CCL4L1, CCL5, CCR5, CD160, CD2, CD244, CD247, CD27, CD28, CD3D, CD3G, CD6, CD72, CD74, CD84, CD86, CD8A, CD8B, CD96, CDCA7, CDH13, CDH5, CENPA, CIITA, CLEC12A, CLEC2D, CLEC7A, CLEC9A, COL13A1, CRTAM, CSF2RA, CST7, CTLA4, CTSS, CX3CL1, CX3CR1, CXCL10, CXCL11, CXCL13, CXCL9, CXCR6, CYBB, CYLD, DUSP2, DUSP4, ECSCR, EOMES, ESM1, ETV7, FAM72A, FASLG, FCGR1A, FCGR1B, FCGR3A, FCN1, FCRL6, FGFBP2, FYB1, GABBR1, GBP1, GBP2, GBP4, GBP5, GCNT1, GIMAP1-GIMAP5, GIMAP4, GIMAP6, GJD3, GNG11, GNLY, GZMA, GZMB, GZMH, GZMK, HAPLN3, HCK, HCP5, HCST, HLA-A, HLA-B, HLA-C, HLA-DMA, HLA-DMB, HLA-DOA, HLA-DPA1, HLA-DPB1, HLA-DQA1, HLA-DRA, HLA-DRB1, HLA-E, HLA-F, HSPA12B, ICAM1, ICAM2, ICOS, IDO1, IFI27, IFNG, IGFLR1, IL15, IL15RA, IL18BP, IL23A, IL2RA, IL2RB, IRF1, ITGAL, ITK, JAKMIP1, KLF4, KLRC1, KLRC3, KLRC4-KLRK1, KLRD1, KLRF1, LAG3, LAIR1, LAIR2, LAP3, LAYN, LCK, LCP2, LILRA1, LILRB1, LILRB2, LILRB4, LPXN, LST1, LYN, LYPD5, MALL, MARCH1, MKI67, MMRN2, MS4A6A, MS4A7, MYB, MYBL1, NCKAP1L, NCR1, NKG7, NLRC5, P2RX7, PCED1B, PDCD1LG2, PECAM1, PLA1A, PLAT, PLEK, |

| | |
|---|---|
| | PREX1, PRF1, PSMB10, PSMB8, PSMB9, PSME2, PSTPIP1, PSTPIP2, PTPN7, PTPRCAP, PVRIG, PYHIN1, RAMP3, RAPGEF5, RARRES3, RASIP1, RNASE2, RNF213, ROBO4, RRM2, RUNX3, S100A3, S1PR1, S1PR5, SCIMP, SELE, SERPINB9, SH2B3, SH2D1A, SH2D1B, SH2D2A, SIRPG, SLA, SLA2, SLAMF6, SLAMF7, SLAMF8, SP140, ST8SIA4, STAT1, STX11, TAP1, TAP2, TBX21, TEK, TIGIT, TLR8, TM4SF1, TM4SF18, TNF, TNFAIP3, TNFAIP8L2, TNFRSF9, TNFSF13B, TOX2, TRAC, TRAFD1, TRBC1, TRBC2, TRDC, TRDV3, UBE2L6, WARS, WIPF1, XCL1, YME1L1, ZAP70, ZBED2, ZNF831 |

| | |
|---|---|
| **Rej-RATs** | AIF1, AOAH, APOL1, APOL3, APOL6, BCL2A1, BTN3A1, BTN3A2, BTN3A3, CALHM6, CARD16, CCL3, CCL4, CCL4L1, CCL5, CD160, CD244, CD247, CD3G, CD74, CD8A, CIITA, CLEC12A, CLEC7A, CRTAM, CST7, CTSS, CXCL10, CXCL11, CXCL9, EOMES, FASLG, FCGR3A, FCN1, FCRL6, FGFBP2, GABBR1, GBP1, GBP2, GBP4, GBP5, GIMAP4, GIMAP6, GNLY, GZMA, GZMB, GZMH, HCK, HCP5, HCST, HLA-A, HLA-B, HLA-C, HLA-DMA, HLA-DMB, HLA-DOA, HLA-DPA1, HLA-DPB1, HLA-DQA1, HLA-DRA, HLA-DRB1, HLA-E, HLA-F, ICAM1, IDO1, IL15, IL15RA, IL18BP, IL2RB, IRF1, ITGAL, KLRC1, KLRC3, KLRC4-KLRK1, KLRD1, LCP2, LILRB1, LILRB2, LST1, LYN, LYPD5, NKG7, NLRC5, PLA1A, PLEK, PRF1, PSMB10, PSMB8, PSMB9, RARRES3, RUNX3, SCIMP, SERPINB9, SLAMF7, STX11, TAP1, TAP2, TBX21, TLR8, TNFRSF9, TRDC, TRDV3, UBE2L6, WARS, XCL1 |
| **TCB** | CD3D, CXCR6, GPR171, NELL2, TRAC |

| | |
|---|---|
| **TCMR-RATs** | ADAMDEC1, AIM2, ANKRD22, AOAH, APOBEC3G, APOBEC3H, ARMH1, BATF, BTLA, BUB1, C1QA, C1QB, CALHM6, CCL5, CCR5, CD2, CD27, CD28, CD3D, CD3G, CD6, CD72, CD84, CD86, CD8A, CD8B, CD96, CDCA7, CENPA, CLEC12A, CLEC2D, CLEC7A, CRTAM, CSF2RA, CTLA4, CXCL13, CXCR6, CYBB, CYLD, DUSP2, DUSP4, EOMES, ETV7, FAM72A, FASLG, FCGR1A, FCGR1B, FYB1, GABBR1, GCNT1, GIMAP1-GIMAP5, GZMK, HAPLN3, HLA-DMB, HLA-DOA, HLA-F, ICOS, IFNG, IGFLR1, IL18BP, IL23A, IL2RA, IL2RB, ITK, JAKMIP1, KLRC4-KLRK1, KLRD1, LAG3, LAIR1, LAIR2, LAP3, LCK, LILRB4, LPXN, MARCH1, MKI67, MS4A6A, MYB, NCKAP1L, P2RX7, PCED1B, PDCD1LG2, PLAAT4, PREX1, PSMB10, PSMB9, PSME2, PSTPIP1, PSTPIP2, PTPN7, PTPRCAP, PVRIG, PYHIN1, RNASE2, RNF213, RRM2, SCIMP, SH2D1A, SH2D2A, SIRPG, SLA, SLA2, SLAMF6, SLAMF8, SP140, ST8SIA4, STAT1, TAP1, TIGIT, TLR8, TNFAIP8L2, TNFRSF9, TNFSF13B, TOX2, TRAC, TRAFD1, TRBC1, TRBC2, WIPF1, YME1L1, ZAP70, ZBED2, ZNF831 |
| **eDSAST** | ACKR1, CDH13, CDH5, COL13A1, GNG11, ICAM2, MALL, PLAT, ROBO4, SOX7, TEK |

**Abbreviations:** AAG, acute antibody-mediated injury-associated gene set; ABMR-RATs, antibody-mediated rejection-associated transcripts; AEG, antibody-mediated rejection-associated endothelial genes; AMAT1, alternative macrophage activation transcripts 1; BAT, B cell-associated transcripts; DAMP, damage-associated molecular pattern transcripts; DSAST, donor-specific antibody-selective transcripts; eDSAST, endothelial donor-specific antibody-selective transcripts; ENDAT, endothelial cell-associated transcripts; FICOL, fibrillar collagen transcripts; GRIT1, gamma-interferon and rejection-induced transcripts 1; GRIT2, gamma-interferon and rejection-induced transcripts 2; GRIT3, new gamma-interferon and rejection-induced transcripts; IIAAG, interferon-inducible acute antibody-mediated gene set; IRITD1, injury- and repair-induced transcripts, early; IRITD3, injury- and repair-induced transcripts, intermediate; IRITD5, injury- and repair-induced transcripts, late; IRRAT, injury-repair response-associated transcripts; KT1, kidney transcripts set 1; KT2, kidney transcripts set 2; MCAT, mast cell-associated transcripts; NKAAG, NK cell-expressed antibody-mediated rejection activity genes; NKB, NK cell transcript burden; PEC, parietal epithelial cell-associated transcripts; QCAT, quantitative cytotoxic T lymphocyte-associated transcripts; QCMAT, quantitative constitutive macrophage-associated transcripts; RATs, rejection-associated transcripts; Rej-RATs, rejection-associated transcripts; TCB, T cell transcript burden; TCMR-RATs, T cell-mediated rejection-associated transcripts

**Supplementary Table S4.** Cell types and abbreviations included in CIBERSORTx deconvolution analysis.

| Abbreviation | Cell Type / Description |
|---|---|
| **PT** | Proximal Tubule |
| **PT injured** | Proximal Tubule (injured) |
| **PEC** | Parietal Epithelial Cells |
| **PE** | Pelvic Epithelium |
| **LOH** | Loop of Henle |
| **DCT** | Distal Convoluted Tubule cells |
| **PC** | Principal Cells |
| **IC A/B** | Intercalated Cells type A and B |
| **vSMp** | Vascular Smooth Muscle cells and pericytes |
| **ECg** | Endothelial glomerular cells |
| **ECvr** | Endothelial cells, vasa recta |
| **ECptc** | Endothelial cells, peritubular capillary |
| **Podocytes** | Podocytes |
| **Macrophages** | Macrophages |
| **Monocytes** | Monocytes |
| **B cells** | B cells |
| **Plasma cells** | Plasma cells |
| **CD4 T cells** | CD4 T cells |
| **CD8 T cells** | CD8 T cells |
| **NK cells** | Natural Killer cells |
| **cDC1** | Conventional dendritic cell 1 |
| **cDC2** | Conventional dendritic cell 2 |
| **pDC** | Plasmacytoid dendritic cell |

**Supplementary Table S1.** Agreement of individual Banff lesion scores based on Gwet AC2.

| Banff lesion score | % agreement | % chance | Gwet AC2 | 95% CI |
|---|---|---|---|---|
| **g-score** | 0.83 | 0.43 | 0.71 | 0.61-0.81 |
| **cg-score** | 0.89 | 0.31 | 0.84 | 0.77-0.92 |
| **mm-score** | 0.80 | 0.41 | 0.66 | 0.57-0.76 |
| **i-score** | 0.82 | 0.50 | 0.64 | 0.54-0.74 |
| **t-score** | 0.81 | 0.50 | 0.63 | 0.53-0.73 |
| **ti-score** | 0.83 | 0.57 | 0.60 | 0.55-0.65 |
| **ci-score** | 0.81 | 0.54 | 0.58 | 0.49-0.67 |
| **ct-score** | 0.81 | 0.53 | 0.60 | 0.52-0.68 |
| **IFTA** | 0.80 | 0.55 | 0.57 | 0.49-0.65 |
| **i-IFTA** | 0.72 | 0.57 | 0.36 | 0.24-0.48 |
| **t-IFTA** | 0.80 | 0.47 | 0.61 | 0.49-0.73 |
| **ptc-score** | 0.74 | 0.50 | 0.49 | 0.34-0.64 |
| **v-score** | 0.92 | 0.31 | 0.88 | 0.84-0.93 |
| **cv-score** | 0.83 | 0.59 | 0.57 | 0.50-0.64 |
| **ah-score** | 0.77 | 0.53 | 0.51 | 0.38-0.64 |

**Abbreviations:** *i*, interstitial inflammation; *t*, tubulitis; *v*, intimal arteritis; *g*, glomerulitis; *ptc*, peritubular capillaritis; *C4d*, C4d staining; *ci*, interstitial fibrosis; *ct*, tubular atrophy; *cv*, vascular fibrous intimal thickening; *cg*, glomerular basement membrane double contours; *mm*, mesangial matrix expansion; *ah*, arteriolar hyalinosis; *aah*, alternative arteriolar hyalinosis / hyaline arteriolar thickening score; *ti*, total inflammation; *i-IFTA*, inflammation in areas of interstitial fibrosis and tubular atrophy; *t-IFTA*, tubulitis in areas of interstitial fibrosis and tubular atrophy.

**Supplementary Table S2.** Mixed-effects logistic regression models of pairwise diagnostic agreement.

| Predictor | β coefficient | SE | OR | 95% CI for OR | p-value |
|---|---|---|---|---|---|
| **Base model** | | | | | |
| **Experience difference (per year)** | **0.01** | **0.001** | **1.01** | **1.01 – 1.01** | **< 0.001** |
| **Same continent** | **0.05** | **0.02** | **1.05** | **1.01 – 1.09** | **0.01** |
| **Core lesion model** | | | | | |
| **v-score** | **-1.89** | **0.02** | **0.15** | **0.14 - 0.16** | **< 0.001** |
| **ptc-score** | **-0.63** | **0.01** | **0.54** | **0.52 – 0.55** | **< 0.001** |
| **i-score** | **-0.64** | **0.02** | **0.53** | **0.52 – 0.55** | **< 0.001** |
| **t-score** | **-0.80** | **0.02** | **0.45** | **0.44 – 0.46** | **< 0.001** |
| **ci-score** | **-0.08** | **0.02** | **0.92** | **0.89 – 0.96** | **< 0.001** |
| **ct-score** | **-0.07** | **0.02** | **0.93** | **0.90 – 0.97** | **0.001** |
| **Experience difference (per year)** | **0.01** | **0.001** | **1.01** | **1.00 – 1.01** | **< 0.001** |
| **Same continent** | -0.04 | 0.02 | 0.96 | 0.92 – 1.00 | 0.06 |
| **Full lesion model** | | | | | |
| **v-score** | **-1.88** | **0.03** | **0.15** | **0.14 – 0.16** | **< 0.001** |
| **ptc-score** | **-0.62** | **0.01** | **0.54** | **0.52 – 0.55** | **< 0.001** |
| **i-score** | **-0.60** | **0.02** | **0.55** | **0.53 – 0.56** | **< 0.001** |
| **t-score** | **-0.77** | **0.02** | **0.46** | **0.45 – 0.48** | **< 0.001** |

| | | | | | |
|---|---|---|---|---|---|
| **g-score** | **-0.05** | **0.01** | **0.95** | **0.93 – 0.98** | **< 0.001** |
| **cg-score** | **-0.05** | **0.02** | **0.95** | **0.92 – 0.99** | **0.01** |
| **mm-score** | **-0.05** | **0.01** | **0.95** | **0.93 – 0.98** | **< 0.001** |
| **ti-score** | **-0.01** | **0.001** | **0.99** | **0.99 – 1.00** | **< 0.001** |
| **IFTA** | **0.01** | **0.001** | **1.01** | **1.00 – 1.01** | **< 0.001** |
| **ci-score** | **-0.05** | **0.02** | **0.96** | **0.91 – 1.00** | **0.04** |
| **ct-score** | **-0.08** | **0.02** | **0.93** | **0.89 – 0.97** | **< 0.001** |
| **i-IFTA** | **-0.04** | **0.01** | **0.96** | **0.94 – 0.98** | **< 0.001** |
| **t-IFTA** | **-0.33** | **0.02** | **0.72** | **0.70 – 0.74** | **< 0.001** |
| **cv-score** | **-0.22** | **0.01** | **0.81** | **0.79 – 0.83** | **< 0.001** |
| **ah-score** | 0.001 | 0.01 | 1.00 | 0.98 – 1.03 | 0.94 |
| **Experience difference (per year)** | **0.01** | **0.001** | **1.01** | **1.00 – 1.01** | **< 0.001** |
| **Same continent** | **-0.04** | **0.02** | **0.96** | **0.92 – 1.00** | **0.04** |

Results are shown for mixed-effects logistic regression models with case included as a random intercept. The outcome was whether a reader pair assigned the same final diagnosis to a biopsy. Odds ratios (ORs) are shown per 1-unit increase in absolute between-reader lesion score difference or per 1-year increase in experience difference. ORs below 1 indicate lower odds of diagnostic agreement. The base model included experience difference and shared continent. The core lesion model included selected key Banff lesions (v, ptc, i, t, ci, and ct). Significant associations ($p < 0.05$) are shown in bold.

**Abbreviations:** *i*, interstitial inflammation; *t*, tubulitis; *v*, intimal arteritis; *g*, glomerulitis; *ptc*, peritubular capillaritis; *C4d*, C4d staining; *ci*, interstitial fibrosis; *ct*, tubular atrophy; *cv*, vascular fibrous intimal thickening; *cg*, glomerular basement membrane double contours; *mm*, mesangial matrix expansion; *ah*, arteriolar hyalinosis; *aah*, alternative arteriolar hyalinosis / hyaline arteriolar thickening score; *ti*, total inflammation; *i-IFTA*, inflammation in areas of interstitial fibrosis and tubular atrophy; *t-IFTA*, tubulitis in areas of interstitial fibrosis and tubular atrophy, SE, standard error; OR, odds ratio; CI, confidence interval.

**Supplementary Table S3.** Unadjusted diagnostic agreement by geographic subgroup in the full reader cohort.

| | Readers, n | Cases, n | Gwet AC1 | 95% CI | Mean difference | 95% CI of difference |
|---|---|---|---|---|---|---|
| **Continent / comparison** | | | | | | |
| **Agreement by continent** | | | | | | |
| **Asia** | 12 | 36 | 0.51 | 0.42-0.59 | - | - |
| **Americas** | 24 | 36 | 0.55 | 0.46-0.64 | - | - |
| **Europe** | 24 | 36 | 0.58 | 0.49-0.66 | - | - |
| **Pairwise bootstrap comparison** | | | | | | |
| **Americas vs Europe** | - | - | - | - | -0.03 | -0.061 – 0.01 |
| **Americas vs Asia** | - | - | - | - | **0.04** | **0.004 - 0.08** |
| **Europe vs Asia** | - | - | - | - | **0.07** | **0.04 - 0.11** |

Agreement in final diagnostic classification is shown as Gwet AC1 with 95% CI for readers grouped by continent. Pairwise differences were estimated by bootstrap resampling and are reported as mean differences in AC1 with corresponding 95% CI. The Americas consist of both North and South America. Continents with fewer than 5 readers were excluded from bootstrap pairwise comparisons: Oceania (n = 4), Africa (n = 3). All 67 readers had complete geography data, and each subgroup evaluated the full set of 36 biopsies. Significant comparisons are shown in bold (CI does not contain 0).

**Abbreviations:** CI, confidence interval

**Supplementary Table S4.** Balanced-bootstrap analysis of diagnostic agreement by geographic subgroup.

| | Original readers, n | Readers sampled per bootstrap, n | Gwet's AC1 | 95% CI | Mean differences | 95% CI of difference |
|---|---|---|---|---|---|---|
| **Continent / comparison** | | | | | | |
| **Balanced bootstrap agreement by continent** | | | | | | |
| **Africa** | 3 | 3 | 0.35 | 0.22-0.48 | - | - |
| **Asia** | 12 | 3 | 0.49 | 0.24-0.77 | - | - |
| **Americas** | 24 | 3 | 0.54 | 0.31-0.79 | - | - |
| **Oceania** | 4 | 3 | 0.56 | 0.39-0.72 | - | - |
| **Europe** | 24 | 3 | 0.57 | 0.30-0.77 | - | - |
| **Pairwise balanced-bootstrap comparison** | | | | | | |
| **Americas vs Europe** | - | - | - | - | -0.03 | -0.35 – 0.34 |
| **Americas vs Asia** | - | - | - | - | 0.04 | -0.33 – 0.38 |
| **Americas vs Oceania** | - | - | - | - | -0.02 | -0.30 – 0.26 |
| **Americas vs Africa** | - | - | - | - | 0.18 | -0.07 – 0.45 |
| **Europe vs Asia** | - | - | - | - | 0.07 | -0.31 – 0.40 |
| **Europe vs Oceania** | - | - | - | - | 0.01 | -0.29 – 0.25 |
| **Europe vs Africa** | - | - | - | - | 0.22 | -0.07 – 0.45 |
| **Asia vs Oceania** | - | - | - | - | -0.07 | -0.35 – 0.23 |
| **Asia vs Africa** | - | - | - | - | 0.14 | -0.12 – 0.42 |
| **Oceania vs Africa** | - | - | - | - | **0.21** | **0.03 – 0.40** |

Agreement in final diagnostic classification is shown as Gwet AC1 with 95% CI for continent-level geographic subgroups in a balanced-bootstrap sensitivity analysis; in each bootstrap iteration 3 readers per continent were sampled to standardize subgroup size across Africa, Asia, the Americas, Oceania and Europe. Pairwise differences between continents are reported as mean differences in AC1 with corresponding 95% CI. Significant comparisons are shown in bold (CI does not contain 0).

**Abbreviations:** CI, confidence interval

**Supplementary Table S5.** Unadjusted diagnostic agreement by reader experience (in years) in the full reader cohort.

| | Readers, n | Cases, n | Gwet AC1 | 95% CI | Mean difference | 95% CI of difference |
|---|---|---|---|---|---|---|
| **Experience group / comparison** | | | | | | |
| **Agreement by experience group** | | | | | | |
| **< 5 years** | 20 | 36 | 0.58 | 0.49-0.67 | - | - |
| **5 – 10 years** | 25 | 36 | 0.49 | 0.41-0.58 | - | - |
| **> 10 years** | 22 | 36 | 0.60 | 0.51-0.68 | - | - |
| **Pairwise bootstrap comparison** | | | | | | |
| **5 – 10 years vs > 10 years** | - | - | - | - | **-0.10** | **-0.13 to -0.07** |
| **5 – 10 years vs < 5 years** | - | - | - | - | **-0.09** | **-0.12 to -0.06** |
| **> 10 years vs < 5 years** | - | - | - | - | 0.01 | -0.03 – 0.06 |

Agreement in final diagnostic classification is shown as Gwet AC1 with 95% CI for readers grouped by experience. Pairwise differences were estimated by bootstrap resampling and are reported as mean differences in AC1 with corresponding 95% CI. All 67 readers had complete experience data, and each subgroup evaluated the full set of 36 biopsies. Significant comparisons are shown in bold (CI does not contain 0).

**Abbreviations:** CI, confidence interval

**Supplementary Table S6**. Variance explained by the first 10 principal components (PC) of reader similarity.

| Principal component | SD | Proportion of variance | Cumulative proportion |
|---|---|---|---|
| **PC1** | 0.85 | 0.60 | 0.60 |
| **PC2** | 0.39 | 0.13 | 0.73 |
| **PC3** | 0.22 | 0.04 | 0.77 |
| **PC4** | 0.19 | 0.03 | 0.80 |
| **PC5** | 0.17 | 0.02 | 0.82 |
| **PC6** | 0.17 | 0.02 | 0.84 |
| **PC7** | 0.15 | 0.02 | 0.86 |
| **PC8** | 0.14 | 0.02 | 0.88 |
| **PC9** | 0.13 | 0.01 | 0.89 |
| **PC10** | 0.12 | 0.01 | 0.90 |

**Supplementary Table S7.** Correlation of variables with the first four principal components (PC1 – PC4).

| | PC1 | p-value | PC2 | p-value | PC3 | p-value | PC4 | p-value |
|---|---|---|---|---|---|---|---|---|
| **Diagnosis (grouped)** | | | | | | | | |
| Any rejection | **0.70** | **< 0.001** | **0.45** | **< 0.001** | **0.26** | **0.03** | -0.10 | 0.44 |
| No rejection | **-0.70** | **< 0.001** | **-0.45** | **< 0.001** | **-0.26** | **0.03** | 0.10 | 0.44 |
| Borderline | **0.68** | **< 0.001** | 0.06 | 0.61 | 0.09 | 0.45 | **0.36** | **0.002** |
| Any AMR | **-0.28** | **0.02** | -0.02 | 0.86 | 0.17 | 0.16 | **0.37** | **0.002** |
| Any TCMR | 0.24 | 0.05 | **-0.58** | **< 0.001** | **0.54** | **< 0.001** | **-0.31** | **0.01** |
| Mixed | 0.18 | 0.14 | **0.78** | **< 0.001** | -0.22 | 0.07 | **-0.27** | **0.03** |
| **Diagnosis (specific)** | | | | | | | | |
| No rejection | **-0.70** | **< 0.001** | **-0.45** | **< 0.001** | **-0.26** | **0.03** | 0.10 | 0.44 |
| Borderline | **0.68** | **< 0.001** | 0.06 | 0.61 | 0.09 | 0.45 | **0.36** | **0.002** |
| AMR (chronic active) | **-0.51** | **< 0.001** | 0.22 | 0.07 | 0.08 | 0.54 | **0.37** | **0.002** |
| AMR (probable) | **0.32** | **0.01** | -0.16 | 0.21 | 0.09 | 0.48 | 0.02 | 0.88 |
| TCMR (chronic active) | 0.18 | 0.14 | **-0.52** | **< 0.001** | **0.43** | **< 0.001** | **-0.44** | **< 0.001** |
| Mixed | 0.18 | 0.14 | **0.78** | **< 0.001** | -0.22 | 0.07 | **-0.27** | **0.03** |
| AMR (active) | 0.11 | 0.36 | **-0.38** | **0.002** | 0.16 | 0.19 | -0.03 | 0.79 |
| TCMR (acute) | 0.07 | 0.57 | **-0.43** | **< 0.001** | 0.51 | < 0.001 | 0.11 | 0.40 |
| **Indices** | | | | | | | | |

| | | | | | | | | |
|---|---|---|---|---|---|---|---|---|
| TCMR index | **0.64** | **< 0.001** | **0.44** | **< 0.001** | 0.15 | 0.22 | -0.19 | 0.13 |
| Activity index | **0.42** | **< 0.001** | **0.70** | **< 0.001** | 0.05 | 0.67 | -0.20 | 0.10 |
| Chronicity index | 0.09 | 0.49 | 0.17 | 0.16 | -0.23 | 0.06 | 0.001 | 0.99 |
| AMR index | 0.08 | 0.52 | **0.71** | **< 0.001** | -0.07 | 0.58 | -0.11 | 0.36 |
| **Lesion scores** | | | | | | | | |
| i-score | **0.62** | **< 0.001** | **0.25** | **0.04** | 0.14 | 0.26 | -0.08 | 0.53 |
| t-score | **0.45** | **< 0.001** | **0.53** | **< 0.001** | 0.18 | 0.14 | -0.19 | 0.13 |
| v-score | **0.40** | **< 0.001** | **0.36** | **0.003** | 0.13 | 0.29 | **-0.28** | **0.02** |
| t-IFTA | 0.24 | 0.05 | **0.27** | **0.03** | **-0.25** | **0.04** | **-0.26** | **0.04** |
| ci-score | 0.23 | 0.07 | 0.05 | 0.68 | **-0.25** | **0.04** | 0.01 | 0.91 |
| IFTA | 0.18 | 0.14 | 0.12 | 0.33 | **-0.29** | **0.02** | -0.07 | 0.57 |
| cv-score | -0.18 | 0.15 | -0.01 | 0.96 | -0.14 | 0.27 | -0.003 | 0.98 |
| ct-score | 0.08 | 0.54 | 0.11 | 0.38 | -0.15 | 0.23 | -0.10 | 0.43 |
| g-score | 0.07 | 0.58 | **0.51** | **< 0.001** | 0.02 | 0.89 | -0.03 | 0.83 |
| i-IFTA | 0.03 | 0.81 | **0.28** | **0.02** | -0.20 | 0.11 | **-0.32** | **0.01** |
| ptc-score | -0.02 | 0.89 | **0.79** | **< 0.001** | -0.07 | 0.55 | -0.25 | 0.05 |
| **Reader metadata** | | | | | | | | |
| Years of experience | -0.000 | 1.00 | 0.11 | 0.39 | -0.10 | 0.40 | 0.15 | 0.24 |

All variables were correlated with the first four principal components (PC). Values are Spearman correlation coefficients with corresponding p-values. Rows are ordered by the absolute magnitude of the PC1 correlation, from strongest to weakest. Significant associations ($p < 0.05$) are shown in bold.

**Abbreviations**: PC, principal components; AMR, antibody-mediated rejection; TCMR, T cell-mediated rejection; *i*, interstitial inflammation; *t*, tubulitis; *v*, intimal arteritis; *t*-IFTA, tubulitis in areas of interstitial fibrosis and tubular atrophy; *ci*, interstitial fibrosis; IFTA, interstitial fibrosis and tubular atrophy; *cv*, vascular fibrous intimal thickening; *ct*, tubular atrophy; *g*, glomerulitis; *i*-IFTA, inflammation in areas of interstitial fibrosis and tubular atrophy; *ptc*, peritubular capillaritis

**Supplementary Table S8.** All GSVA pathway associated with PC1 and PC2.

| Pathway | Readers, n | Correlation coefficient | p-value | FDR-adjusted p-value |
|---|---|---|---|---|
| **PC1** | | | | |
| QCAT | 67 | **-0.59** | **<0.001** | **<0.001** |
| TCMR-RATs | 67 | **-0.57** | **<0.001** | **<0.001** |
| NKAAG | 67 | **-0.57** | **<0.001** | **<0.001** |
| Rej-RATs | 67 | **-0.54** | **<0.001** | **<0.001** |
| RATs | 67 | **-0.54** | **<0.001** | **<0.001** |
| AAG | 67 | **-0.53** | **<0.001** | **<0.001** |
| QCMAT | 67 | **-0.53** | **<0.001** | **<0.001** |
| TCB | 67 | **-0.52** | **<0.001** | **<0.001** |
| IIAAG | 67 | **-0.52** | **<0.001** | **<0.001** |
| GRIT2 | 67 | **-0.51** | **<0.001** | **<0.001** |
| BAT | 67 | **-0.50** | **<0.001** | **<0.001** |
| ABMR-RATs | 67 | **-0.50** | **<0.001** | **<0.001** |
| GRIT3 | 67 | **-0.45** | **<0.001** | **<0.001** |
| IRRAT | 67 | **-0.49** | **<0.001** | **<0.001** |
| DAMP | 67 | **-0.48** | **<0.001** | **<0.001** |
| NKB | 67 | **-0.48** | **<0.001** | **<0.001** |
| GRIT1 | 67 | **-0.46** | **<0.001** | **<0.001** |
| KT1 | 67 | **0.43** | **<0.001** | **<0.001** |
| DSAST | 67 | **-0.42** | **<0.001** | **<0.001** |
| AMAT1 | 67 | **-0.41** | **<0.001** | **<0.001** |
| IRITD5 | 67 | **-0.36** | **0.003** | **0.004** |
| KT2 | 67 | **0.35** | **0.004** | **0.01** |
| IRITD1 | 67 | **0.34** | **0.01** | **0.01** |
| AEG | 67 | **-0.33** | **0.01** | **0.01** |
| eDSAST | 67 | **-0.32** | **0.01** | **0.01** |
| ENDAT | 67 | **-0.27** | **0.03** | **0.02** |
| IRITD3 | 67 | **-0.26** | **0.04** | **0.04** |
| MCAT | 67 | -0.14 | 0.25 | 0.26 |
| FICOL | 67 | -0.10 | 0.43 | 0.43 |
| **PC2** | | | | |
| ABMR-RATs | 64 | **-0.43** | **<0.001** | **0.01** |
| IIAAG | 64 | **-0.40** | **0.001** | **0.01** |
| DSAST | 64 | **-0.34** | **0.001** | **0.01** |
| GRIT1 | 64 | **-0.37** | **0.003** | **0.02** |
| AAG | 64 | **-0.37** | **0.003** | **0.02** |
| GRIT3 | 64 | **-0.36** | **0.003** | **0.02** |
| NKAAG | 64 | **-0.33** | **0.01** | **0.04** |
| NKB | 64 | **-0.32** | **0.01** | **0.04** |

| | | | | |
|---|---|---|---|---|
| DAMP | 64 | -0.28 | 0.02 | 0.08 |
| RATs | 64 | -0.26 | 0.04 | 0.11 |
| eDSAST | 64 | -0.25 | 0.05 | 0.12 |
| Rej-RATs | 64 | -0.24 | 0.05 | 0.13 |
| GRIT2 | 64 | -0.24 | 0.06 | 0.14 |
| IRITD3 | 64 | -0.21 | 0.10 | 0.20 |
| BAT | 64 | -0.20 | 0.12 | 0.22 |
| AMAT1 | 64 | -0.18 | 0.15 | 0.28 |
| IRRAT | 64 | 0.16 | 0.21 | 0.35 |
| FICOL | 64 | -0.16 | 0.22 | 0.35 |
| IRITD5 | 64 | -0.14 | 0.26 | 0.39 |
| TCMR-RATs | 64 | -0.14 | 0.27 | 0.39 |
| MCAT | 64 | 0.13 | 0.30 | 0.41 |
| KT2 | 64 | 0.12 | 0.34 | 0.44 |
| IRITD1 | 64 | -0.11 | 0.39 | 0.49 |
| QCMAT | 64 | -0.08 | 0.53 | 0.65 |
| QCAT | 64 | -0.06 | 0.63 | 0.74 |
| KT1 | 64 | 0.06 | 0.67 | 0.75 |
| ENDAT | 64 | 0.03 | 0.80 | 0.86 |
| TCB | 64 | 0.02 | 0.85 | 0.87 |
| AEG | 64 | -0.02 | 0.87 | 0.87 |

Rows are grouped by principal component and ordered by the absolute magnitude of the correlation within each component. Correlations were calculated using Spearman correlation. Significant associations (FDR p-value < 0.05) are shown in bold.

**Abbreviations**: AAG, acute antibody-mediated injury-associated gene set; ABMR-RATs, antibody-mediated rejection-associated transcripts; AEG, antibody-mediated rejection-associated endothelial genes; AMAT1, alternative macrophage activation transcripts 1; BAT, B cell-associated transcripts; DAMP, damage-associated molecular pattern transcripts; DSAST, donor-specific antibody-selective transcripts; eDSAST, endothelial donor-specific antibody-selective transcripts; ENDAT, endothelial cell-associated transcripts; FICOL, fibrillar collagen transcripts; GRIT1, gamma-interferon and rejection-induced transcripts 1; GRIT2, gamma-interferon and rejection-induced transcripts 2; GRIT3, new gamma-interferon and rejection-induced transcripts; IIAAG, interferon-inducible acute antibody-mediated gene set; IRITD1, injury- and repair-induced transcripts, early; IRITD3, injury- and repair-induced transcripts, intermediate; IRITD5, injury- and repair-induced transcripts, late; IRRAT, injury-repair response-associated transcripts; KT1, kidney transcripts set 1; KT2, kidney transcripts set 2; MCAT, mast cell-associated transcripts; NKAAG, NK cell-expressed antibody-mediated rejection activity genes; NKB, NK cell transcript burden; PEC, parietal epithelial cell-associated transcripts; QCAT, quantitative cytotoxic T lymphocyte-associated transcripts; QCMAT, quantitative constitutive macrophage-associated transcripts; RATs, rejection-associated transcripts; Rej-RATs, rejection-associated transcripts; TCB, T cell transcript burden; TCMR-RATs, T cell-mediated rejection-associated transcripts

**Supplementary Figure S1. Correlation structure of pairwise Banff lesion interpretation differences.**

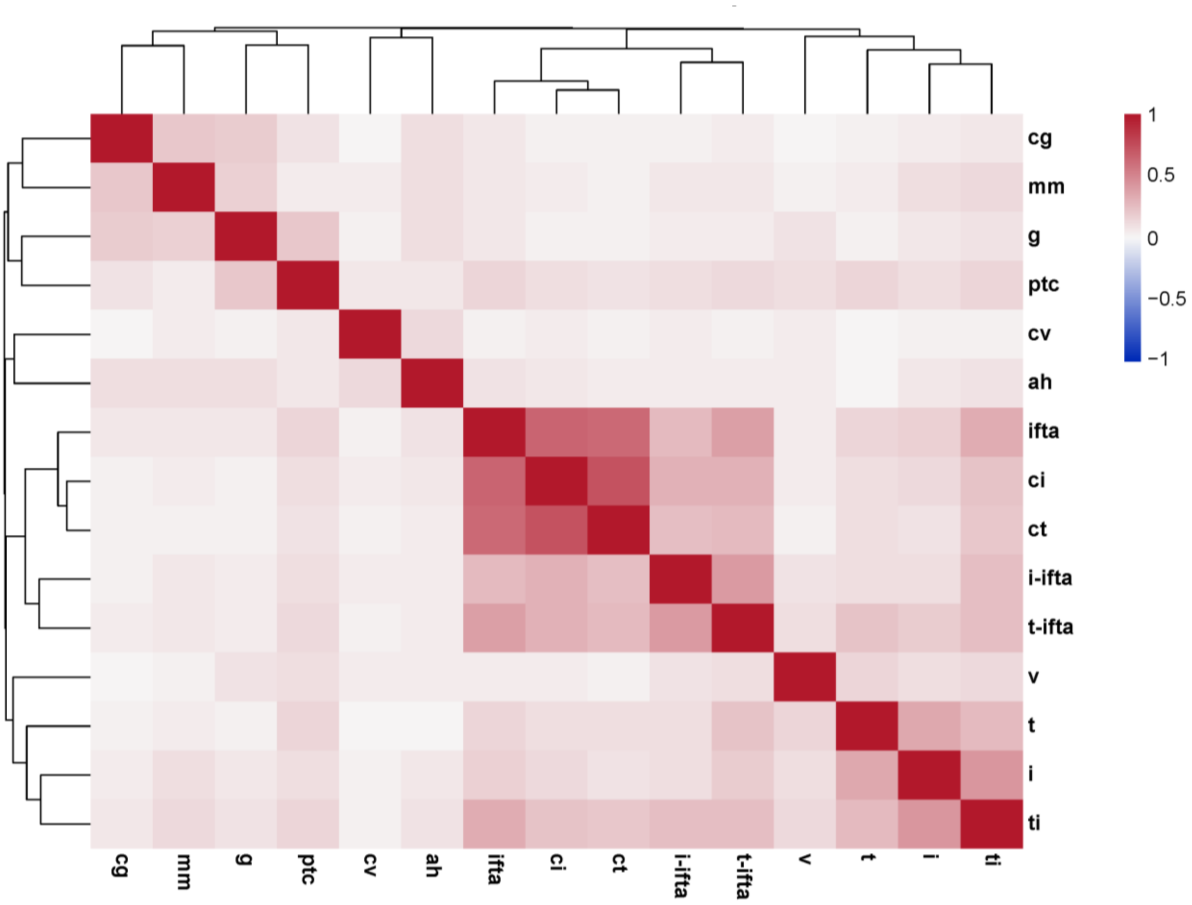

Heatmap showing correlations between pairwise reader differences in Banff lesion scores across biopsies. Rows and columns represent individual Banff lesions, and colors indicate the strength and direction of correlation between lesion-score differences. Hierarchical clustering was applied to visualize groups of lesions with similar disagreement patterns. Chronic parenchymal injury lesions, including ci, ct, IFTA, i-IFTA, and t-IFTA, showed correlated disagreement patterns, while tubulointerstitial inflammatory lesions, including i, t, and ti, clustered separately. These findings indicate that reader disagreement was not independent across Banff lesions but showed structured correlation across biologically and diagnostically related lesion compartments.

**Abbreviations**: *ah*, arteriolar hyalinosis; *cg*, transplant glomerulopathy; *ci*, interstitial fibrosis; *ct*, tubular atrophy; *cv*, vascular fibrous intimal thickening; *g*, glomerulitis; *i*, interstitial inflammation; *i-IFTA*, inflammation in areas of interstitial fibrosis and tubular atrophy; *IFTA*, interstitial fibrosis and tubular atrophy; *mm*, mesangial matrix expansion; *ptc*, peritubular capillaritis; *t*, tubulitis; *t-IFTA*, tubulitis in areas of interstitial fibrosis and tubular atrophy; *ti*, total inflammation; *v*, intimal arteritis.

**Supplementary Figure S2. Diagnostic agreement by geographic region and reader experience.**

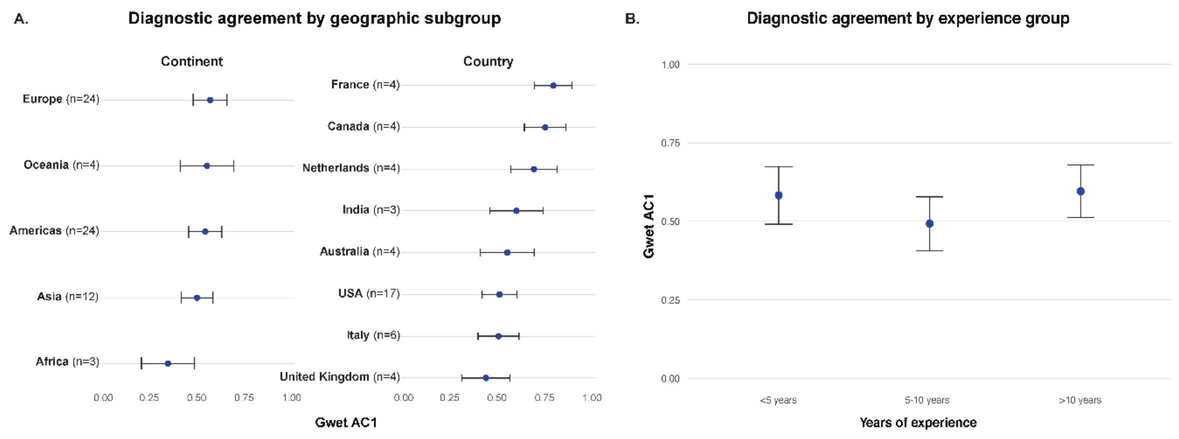

Gwet's AC1 estimates for final Banff diagnostic classification stratified by reader subgroup. Points indicate subgroup-specific agreement estimates and error bars indicate 95% confidence intervals. **A.** Agreement estimates by geographic subgroup, shown at the continent level and, where sufficient reader representation was available, at the country level. Continent-level analyses included Africa, Asia, the Americas, Oceania, and Europe; country-level estimates are shown for countries contributing multiple readers. Agreement estimates varied across geographic subgroups, with numerically lower agreement in Asia and higher agreement in Europe. However, subgroup comparisons were exploratory, and differences between the largest geographic groups, including Europe and the Americas, were not statistically significant in bootstrap comparisons. **B.** Agreement estimates stratified by years of renal pathology experience. Agreement was numerically lowest among readers with 5–10 years of experience and higher among readers with <5 years and >10 years of experience. These patterns did not indicate a consistent monotonic relationship between experience and diagnostic agreement. Overall, subgroup analyses suggested that geographic region and years of experience were not the primary determinants of diagnostic variability. Results should be interpreted descriptively, particularly for subgroups with small reader numbers.
**Abbreviations**: AC1, agreement coefficient 1; CI, confidence interval.

**Supplementary Figure S3. Reader interpretive space derived from the pairwise diagnostic agreement matrix.**

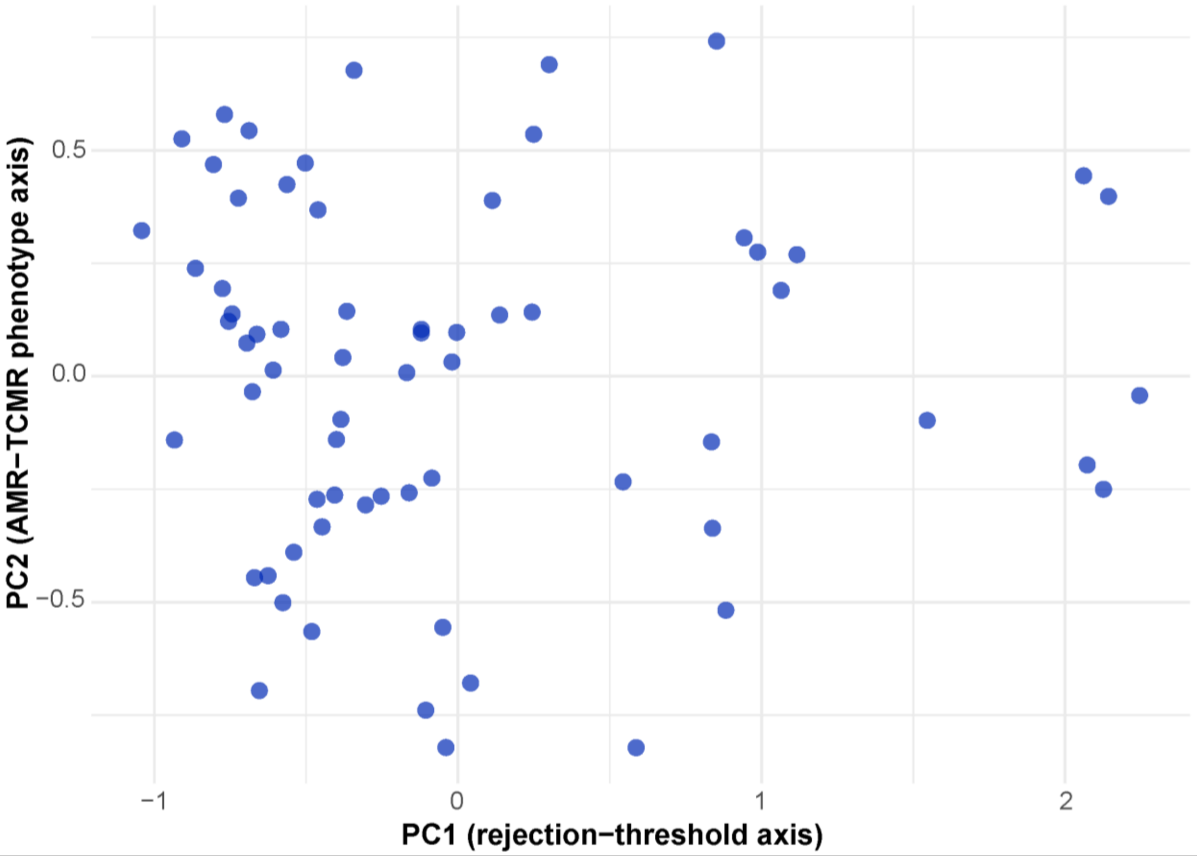

Principal component analysis (PCA) plot of reader coordinates derived from the pairwise Gwet's AC1 similarity matrix for final Banff diagnostic classification. Each point represents an individual reader. PC1 represents the dominant rejection-calling threshold axis, reflecting variation in readers' tendency to assign rejection-related diagnoses, particularly in biopsies with tubulointerstitial inflammatory injury. PC2 represents an AMR/MVI-oriented phenotypic classification axis, reflecting variation in how readers resolved rejection-type biopsies toward antibody-mediated, microvascular inflammation, mixed-rejection, or T cell-mediated rejection patterns. The distribution of readers in this space supports the presence of continuous interpretive variation rather than discrete reader clusters.

**Abbreviations:** AC1, agreement coefficient 1; AMR, antibody-mediated rejection; PC, principal component; TCMR, T cell-mediated rejection.
